# All-cause mortality among patients treated with repurposed antivirals and antibiotics for COVID-19 in Mexico City: A Real-World Observational Study

**DOI:** 10.1101/2020.10.13.20211797

**Authors:** Javier Mancilla-Galindo, Jorge Óscar García-Méndez, Jessica Márquez-Sánchez, Rodrigo Estefano Reyes-Casarrubias, Eduardo Aguirre-Aguilar, Héctor Isaac Rocha-González, Ashuin Kammar-García

**Author notes:** Corresponding author: A. Kammar-García. Vasco de Quiroga 15, Tlalpan, Col. Belisario Domínguez Sección XVI, CP 14080 Ciudad de México, México. **Principal investigator statement:** The collection and public release of datasets used for this study was conducted and authorized by the Secretariat of Health of Mexico City. Since this is a retrospective analysis of an open dataset, no Principal Investigator has been identified and listed as an author. **Funding:** This research did not receive any specific grant from funding agencies in the public, commercial, or not-for-profit sectors. **Declarations of interest:** All authors declare no competing interests. **Data availability statement:** The data that support the findings of this study are openly available in the Open Data Platform of Mexico City’s Government at https://datos.cdmx.gob.mx/explore/dataset/base-covid-sinave/information/, reference number 19, and the Directorate General of Epidemiology of Mexico City’s Weekly Epidemiological Surveillance Reports of Influenza 2020 at https://www.gob.mx/salud/documentos/informes-semanales-para-la-vigilancia-epidemiologica-de-influenza-2020, reference number 21. **Ethics statement:** This is a retrospective analysis of an open-source dataset of patients evaluated for suspected COVID-19 in Mexico City. The Ministry of Health of Mexico City approved the collection and release of publicly available datasets.

## Abstract

**Aim:** To evaluate all-cause mortality risk in patients with laboratory-confirmed COVID-19 in Mexico City treated with repurposed antivirals and antibiotics.

**Methods:** This real-world retrospective cohort study contemplated 395,343 patients evaluated for suspected COVID-19 between February 24 and September 14, 2020 in 688 primary-to-tertiary medical units in Mexico City. Patients were included with a positive RT-PCR for SARS-CoV-2; those receiving unspecified antivirals, excluded; and antivirals prescribed in <30 patients, eliminated. Survival and mortality risks were determined for patients receiving antivirals, antibiotics, both, or none.

**Results:** 136,855 patients were analyzed; mean age 44.2 (SD:16.8) years; 51.3% were men. 16.6% received antivirals (3%), antibiotics (10%), or both (3.6%). Antivirals studied were Oseltamivir (n=8414), Amantadine (n=319), Lopinavir-Ritonavir (n=100), Rimantadine (n=61), Zanamivir (n=39), and Acyclovir (n=36). Survival with antivirals (73.7%, p<0.0001) and antibiotics (85.8%, p<0.0001) was lower than no antiviral/antibiotic (93.6%). After multivariable adjustment, increased risk of death occurred with antivirals (HR=1.72, 95%CI:1.61-1.84) in ambulatory (HR=4.7, 95%CI:3.94-5.62) and non-critical (HR=2.03, 95%CI:1.86-2.21) patients. Oseltamivir increased mortality risk in the general population (HR=1.72, 95%CI:1.61-1.84), ambulatory (HR=4.79, 95%CI:4.01-5.75), non-critical (HR=2.05, 95%CI:1.88-2.23), and pregnancy (HR=8.35, 95%CI:1.77-39.30); as well as hospitalized (HR=1.13, 95%CI:1.01-1.26) and critical patients (HR:1.22, 95%CI:1.05-1.43) after propensity score-matching. Antibiotics were a risk factor in general population (HR=1.13, 95%CI:1.08-1.19) and pediatrics (HR=4.22, 95%CI:2.01-8.86), but a protective factor in hospitalized (HR=0.81, 95%CI:0.77-0.86) and critical patients (HR=0.67, 95%CI:0.63-0.72).

**Conclusions:** No significant benefit for repurposed antivirals was observed; oseltamivir was associated with increased mortality. Antibiotics increased mortality risk in the general population but may increase survival in hospitalized and critical patients.

**WHAT IS ALREADY KNOWN:**

- Current recommendations for using repurposed antivirals and antibiotics for COVID-19 are conflicting.
- Few antivirals (i.e. lopinavir-ritonavir) have been shown to provide no additional benefit for COVID-19 in clinical trials; other antivirals may be having widespread use in real-world settings without formal assessment in clinical trials.
- Real-world use of repurposed antivirals and antibiotics for COVID-19 in population-based studies have not been performed; important populations have been left largely understudied (ambulatory patients, pregnant women, and pediatrics).

**WHAT THIS STUDY ADDS:**

- This is the first real-world observational study evaluating amantadine, rimantadine, zanamivir, and acyclovir for COVID-19; no registered studies to evaluate these drugs exist. Only one study has evaluated risk of death for oseltamivir. Lopinavir-ritonavir have been previously evaluated in clinical trials.
- Repurposed antivirals and antibiotics were commonly prescribed in 688 ambulatory units and hospitals of Mexico City despite unclear recommendations for their use out of clinical trials.
- Oseltamivir was associated with increased mortality risk; other repurposed antivirals (zanamivir, amantadine, rimantadine, and acyclovir) had no significant and consistent impact on mortality. Antibiotics were associated with increased mortality risk in the general population but may increase survival in hospitalized and critical patients.

## INTRODUCTION

The severe acute respiratory syndrome coronavirus 2 (SARS-CoV-2) is the etiologic agent of the coronavirus disease (COVID-19) pandemic, one of the most devastating infectious diseases of this century. Non-pharmacological interventions are the most effective means of limiting the impact of COVID-19.^1^ However, several countries have not been able to contain the disease.^2^

One of the main strategies for finding ways to combat COVID-19 is drug repurposing since developing novel antivirals against SARS-CoV-2 may be protracted.^3^ Repurposing existing antivirals is attractive due to their relative safeness and potential anti-SARS-CoV-2 mechanisms.^4^ Neuraminidase inhibitors (i.e. oseltamivir, zanamivir) and HIV protease inhibitors (i.e. lopinavir-ritonavir) have been hypothesized to inhibit SARS-CoV-2 proteases involved in the degradation of polyproteins that control viral replication.^5^ Adamantanes (i.e. amantadine, rimantadine) are thought to disrupt lysosomal trafficking, thereby impeding the release of SARS-CoV-2 ribonucleic acid (RNA) into the cell,^6^ and by inhibiting conductance of the envelope (E) protein.^7^ Acyclovir, a nucleotide analog antiviral, was found as a candidate drug for COVID-19 by potentially counteracting gene expression changes observed after SARS-CoV-2 infection.^8^

Evaluating repurposed drugs during a pandemic comprises numerous challenges.^9^ Up to December 8, 2020 there were 413 registered studies to test antivirals for COVID-19, of which 400 were still active.^10^ Most trials in the World Health Organization (WHO) platform were for lopinavir/ritonavir (156), favipiravir (62), remdesivir (52), oseltamivir (19), and ribavirin (16).^11^ Other common antivirals are not being tested for COVID-19 but could be having widespread use in the community and hospitals since practice guidelines do not discourage/recommend most antivirals due to a lack of evidence,^12,13^ others advise against most,^14,15^ or recommend oseltamivir empirically during the influenza season^16^ and when coinfection exists.^17^

The WHO recommends early antibiotic therapy in patients with severe COVID-19, but not for mild-to-moderate disease.^14^ However, questions have arisen on the effectiveness of antibiotics due to its viral origin, whilst indiscriminate use of antibiotics could aggravate antimicrobial resistance.^18^

In this population-based study, we hypothesized that repurposed antivirals and antibiotics could be having widespread use in real-world settings. To determine their impact on mortality, we studied survival and all-cause mortality risk in patients with laboratory-confirmed COVID-19 in Mexico City receiving these drugs in both ambulatory and in-hospital settings.

## METHODS

### Study Design

We conducted a real-world multicenter retrospective cohort study in patients who received medical attention for suspected COVID-19 in any of the 688 registered and accredited COVID-19 medical units in Mexico City, to evaluate all-cause mortality (main outcome) in those receiving antivirals, antibiotics, both, or none (exposition groups).

We considered 395,343 patients for eligibility evaluated for COVID-19 in 688 medical units (primary-to-tertiary care) between February 24, 2020 and September 14, 2020. All patients with a positive RT-PCR for SARS-CoV-2 were included to maximize the power and generalizability. Patients treated with unspecified antivirals were excluded. To perform reliable analyses, a cut-off value of 30 patients receiving the same antiviral was set and groups of antivirals with <30 patients were eliminated.

### Source of Data

We used the COVID-19 open dataset available in Mexico City Government’s Open Data platform,^19^ collected and updated daily by the Secretariat of Health of Mexico City. Patients meeting criteria of suspected COVID-19 case have been included in this dataset starting on February 24, 2020 when the first suspected cases arrived in Mexico. Criteria for suspected COVID-19 case in Mexico included having at least two of three signs/symptoms (cough, fever, or headache) plus at least one other (dyspnea, arthralgias, myalgias, sore throat, rhinorrhea, conjunctivitis, or chest pain) in the last 7 days. This operational definition was changed on August 24, 2020 to increase sensitivity:^20^ at least one of four signs/symptoms (cough, fever, dyspnea, or headache), plus at least one other (myalgias, arthralgias, sore throat, chills, chest pain, rhinorrhea, anosmia, dysgeusia, or conjunctivitis) in the last 10 days.

For epidemiologic purposes, two strategies are outlined in the National COVID-19 Epidemiologic Surveillance Plan:^20^ 1. testing of 10% of ambulatory patients with mild symptoms of respiratory disease and 100% of patients with respiratory distress at evaluation in monitoring units of viral respiratory disease (USMER, for its acronym in Spanish), and 2. testing 100% of patients who meet diagnostic criteria of Severe Acute Respiratory Infection (defined as shortness of breath, temperature ≥38 °C, cough, and ≥1 of the following: chest pain, tachypnea, or acute respiratory distress syndrome) in non-USMER units.

Upon evaluating a patient suspected of having COVID-19, healthcare professionals are required to fill out a format (Supplementary Appendix) containing demographic, clinical, epidemiological, and treatment variables, later complemented with follow-up by accredited hospital epidemiologists (inpatients) and healthcare professionals in primary care units (ambulatory patients). For ambulatory patients, follow-up is performed daily for a minimum 7 days and patients are considered recovered 14 days after the onset of symptoms if alive and not hospitalized. For hospitalized patients, follow-up is done daily until death or discharge; follow-up time for patients discharged from hospital is highly variable since no consensus or requirements by authorities exist but may extend from 14 days to 3-6 months after discharge. Duration of follow-up for each patient is not provided in the dataset and cannot be calculated.

For every medical unit there is only one responsible authority who ultimately uploads data into the Respiratory Diseases Epidemiologic Surveillance System and is accountable for accuracy. Results of diagnostic RT-PCR for SARS-CoV-2 are directly uploaded by the diagnostic facility; accreditation of diagnostic procedures by the Mexican Institute of Diagnostics and Epidemiological Reference is required to upload results. Reporting of all deaths of COVID-19 suspected or confirmed cases is obligatory and must be done in the first 48 hours after occurrence; in cases of deaths occurring in patients who had completed follow-up, registries are matched to death certificates and updated. There have been concerns that patients tested more than once may be duplicated. Since no variables that could lead to identification of patients are released, we searched for patients with identical demographic variables and only one registry was kept.

To determine whether prescription of oseltamivir occurred for cases of COVID-19 or influenza during the pandemic period, dates of prescription of oseltamivir and dates of admission of patients with laboratory-confirmed COVID-19 were obtained from the previously mentioned COVID-19 dataset and grouped according to epidemiological weeks. Data of patients who had a positive test for influenza in Mexico City for every epidemiological week were obtained from the Directorate General of Epidemiology’s Weekly Surveillance Reports of Influenza.^21^

### Management of Variables

All categorical variables were classified as dummy variables (present/absent). Polytomous variables were created from frequencies of use of antivirals and antibiotics (no antiviral/antibiotic, antiviral only, antibiotic only, and antiviral plus antibiotic), type of antiviral with >30 patients, and the combination of every individual antiviral with antibiotics. These were considered as the exposition groups. Special populations for subgroup analyses were defined as: children and adolescents (<18 years), pregnancy, puerperium, and non-pregnant/puerperal adults (≥18 years). Further subgroups included ambulatory and hospitalized patients, as well as patients admitted to intensive care unit (ICU) and those requiring invasive mechanical ventilation (IMV). A variable of critical patients was built by grouping patients admitted to ICU and/or requiring IMV, whereas non-critical patients did not meet any of both.

Since it has been hypothesized that early use of antivirals for COVID-19 could diminish hospitalization rate^22^ and detain disease progression,^23^ thereby decreasing mortality, we distinguished early (≤2 days from symptom onset to initiation of antivirals) from late (>2 days) use of antivirals, and studied their relation to hospitalization rates and mortality; only patients who received antivirals before being evaluated in an accredited COVID-19 unit were included for this analysis since dates of initiation of antivirals are only collected for such patients. Therefore, we studied these patients as a different sub-cohort.

Occupations were grouped as follows: technical services (laborers), education (students and teachers), healthcare (dentists, nurses, diagnostic laboratorian, physicians, and other healthcare workers), agricultural activities (peasants), commerce (drivers, informal commerce, employees, and businesspeople), unemployed, stay-at-home (stay-at-home parents and retired/pensioners), and other occupations (others, and other professions). Variables for adjustment of models were: sex, age, indigenous self-identification, diabetes, COPD, immunosuppression, hypertension, HIV/AIDS, cardiovascular disease, obesity, CKD, smokers, unemployed, time from symptom onset to medical attention, fever, cough, sore throat, shortness of breath, irritability, diarrhea, chest pain, chills, headache, myalgias, arthralgias, abrupt deterioration, rhinorrhea, polypnea, vomit, abdominal pain, conjunctivitis, cyanosis, and sudden onset of symptoms.

### Statistical Analysis

Descriptive data were calculated and are provided as frequencies, percentages, mean with standard deviation (SD) or median with interquartile range (IQR). Qualitative comparisons were made with χ^2^ or Fisher’s exact test. Independent-samples t-test and ANOVA were used for quantitative comparisons. Survival was calculated for all treatment groups (antiviral only, antibiotic only, antiviral plus antibiotic, and no antiviral/antibiotic) and specific antivirals (acyclovir, amantadine, lopinavir-ritonavir, oseltamivir, rimantadine, and zanamivir) alone or combined with antibiotics; survival curves were created for general population, ambulatory, hospitalized, non-critical, and critical patients. Survival between groups receiving distinct treatments were compared through the Log-Rank test against patients not receiving antivirals/antibiotics. Cox regression models were applied for general population, ambulatory, hospitalized, non-critical, and critical patients to determine mortality risk in patients receiving any treatment compared to no antivirals/antibiotics (reference). Resulting hazard ratios (HR) were adjusted for demographic and clinical variables considered as risk factors in the univariate analysis for every group; all variables with p<0.1 were included in the final model using the Enter method.

To account for multicenter variability, adjusted risk was calculated through generalized estimating equations (GEE), setting the medical unit with the lowest CFR and the highest number of patients for every subgroup as the reference value. Further subgroup survival analyses and multivariable Cox regression models were applied for special populations (children and adolescents, pregnancy, puerperium, and non-pregnant/puerperal adults), invasive mechanical ventilation (IMV), and intensive care unit (ICU). To quantify the minimal association strength of an unmeasured confounding factor that could reduce the risk conferred by exposures in our study, E-values were calculated for the point estimate and lower limit of the confidence interval.

To reduce potential confounding and selection bias, we applied a propensity score analysis. Propensity scores were calculated with a logistic regression model adjusted for: sex, age, signs and symptoms (fever, dyspnea, arthralgias, myalgias), and comorbidities (hypertension, diabetes, chronic kidney disease, cardiovascular disease, obesity, and immunosuppression). A 1:1 pairing was performed through the nearest-neighbor algorithm matching. Density functions of treated patients against controls before and after matching were graphed to determine appropriate matching. Subsequently, multivariable Cox regression models were applied to determine mortality risk in patients treated with antivirals, antibiotics, and oseltamivir versus a control group. A two-sided p value <0.05 was used to define statistical significance. Analyses and figures were created with SPSS software v.21 and GraphPad Prism v.8.0.1.

## RESULTS

No duplicated registries were found. After selection of eligible participants (Figure 1), 136,855 patients from all 688 medical units were analyzed. 97.83% (n=133,887) were residents of the Mexico City Metropolitan Area, conformed by 17 municipalities of Mexico City (83.29%, n=111,768), and 60 municipalities (16.71%, n=22,119) of the State of Mexico. The remaining 2.17% (n=2,968) sought medical attention from all other 30 states of the republic.

**Figure 1.**
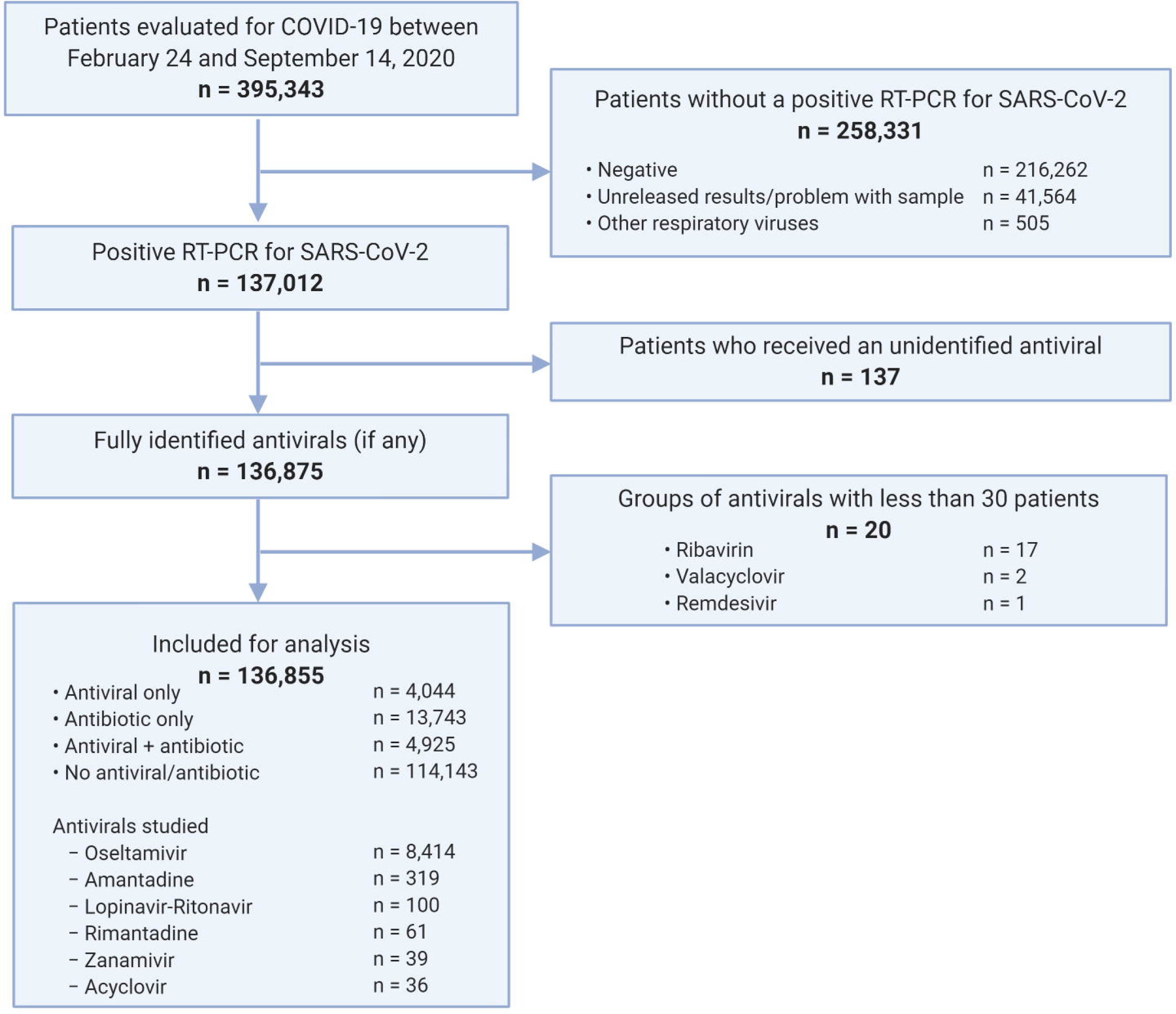
Flow diagram of patients assessed for eligibility.

Of all patients, 10.0% (n=13,743) received antibiotics only; 3.0% (n=4,044), antivirals only; 3.6% (n=4,925), antivirals plus antibiotics, and 83.4% (n=114,143), none (Table 1). More symptomatic ambulatory patients received antivirals and antibiotics more frequently (Supplementary Table 1); hospitalized patients with more signs/symptoms had greater use of antivirals, but less antibiotics (Supplementary Table 2).

**Table 1.** Baseline characteristics of patients with laboratory confirmed COVID-19 who were treated with or without antivirals/antibiotics, in 688 accredited COVID-19 medical units in Mexico City.

|  | All patients<br>n=136855 | No antiviral /<br>antibiotic<br>n=114143 | Antiviral<br>only<br>n=4044 | Acyclovir<br>n=36 | Amantadine<br>n=319 | Lopinavir-Ritonavir<br>n=100 | Oseltamivir<br>n=8414 | Rimantadine<br>n=61 | Zanamivir<br>n=39 | Antibiotic only<br>n=13743 |
| --- | --- | --- | --- | --- | --- | --- | --- | --- | --- | --- |
| Sex |  |  |  |  |  |  |  |  |  |  |
| Women | 66683 (48.7) | 56999 (49.9) | 1813 (44.8) | 19 (52.8) | 182 (57.1) | 31 (31) | 3407 (40.5) | 28 (45.9) | 17 (43.6) | 6000 (43.7) |
| Men | 70172 (51.3) | 57144 (50.1) | 2231 (55.2) | 17 (47.2) | 137 (42.9) | 69 (69) | 5007 (59.5) | 33 (54.1) | 22 (56.4) | 7743 (56.3) |
| Age, mean (SD) | 44.2 (16.8) | 43.1 (16.6) | 50.5 (16.5) | 46.9 (14.9) | 43.9 (14.8) | 56.9 (15.9) | 51.8 (15.9) | 46 (15.1) | 50 (14.1) | 48.3 (16.8) |
| Age categories |  |  |  |  |  |  |  |  |  |  |
| 0-19 years | 7558 (5.5) | 6963 (6.1) | 57 (1.4) | 0 (0) | 12 (3.8) | 1 (1) | 97 (1.2) | 2 (3.3) | 0 (0) | 483 (3.5) |
| 20-29 years | 20098 (14.7) | 18027 (15.8) | 375 (9.3) | 6 (16.7) | 38 (11.9) | 2 (2) | 638 (7.6) | 5 (8.2) | 3 (7.7) | 1379 (10) |
| 30-39 years | 29434 (21.5) | 25586 (22.4) | 707 (17.5) | 6 (16.7) | 86 (27.0) | 10 (10) | 1286 (15.3) | 16 (26.2) | 7 (17.9) | 2437 (17.7) |
| 40-49 years | 29553 (21.6) | 24683 (21.6) | 837 (20.7) | 10 (27.8) | 71 (22.3) | 21 (21) | 1780 (21.2) | 14 (23) | 8 (20.5) | 2966 (21.6) |
| 50-59 years | 24928 (18.2) | 20011 (17.5) | 852 (21.1) | 5 (13.9) | 63 (19.7) | 23 (23) | 1895 (22.5) | 15 (24.6) | 10 (25.6) | 2906 (21.1) |
| 60-69 years | 15070 (11.0) | 11441 (10.0) | 632 (15.6) | 7 (19.4) | 32 (10) | 18 (18) | 1515 (18.0) | 3 (4.9) | 6 (15.4) | 2048 (14.9) |
| 70-79 years | 7183 (5.2) | 5213 (4.6) | 418 (10.3) | 2 (5.6) | 14 (4.4) | 17 (17) | 855 (10.2) | 5 (8.2) | 5 (12.8) | 1072 (7.8) |
| 80-89 years | 2594 (1.9) | 1902 (1.7) | 146 (3.6) | 0 (0) | 2 (0.6) | 8 (8) | 292 (3.5) | 1 (1.6) | 0 (0) | 389 (2.8) |
| 90-99 years | 419 (0.3) | 303 (0.3) | 20 (0.5) | 0 (0) | 1 (0.3) | 0 (0) | 56 (0.7) | 0 (0) | 0 (0) | 59 (0.4) |
| ≥100 years | 18 (0.01) | 14 (0.01) | 0 (0) | 0 (0) | 0 (0) | 0 (0) | 0 (0) | 0 (0) | 0 (0) | 4 (0.03) |
| Indigenous self-identification | 713 (0.5) | 537 (0.5) | 37 (0.9) | 0 (0) | 2 (0.6) | 1 (1) | 79 (0.9) | 0 (0) | 2 (5.1) | 92 (0.7) |
| Occupation |  |  |  |  |  |  |  |  |  |  |
| Technical services | 1916 (1.4) | 1527 (1.3) | 70 (1.7) | 0 (0) | 3 (0.9) | 3 (3) | 145 (1.7) | 0 (0) | 1 (2.6) | 237 (1.7) |
| Education | 10006 (7.3) | 9129 (8) | 105 (2.6) | 1 (2.8) | 24 (7.5) | 1 (1) | 187 (2.2) | 2 (3.3) | 0 (0) | 662 (4.8) |
| Healthcare | 17281 (12.6) | 14910 (13.1) | 655 (16.2) | 3 (8.3) | 47 (14.7) | 9 (9) | 1029 (12.2) | 7 (11.5) | 2 (5.1) | 1274 (9.3) |
| Agricultural activities | 302 (0.2) | 232 (0.2) | 5 (0.1) | 0 (0) | 0 (0) | 0 (0) | 19 (0.2) | 0 (0) | 0 (0) | 51 (0.4) |
| Commerce | 50450 (36.9) | 42625 (37.3) | 1078 (26.7) | 15 (41.7) | 111 (34.8) | 36 (36) | 2569 (30.5) | 27 (44.3) | 12 (30.8) | 5055 (36.8) |
| Other | 24630 (18) | 19906 (17.4) | 911 (22.5) | 6 (16.7) | 63 (19.7) | 20 (20) | 2021 (24) | 9 (14.8) | 6 (15.4) | 2599 (18.9) |
| Unemployed | 5685 (4.2) | 4747 (4.2) | 277 (6.8) | 1 (2.8) | 5 (1.6) | 2 (2) | 463 (5.5) | 3 (4.9) | 7 (17.9) | 457 (3.3) |
| Stay-at-home | 26585 (19.4) | 21067 (18.5) | 943 (23.3) | 10 (27.8) | 66 (20.7) | 29 (29) | 1981 (23.5) | 13 (21.3) | 11 (28.2) | 3408 (24.8) |
| Last-season flu vaccination | 27087 (19.8) | 22972 (20.1) | 695 (17.2) | 9 (25) | 85 (26.6) | 9 (9) | 1244 (14.8) | 13 (21.3) | 4 (10.3) | 2751 (20) |
| Special populations |  |  |  |  |  |  |  |  |  |  |
| Pregnancy | 583 (0.9) | 530 (0.9) | 12 (0.3) | 0 (0) | 2 (1.1) | 0 (0) | 16 (0.5) | 0 (0) | 0 (0) | 35 (0.6) |
| Age during pregnancy, mean (SD) | 29.8 (7.4) | 29.5 (6.9) | 30.3 (5.2) | - | 32 (4.2) | - | 30 (5.9) | - | - | 34.8 (12.4) |
| Last-season flu vaccination | 161 (27.6) | 153 (28.9) | 1 (8.3) | 0 (0) | 0 (0) | 0 (0) | 1 (6.3) | 0 (0) | 0 (0) | 7 (20) |
| Pregnancy age group |  |  |  |  |  |  |  |  |  |  |
| Early adolescent (≤14 years) | 2 (0.3) | 2 (0.4) | 0 (0) | 0 (0) | 0 (0) | 0 (0) | 0 (0) | 0 (0) | 0 (0) | 0 (0) |
| Late adolescent (15-19 years) | 34 (5.8) | 33 (6.2) | 1 (8.3) | 0 (0) | 0 (0) | 0 (0) | 1 (6.3) | 0 (0) | 0 (0) | 0 (0) |
| Normal age (20-34 years) | 404 (69.3) | 373 (70.4) | 9 (75) | 0 (0) | 1 (50) | 0 (0) | 12 (75) | 0 (0) | 0 (0) | 18 (51.4) |
| Advanced maternal age (≥35 years) | 143 (24.5) | 122 (23) | 2 (16.7) | 0 (0) | 1 (50) | 0 (0) | 3 (18.8) | 0 (0) | 0 (0) | 17 (48.6) |
| Trimester of pregnancy |  |  |  |  |  |  |  |  |  |  |
| First trimester | 114 (19.6) | 102 (19.2) | 5 (41.7) | 0 (0) | 1 (50) | 0 (0) | 5 (31.3) | 0 (0) | 0 (0) | 6 (17.1) |
| Second trimester | 177 (30.4) | 161 (30.4) | 3 (25) | 0 (0) | 0 (0) | 0 (0) | 4 (25) | 0 (0) | 0 (0) | 12 (34.3) |
| Third trimester | 292 (50.1) | 267 (50.4) | 4 (33.3) | 0 (0) | 1 (50) | 0 (0) | 7 (43.8) | 0 (0) | 0 (0) | 17 (48.6) |
| Puerperium | 103 (0.2) | 64 (0.1) | 2 (0.05) | 0 (0) | 0 (0.0) | 0 (0) | 7 (0.2) | 0 (0) | 0 (0) | 32 (0.5) |
| Days of puerperium |  |  |  |  |  |  |  |  |  |  |
| 1 day | 33 (32) | 21 (32.8) | 1 (50) | 0 (0) | 0 (0) | 0 (0) | 3 (42.9) | 0 (0) | 0 (0) | 9 (28.1) |
| 2-7 days | 33 (32) | 16 (25) | 1 (50) | 0 (0) | 0 (0) | 0 (0) | 3 (42.9) | 0 (0) | 0 (0) | 14 (43.8) |
| 8-42 days | 37 (35.9) | 27 (42.2) | 0 (0) | 0 (0) | 0 (0) | 0 (0) | 1 (14.3) | 0 (0) | 0 (0) | 9 (28.1) |
| Age during puerperium, mean (SD) | 31.9 (9.7) | 31.1 (9.2) | 33.5 (3.5) | - | - | - | 31.1 (2.7) | - | - | 33.6 (11.5) |
| Last-season flu vaccination | 22 (21.4) | 14 (21.9) | 14 (21.9) | 0 (0) | 0 (0) | 0 (0) | 1 (14.3) | 0 (0) | 0 (0) | 7 (21.9) |
| Children and adolescents (<18 years) | 5791 (4.2) | 5336 (4.7) | 40 (1) | 0 (0) | 9 (2.8) | 1 (1) | 67 (0.8) | 2 (3.3) | 0 (0) | 376 (2.7) |
| Age, mean (SD) | 10.9 (5.2) | 10.9 (5.2) | 10.9 (5.9) | - | 13.4 (4.5) | - | 11 (5.8) | 14.5 (3.6) | - | 9.9 (5.9) |
| Last-season flu vaccination | 1213 (20.9) | 1113 (20.9) | 4 (10) | 0 (0) | 1 (11.1) | 0 (0) | 9 (13.4) | 0 (0) | 0 (0) | 90 (23.9) |
| Non-pregnant/puerperal adults (≥18 years) | 130378 (95.3) | 108213 (94.8) | 3990 (98.7) | 36 (100) | 308 (96.6) | 99 (9) | 8324 (98.9) | 59 (96.7) | 39 (100) | 13300 (96.8) |
| Age, mean (SD) | 45.7 (15.5) | 44.7 (15.3) | 50.9 (16.1) | 46.9 (14.9) | 44.8 (14) | 57.5 (15.1) | 52.1 (15.6) | 47.1 (14.1) | 50 (14.1) | 48.5 (15.7) |
| Last-season flu vaccination | 25691 (19.7) | 21692 (20) | 690 (17.3) | 9 (25) | 84 (27.3) | 9 (9.1) | 1233 (14.8) | 13 (22) | 4 (10.3) | 2647 (19.9) |
| Comorbidities |  |  |  |  |  |  |  |  |  |  |
| Diabetes | 18229 (13.3) | 13458 (11.8) | 910 (22.5) | 2 (5.6) | 36 (11.3) | 35 (35) | 2007 (23.9) | 8 (13.1) | 6 (15.4) | 2677 (19.5) |
| COPD | 1741 (1.3) | 1273 (1.1) | 119 (2.9) | 0 (0) | 3 (0.9) | 2 (2) | 212 (2.5) | 2(3.3) | 1 (2.6) | 248 (1.8) |
| Asthma | 3035 (2.2) | 2561 (2.2) | 96 (2.4) | 1 (2.8) | 13 (4.1) | 4 (4) | 165 (2) | 0 (0) | 3 (7.7) | 288 (2.1) |
| Immunosuppression | 1758 (1.3) | 1368 (1.2) | 84 (2.1) | 0 (0) | 7 (2.2) | 3 (3) | 145 (1.7) | 0 (0) | 0 (0) | 235 (1.7) |
| Hypertension | 22185 (16.2) | 16799 (14.7) | 1074 (26.6) | 10 (27.8) | 40 (12.5) | 35 (35) | 2211 (26.3) | 12 (19.7) | 5 (12.8) | 3073 (22.4) |
| HIV/AIDS | 573 (0.4) | 462 (0.4) | 168 (4.2) | 0 (0) | 0 (0) | 2 (2) | 47 (0.6) | 0 (0) | 0 (0) | 62 (0.5) |
| Cardiovascular disease | 2724 (2.0) | 2064 (1.8) | 153 (3.8) | 2 (5.6) | 4 (1.3) | 5 (5) | 277 (3.3) | 2 (3.3) | 0(0) | 370 (2.7) |
| Obesity | 23848 (17.4) | 18924 (16.6) | 837 (20.7) | 12 (33.3) | 83 (26) | 18 (18) | 1817 (21.6) | 5 (8.2) | 6 (15.4) | 2983 (21.7) |
| Chronic kidney disease | 2067(1.5) | 1471 (1.3) | 150 (3.7) | 0 (0) | 2 (0.6) | 7 (7) | 286 (3.4) | 1 (1.6) | 0 (0) | 300 (2.2) |
| Smoker | 14727 (10.8) | 12214 (10.7) | 461 (11.4) | 2 (5.6) | 46 (14.4) | 6 (6) | 885 (10.5) | 8 (13.1) | 5 (12.8) | 1561 (11.4) |
| Type of medical attention |  |  |  |  |  |  |  |  |  |  |
| Ambulatory | 109902 (80.3) | 98060 (85.9) | 2012 (49.8) | 30 (83.3) | 282 (88.4) | 8 (8) | 3187 (37.9) | 48 (78.7) | 19 (48.7) | 8268 (60.2) |
| Hospitalization | 26953 (19.7) | 16083 (14.1) | 2032 (50.2) | 6 (16.7) | 37 (11.6) | 92 (92) | 5227 (62.1) | 13 (21.3) | 20 (51.3) | 5475 (39.8) |
| Severity of the disease |  |  |  |  |  |  |  |  |  |  |
| Non-critical | 129658 (94.7) | 110009 (96.4) | 3518 (87) | 34 (94.4) | 310 (97.2) | 69 (69) | 7126 (84.7) | 58 (95.1) | 34 (87.2) | 12018 (87.4) |
| Critical | 7197 (5.3) | 4134 (3.6) | 526 (13) | 2 (5.6) | 9 (2.8) | 31 (31) | 1288 (15.3) | 3 (4.9) | 5 (12.8) | 1725 (12.6) |
| Time from symptom onset to medical attention | 4.5 (3.8) | 4.4 (3.8) | 4.49 (3.9) | 6.36 (4.6) | 4.9 (3.6) | 6.8 (4.1) | 4.7 (3.7) | 5.6 (4.2) | 5.33 (4.7) | 5.5 (3.8) |
| Baseline symptoms |  |  |  |  |  |  |  |  |  |  |
| Fever | 83120 (60.7) | 66011 (57.8) | 3332 (82.4) | 25 (69.4) | 198 (62.1) | 81 (81) | 6963 (82.8) | 42 (68.9) | 31 (79.5) | 9769 (71.1) |
| Cough | 96206 (70.3) | 78367 (68.7) | 3406 (84.2) | 24 (66.7) | 245 (76.8) | 67 (67) | 7055 (83.8) | 42 (68.9) | 35 (89.7) | 10371 (75.5) |
| Sore throat | 59040 (43.1) | 48845 (42.8) | 2014 (49.8) | 12 (33.3) | 159 (49.8) | 31 (31) | 3701 (44) | 29 (47.5) | 22 (56.4) | 6241 (45.4) |
| Shortness of breath | 42942 (31.4) | 31061 (27.2) | 2234 (55.2) | 14 (38.9) | 85 (26.6) | 66 (66) | 5170 (61.4) | 22 (36.1) | 23 (59) | 6501 (47.3) |
| Irritability | 24098 (17.6) | 19460 (17) | 1079 (26.7) | 5 (13.9) | 81(25.4) | 7 (7) | 1993 (23.7) | 14 (23) | 13 (33.3) | 2525 (18.4) |
| Diarrhea | 31649 (23.1) | 25821 (22.6) | 1152 (28.5) | 11 (30.6) | 96 (30.1) | 19 (19) | 2180 (25.9) | 15 (24.6) | 13 (33.3) | 3494 (25.4) |
| Chest pain | 36851 (26.9) | 29524 (25.9) | 1662 (41.1) | 12 (33.3) | 105 (32.9) | 22 (22) | 2979 (35.4) | 25 (41) | 15 (38.5) | 4169 (30.3) |
| Chills | 48282 (35.3) | 39405 (34.5) | 2138 (52.9) | 16 (44.4) | 158 (49.5) | 34 (34) | 3616 (43) | 33 (54.1) | 17 (43.6) | 5003 (36.4) |
| Headache | 95018 (69.4) | 78893 (69.1) | 3284 (81.2) | 22 (61.1) | 227 (71.2) | 59 (59) | 6348 (75.4) | 40 (65.6) | 29 (74.4) | 9400 (68.4) |
| Myalgias | 70666 (51.6) | 57192 (50.1) | 2633 (65.1) | 27 (75) | 201 (63.0) | 60 (60) | 5074 (60.3) | 43 (70.5) | 24 (61.5) | 8045 (58.5) |
| Arthralgias | 64381 (47) | 51792 (45.4) | 2377 (58.8) | 22 (61.1) | 179 (56.1) | 48 (48) | 4846 (57.6) | 42(68.9) | 23 (59) | 7429 (54.1) |
| Abrupt deterioration | 62460 (45.6) | 48991 (42.9) | 2704 (66.9) | 20 (55.6) | 180 (56.4) | 66 (66) | 5367 (63.8) | 34 (55.7) | 20 (51.3) | 7782 (56.6) |
| Rhinorrhea | 38288 (28) | 32135 (28.2) | 1350 (33.4) | 6 (16.7) | 118 (37.0) | 20 (20) | 2283 (27.1) | 27 (44.3) | 11 (28.2) | 3688 (26.8) |
| Polypnea | 15868 (11.6) | 11977 (10.5) | 1081 (26.7) | 3 (8.3) | 46 (14.4) | 13 (13) | 1847 (22) | 7 (11.5) | 6 (15.4) | 1969 (14.3) |
| Vomit | 10123 (7.4) | 8094 (7.1) | 479 (11.8) | 3 (8.3) | 23 (7.2) | 7 (7) | 870 (10.3) | 8 (13.1) | 2 (5.1) | 1116 (8.1) |
| Abdominal pain | 17338 (12.7) | 14080 (12.3) | 1025 (25.3) | 3 (8.3) | 60 (18.8) | 8 (8) | 1561 (18.6) | 10 (16.4) | 7 (17.9) | 1609 (11.7) |
| Conjunctivitis | 16941 (12.4) | 14277 (12.5) | 513 (12.7) | 4 (11.1) | 55 (17.2) | 7 (7) | 962 (11.4) | 11 (18) | 6 (15.4) | 1619 (11.8) |
| Cyanosis | 5917 (4.3) | 4461 (3.9) | 463 (11.4) | 1 (2.8) | 12 (3.8) | 4 (4) | 816 (9.7) | 8 (13.1) | 3 (7.7) | 612 (4.5) |
| Sudden onset of symptoms | 46723 (34.1) | 37607 (32.9) | 1574 (38.9) | 10 (27.8) | 96 (30.1) | 48 (48) | 3831 (45.5) | 26 (42.6) | 16 (41) | 5089 (37) |
| Concomitant use of antibiotics | 18840 (13.8) | 172 (0.2) | - | 29 (80.6) | 151 (47.3) | 77 (77) | 4627 (55) | 26 (42.6) | 15 (38.5) | - |
| Data expressed as Frequency (%) or mean (SD) |  |  |  |  |  |  |  |  |  |  |
| SD: Standard deviation, COPD: Chronic obstructive pulmonary disease, HIV/AIDS: Human immunodeficiency virus/acquired immune deficiency syndrome. |  |  |  |  |  |  |  |  |  |  |

Baseline and follow-up characteristics of survivors (91.47%, n=136,855) and non-survivors (8.53%, n=11,679) are shown in Supplementary Table 3. Case-fatality rates (CFR) in special populations were: 8.92% (95%CI:8.76-9.07%), for non-pregnant/puerperal adults; 1.72% (95%CI:0.66-2.77), pregnancy; 0.97% (95%CI:0-2.90), puerperium; and 0.69% (95%CI:0.48-0.90), children and adolescents. Of all deaths, 92.7% (95%CI:92.2-93.2) and 99.6% (95%CI:99.5-99.7) occurred by day 28 and 56, respectively.

Patients treated only with antivirals had lower survival than those not receiving antivirals/antibiotics in the general population (Figure 2a), ambulatory (Figure 2b), hospitalized (Figure 2c), non-critical (Figure 3a), critical (Figure 3c), IMV (Supplementary Table 4), ICU (Supplementary Table 5) and non-pregnant/puerperal adults (Supplementary Table 6); for children and adolescents (Supplementary Table 7) and pregnancy (Supplementary Table 8) differences in survival were not significant; there were not enough events for analysis in puerperal women. Increased survival with only antibiotics was observed in hospitalized, critical, and IMV, whereas decreased survival occurred in the general population, non-pregnant/puerperal adults, ambulatory, non-critical, ICU, and children and adolescents; there were no differences for pregnancy. Antivirals plus antibiotics resulted in decreased survival in the general population, ambulatory, non-critical, non-pregnant/puerperal adults, children and adolescents, pregnancy, and ICU; increased survival, in hospitalized; and no differences, in critical and IMV groups.

**Figure 2.**
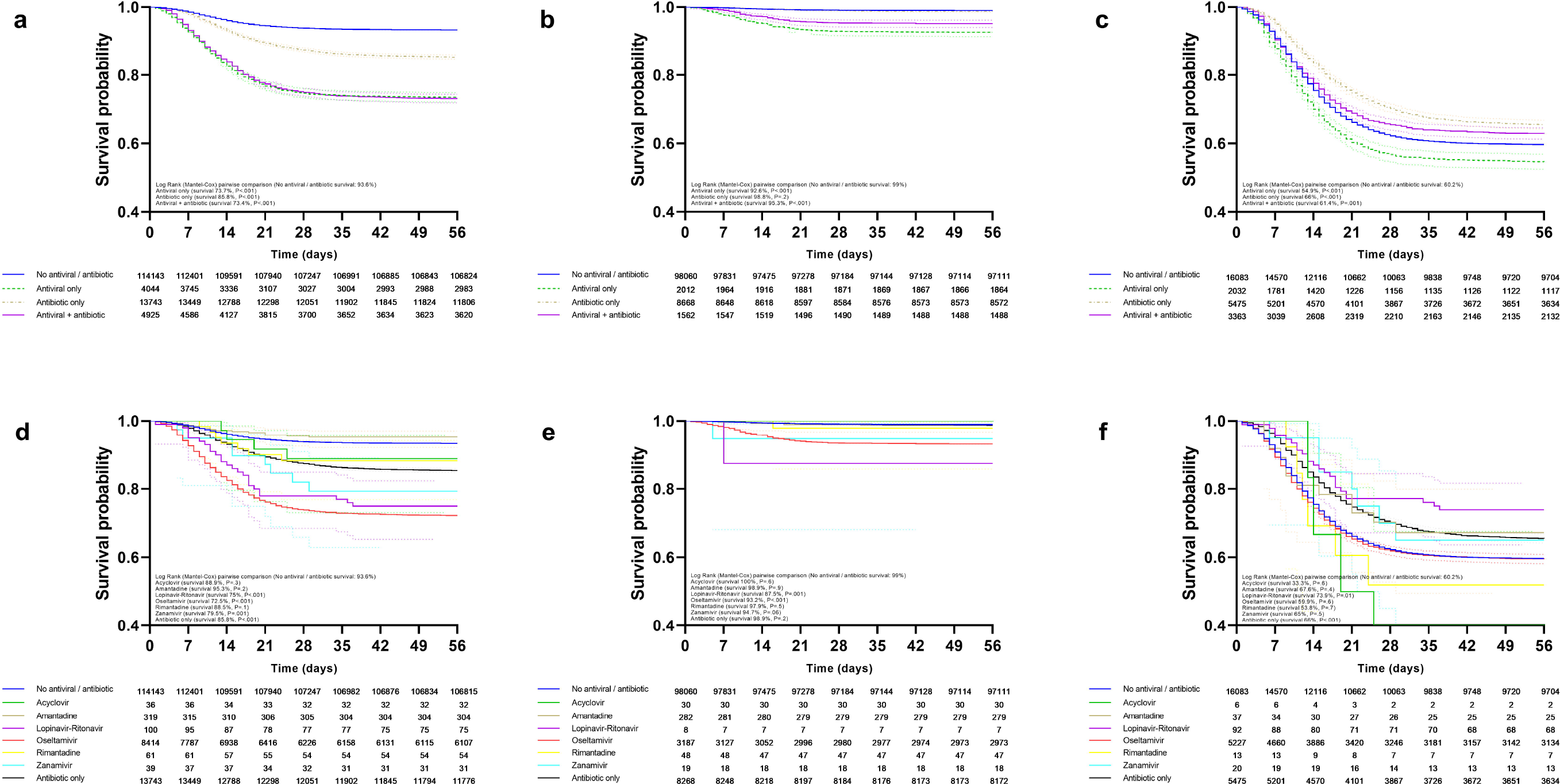
Survival of patients (general population, ambulatory, and hospitalized) treated with antivirals and/or antibiotics. Survival curves are shown according to treatment modality in the general population (a), ambulatory (b), and hospitalized (c) patients. Survival in patients receiving specific antivirals, antibiotics, both, or none in the general population (d), ambulatory (e), and hospitalization (f) settings.

**Figure 3.**
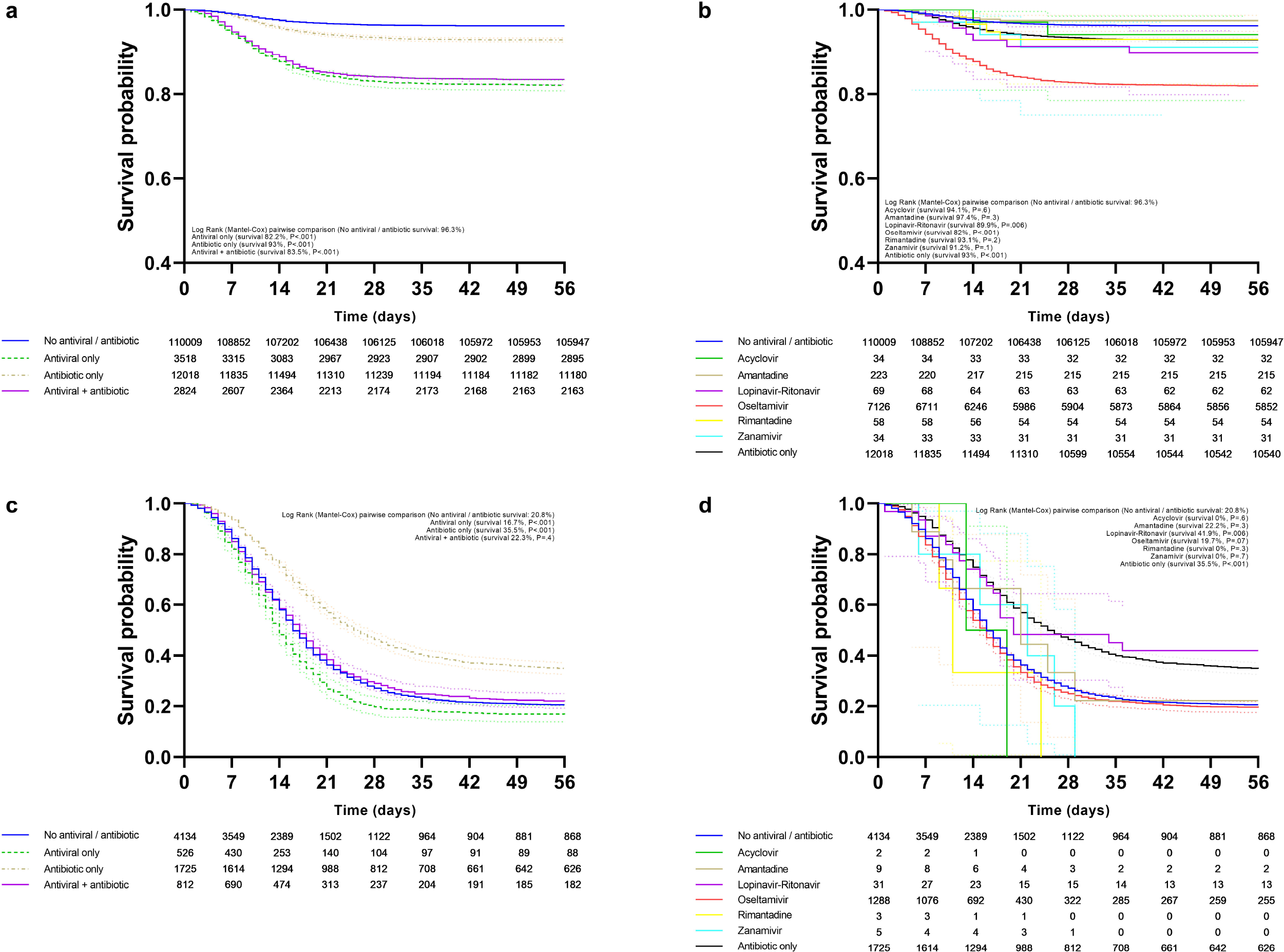
Survival of patients (non-critical and critical) treated with antivirals and/or antibiotics. Survival curves are shown according to treatment modality in non-critical (a) and critical (b) patients. Survival in critical (c) and non-critical (d) patients receiving specific antivirals, antibiotics, both, or none.

Decreased survival with oseltamivir was observed in the general population (Figure 2d), ambulatory (Figure 2e), non-critical (Figure 2d), ICU (Supplementary Table 5), non-pregnant/puerperal adults (Supplementary Table 6), children and adolescents (Supplementary Table 7), and pregnancy (Supplementary Table 8); no differences in survival occurred in hospitalized (Figure 2f), critical (Figure 3d), and IMV (Supplementary Table 4). Survival rates for amantadine, zanamivir, rimantadine, acyclovir, and lopinavir-ritonavir are shown in the same figures and tables as oseltamivir.

Unadjusted (Supplementary Table 9) and adjusted (Table 2) risk of death for the general population, ambulatory, hospitalized, non-critical and critical patients, as well as for other subgroups (Supplementary Tables 10-14) were calculated. E-values for statistically significant risk groups are provided in Supplementary Tables 15-16. After adjusting for center through GEE, we found no statistically significant variability in mortality risk for antivirals, antibiotics, or both in all groups. Oseltamivir presented variability in hospitalized and critical patients, with the largest increases in risk occurring in public hospitals. However, center was not a modifying risk factor after logistic regression analysis in hospitalized and critical patients.

**Table 2.** Adjusted mortality risk in laboratory-confirmed COVID-19 patients receiving antivirals, antibiotics, both, or none in 688 accredited COVID-19 medical units in Mexico City.

|  | All patients<sup>a</sup> |  | Ambulatory<sup>b</sup> |  | Hospitalized<sup>c</sup> |  | Non-Critical<sup>d</sup> |  | Critical<sup>e</sup> |  |
| --- | --- | --- | --- | --- | --- | --- | --- | --- | --- | --- |
|  | HR (95%CI) | p | HR (95%CI) | p | HR (95%CI) | p | HR (95%CI) | p | HR (95%CI) | p |
| Models for type of treatment |  |  |  |  |  |  |  |  |  |  |
| No antiviral / antibiotic | Reference |  | Reference |  | Reference |  | Reference |  | Reference |  |
| Antiviral only | 1.72 (1.61-1.84) | <0.0001 | 4.7 (3.94-5.62) | <0.0001 | 1.07 (0.99-1.15) | 0.07 | 2.03 (1.86-2.21) | <0.0001 | 1.09 (0.99-1.21) | 0.09 |
| Antibiotic only | 1.13 (1.08-1.19) | <0.0001 | 0.72 (0.58-0.89) | 0.003 | 0.81 (0.77-0.86) | <0.0001 | 1.05 (0.98-1.14) | 0.2 | 0.67 (0.63-0.72) | <0.0001 |
| Antiviral + antibiotic | 1.57 (1.47-1.67) | <0.0001 | 1.91 (1.47-2.49) | <0.0001 | 0.91 (0.86-0.97) | 0.004 | 1.63 (1.49-1.77) | <0.0001 | 1.02 (0.93-1.11) | 0.7 |
| Models for type of antiviral |  |  |  |  |  |  |  |  |  |  |
| No antiviral / antibiotic | Reference |  | Reference |  | Reference |  | Reference |  | Reference |  |
| Acyclovir | 1.37 (0.51-3.65) | 0.5 | Not estimable |  | 2.75 (1.03-7.33) | 0.04 | 1.19 (0.29-4.75) | 0.8 | 2.85 (0.71-11.4) | 0.1 |
| Amantadine | 0.73 (0.44-1.21) | 0.2 | 0.08 (0.24-2.36) | 0.6 | 0.88 (0.5-1.55) | 0.7 | 0.67 (0.33-1.34) | 0.3 | 1.05 (0.49-2.21) | 0.9 |
| Lopinavir-Ritonavir | 1.04 (0.69-1.55) | 0.9 | 4.28 (0.59-30.7) | 0.1 | 0.59 (0.4-0.89) | 0.01 | 0.69 (0.33-1.46) | 0.3 | 0.66 (0.41-1.04) | 0.08 |
| Oseltamivir | 1.66 (1.58-1.75) | <0.0001 | 3.52 (3.01-4.11) | <0.0001 | 0.98 (0.93-1.03) | 0.4 | 1.84 (1.72-1.96) | <0.0001 | 1.06 (0.99-1.14) | 0.1 |
| Rimantadine | 1.39 (0.66-2.92) | 0.4 | 2.54 (0.36-18.1) | 0.4 | 1.11 (0.49-2.46) | 0.8 | 1.48 (0.56-3.95) | 0.4 | 1.63 (0.52-5.09) | 0.4 |
| Zanamivir | 1.66 (0.83-3.32) | 0.2 | 2.49 (0.35-17.8) | 0.4 | 0.84 (0.39-1.76) | 0.6 | 1.43 (0.46-4.43) | 0.5 | 0.7 (0.29-1.69) | 0.4 |
| Antibiotic only | 1.14 (1.08-1.19) | <0.0001 | 0.72 (0.58-0.9) | 0.004 | 0.81 (0.77-0.86) | <0.0001 | 1.06 (0.98-1.14) | 0.2 | 0.68 (0.63-0.72) | <0.0001 |
| Models for Acyclovir |  |  |  |  |  |  |  |  |  |  |
| No antiviral / antibiotic | Reference |  | Reference |  | Reference |  | Reference |  | Reference |  |
| Acyclovir only | 8.1 (1.14-57.6) | 0.04 | Not estimable |  | 8.98 (1.26-63.9) | 0.03 | Not estimable |  | 2.85 (0.39-20.3) | 0.3 |
| Antibiotic only | 1.11 (1.05-1.17) | <0.0001 | 0.71 (0.57-0.89) | 0.002 | 0.82 (0.77-0.86) | <0.0001 | 1.03 (0.96-1.11) | 0.4 | 0.67 (0.63-0.72) | <0.0001 |
| Acyclovir + antibiotic | 1.07 (0.35-3.33) | 0.9 | Not estimable |  | 2.28 (0.74-7.08) | 0.2 | 1.23 (0.31-4.92) | 0.8 | 3.11 (0.44-22.2) | 0.3 |
| Models for Amantadine |  |  |  |  |  |  |  |  |  |  |
| No antiviral / antibiotic | Reference |  | Reference |  | Reference |  | Reference |  | Reference |  |
| Amantadine only | 1.78 (1.03-3.06) | 0.04 | 1.69 (0.42-6.79) | 0.5 | 1.62 (0.89-2.93) | 0.1 | 1.63 (0.78-3.42) | 0.2 | 1.39 (0.62-3.1) | 0.4 |
| Antibiotic only | 1.11 (1.05-1.17) | <0.0001 | 0.71 (0.57-0.89) | .002 | 0.82 (0.77-0.86) | <0.0001 | 1.03 (0.96-1.12) | 0.4 | 0.67 (0.63-0.72) | <0.0001 |
| Amantadine + antibiotic | 0.15 (0.04-0.59) | 0.007 | 0.34 (0.05-2.39) | 0.3 | 0.15 (0.02-1.06) | 0.06 | 0.13 (0.02-0.9) | 0.04 | 0.44 (0.06-3.11) | 0.4 |
| Models for Lopinavir-Ritonavir |  |  |  |  |  |  |  |  |  |  |

| No antiviral /<br>antibiotic | Reference |  | Reference |  | Reference |  | Reference |  | Reference |  |
| --- | --- | --- | --- | --- | --- | --- | --- | --- | --- | --- |
| Lopinavir-Ritonavir<br>only | 0.68 (0.26-1.82) | 0.4 | 56.9 (7.87-412) | <0.0001 | 0.39 (0.15-1.05) | 0.06 | 0.47 (0.07-3.37) | 0.5 | 0.41 (0.15-1.08) | 0.07 |
| Antibiotic only | 1.11 (1.05-1.17) | <0.0001 | 0.71 (0.57-0.89) | 0.002 | 0.82 (0.77-0.86) | <0.0001 | 1.04 (0.96-1.12) | 0.4 | 0.67 (0.63-0.72) | <0.0001 |
| Lopinavir-Ritonavir<br>+ antibiotic | 1.1 (0.71-1.7) | 0.7 | Not estimable |  | 0.67 (0.43-1.05) | 0.08 | 0.69 (0.31-1.53) | 0.4 | 0.79 (0.47-1.34) | 0.4 |
| Models for Oseltamivir |  |  |  |  |  |  |  |  |  |  |
| No antiviral /<br>antibiotic | Reference |  | Reference |  | Reference |  | Reference |  | Reference |  |
| Oseltamivir only | 1.72 (1.61-1.84) | <0.0001 | 4.79 (4.01-5.75) | <0.0001 | 1.07 (0.99-1.15) | 0.06 | 2.05 (1.88-2.23) | <0.0001 | 1.11 (1.0-1.23) | 0.05 |
| Antibiotic only | 1.13 (1.08-1.19) | <0.0001 | 0.72 (0.58-0.89) | 0.003 | 0.81 (0.77-0.86) | <0.0001 | 1.06 (0.98-1.14) | 0.2 | 0.67 (0.63-0.72) | <0.0001 |
| Oseltamivir +<br>antibiotic | 1.61 (1.51-1.71) | <0.0001 | 2.1 (1.65-2.8) | <0.0001 | 0.92 (0.87-0.98) | 0.01 | 1.68 (1.55-1.83) | <0.0001 | 1.02 (0.94-1.12) | 0.6 |
| Models for Rimantadine |  |  |  |  |  |  |  |  |  |  |
| No antiviral /<br>antibiotic | Reference |  | Reference |  | Reference |  | Reference |  | Reference |  |
| Rimantadine only | 1.88 (0.85-4.21) | 0.1 | 4.9 (0.69-34.9) | 0.1 | 1.21 (0.5-2.91) | 0.7 | 1.81 (0.58-5.62) | 0.3 | 1.69 (0.54-5.27) | 0.4 |
| Antibiotic only | 1.11 (1.05-1.17) | <0.0001 | 0.71 (0.57-0.89) | <0.0001 | 0.82 (0.77-0.86) | <0.0001 | 1.03 (0.96-1.12) | 0.4 | 0.67 (0.63-0.72) | <0.0001 |
| Rimantadine +<br>antibiotic | 0.51 (0.07-3.6) | 0.5 | Not estimable |  | 0.77 (0.11-5.45) | 0.8 | 0.88 (0.12-6.23) | 0.9 | - | - |
| Models for Zanamivir |  |  |  |  |  |  |  |  |  |  |
| No antiviral /<br>antibiotic | Reference |  | Reference |  | Reference |  | Reference |  | Reference |  |
| Zanamivir only | 1.9 (0.85-4.25) | 0.12 | 3.99 (0.55-28.9) | 0.2 | 0.9 (0.37-2.17) | 0.8 | 1.2 (0.17-8.49) | 0.9 | 0.72 (0.29-1.74) | 0.7 |
| Antibiotic only | 1.11 (1.05-1.17) | <0.0001 | 0.71 (0.57-0.89) | <0.0001 | 0.82 (0.77-0.86) | <0.0001 | 1.03 (0.96-1.12) | 0.4 | 0.67 (0.63-0.72) | <0.0001 |
| Zanamivir +<br>antibiotic | 1.14 (0.28-4.55) | 0.9 | Not estimable |  | 0.74 (0.18-2.94) | 0.7 | 1.57 (0.39-6.29) | 0.5 | - | - |
| HR: Hazard ratio, 95%CI: 95% confidence intervals |  |  |  |  |  |  |  |  |  |  |
| a: Model adjusted by: Sex (men), Age, Indigenous self-identification, Diabetes, Chronic obstructive pulmonary disease, Immunosuppression, Hypertension, Human immunodeficiency virus/acquired immune deficiency syndrome, Cardiovascular disease, Obesity, Chronic kidney disease, Unemployed, Time from symptom onset to medical attention, Fever, Cough, Shortness of breath, Irritability, Chest pain, Chills, Myalgias, Arthralgias, Abrupt deterioration, Polypnea, Abdominal pain, Cyanosis. |  |  |  |  |  |  |  |  |  |  |
| b: Model adjusted by: Sex (men), Age, Indigenous self-identification, Diabetes, Chronic obstructive pulmonary disease, Immunosuppression, Hypertension, Cardiovascular disease, Obesity, Chronic kidney disease, Unemployed, Time from symptom onset to medical attention, Fever, Cough, Shortness of breath, Irritability, Chest pain, Chills, Myalgias, Arthralgias, Abrupt deterioration, Polypnea, Vomit, Abdominal pain, Cyanosis. |  |  |  |  |  |  |  |  |  |  |
| c: Model adjusted by: Sex (men), Age, Diabetes, Chronic obstructive pulmonary disease, Immunosuppression, Hypertension, Cardiovascular disease, Chronic kidney disease, Smoker, Unemployed, Cough, Shortness of breath, Chest pain, Chills, Myalgias, Arthralgias, Abrupt deterioration, Polypnea, Cyanosis. |  |  |  |  |  |  |  |  |  |  |
| d: Model adjusted by: Sex (men), Age, Indigenous self-identification, Diabetes, Chronic obstructive pulmonary disease, Immunosuppression, Hypertension, Hypertension, Human immunodeficiency virus/acquired immune deficiency syndrome, Cardiovascular disease, Obesity, Chronic kidney disease, Unemployed, Time from symptom onset to medical attention, Fever, Cough, Shortness of breath, Irritability, Chest pain, Myalgias, Arthralgias, Polypnea, Vomit, Abdominal pain, Cyanosis. |  |  |  |  |  |  |  |  |  |  |
| e: Model adjusted by: Sex (men), Age, Diabetes, Chronic obstructive pulmonary disease, Immunosuppression, Hypertension, Cardiovascular disease, Chronic kidney disease, Unemployed, Time from symptom onset to medical attention, Cough, Sore throat, Shortness of breath, Chest pain, Headache, Myalgias, Arthralgias, Rhinorrhea, Polypnea, Abdominal pain, Cyanosis. |  |  |  |  |  |  |  |  |  |  |

After matching patients receiving antivirals (Supplementary Table 17), oseltamivir (Supplementary Table 18), or antibiotics (Supplementary Table 19) to controls, mortality risks were similar to those in the unmatched cohort (Table 3). However, contrary to the main analyses, antivirals in hospitalized patients, as well as oseltamivir in hospitalized and critical patients were a risk factor for death. Antibiotics were a protective factor in hospitalized and critical patients, but not in ambulatory and non-critical patients.

**Table 3.**
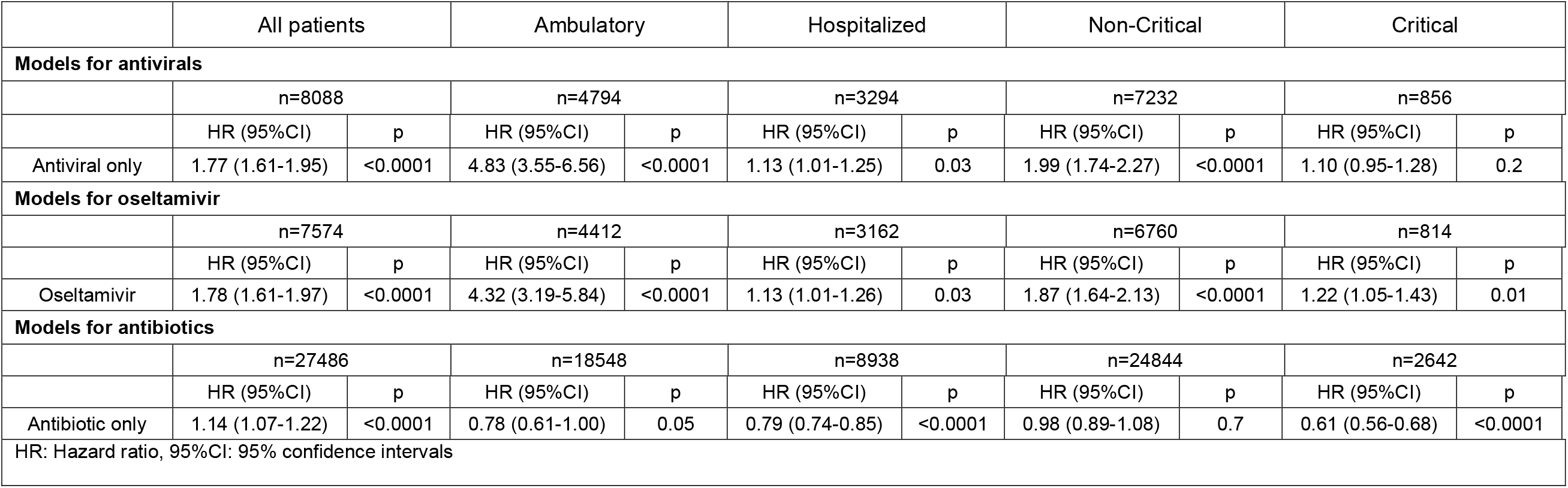
Adjusted mortality risk in laboratory-confirmed COVID-19 patients receiving antivirals, oseltamivir, or antibiotics after propensity score matching.

Density functions before and after matching are shown in (Supplementary Figures 1-3). Of all 8,969 patients receiving antivirals, 10% (n=903) received antivirals before evaluation in COVID-19 accredited units; their baseline and follow-up characteristics are available in Supplementary Table 20. 25.2% (n=227) were admitted to hospital. Median time from symptom onset to initiation of antivirals was 1 day (IQR:0-4) for both ambulatory and hospitalized patients; time from symptom onset to ambulatory care in accredited units was 5 days (IQR:3-8) and 6 days (IQR:4-9) for hospitalization. Time from initiation of antivirals to hospitalization was 3 days (IQR:0-6). Time-to-initiation of antivirals and time-to-hospitalization for specific antivirals are shown in Supplementary Figure 4.

Early (≤2 days) and late (>2 days) initiation of antivirals occurred in 64.2% (n=580) and 35.8% (n=323) patients, respectively. Overall survival in early (91.3%) and late (88.9%) groups was not different (p=0.2). Survival for early/late use of antivirals is shown in Supplementary Table 21. Oseltamivir was associated with increased risk of death in both early (HR=3.00, 95%CI:2.14-4.20) and late (HR=2.99, 95%CI:1.83-4.89) groups, as well as late use of lopinavir-ritonavir (HR=9.9, 95%CI:2.49-39.83); all other early/late antivirals did not reach statistical significance. There were no differences in hospitalization rates between early and late groups for every antiviral (Supplementary Figure 5).

Prescription of oseltamivir during the pandemic period was greater than confirmed influenza cases (Figure 4A) and followed a similar pattern to that of weekly new cases of patients with a positive SARS-CoV-2 test (Figure 4B).

**Figure 4.**
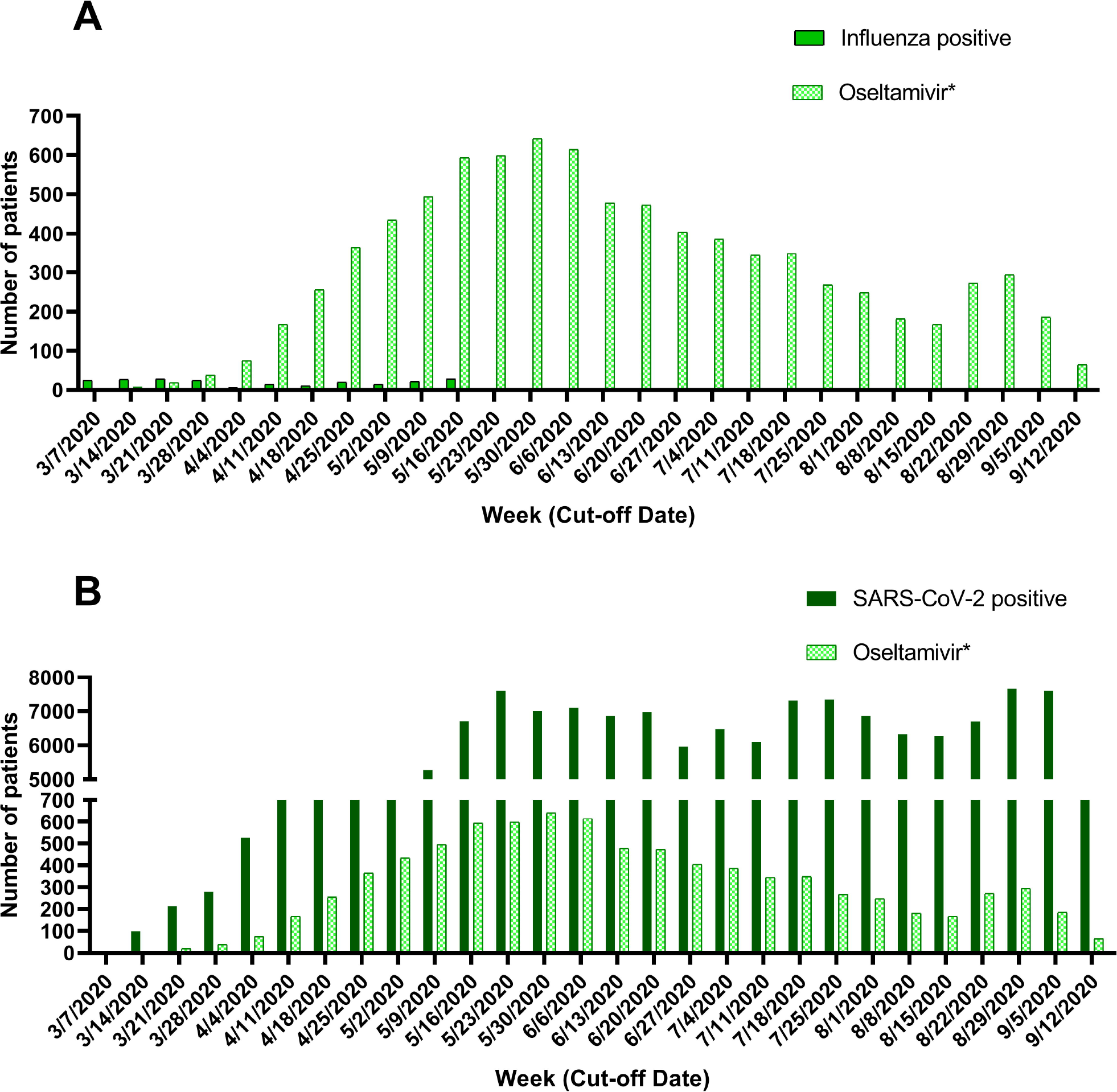
Weekly prescription of oseltamivir and newly diagnosed patients with influenza (a) and COVID-19 (b) during the COVID-19 pandemic in Mexico City. *Data of prescription of oseltamivir for patients tested only for influenza was not collected.

## DISCUSSION

To the best of our knowledge, this is the first observational study evaluating amantadine, rimantadine, zanamivir, and acyclovir for COVID-19; no registered studies to evaluate these drugs exist.^11^ Only one study has evaluated risk of death for oseltamivir;^24^ lopinavir-ritonavir have been evaluated in clinical trials.^25^

We hypothesized that antivirals and antibiotics could be having widespread use in real-world settings as repurposed drugs for COVID-19. Therefore, we studied mortality in laboratory-confirmed COVID-19 patients treated with common antivirals and/or antibiotics in Mexico City. Most patients did not receive them (83.4%), although a substantial proportion received antivirals alone (3.0%) or combined with antibiotics (3.6%) despite national guidelines advising against antivirals out of clinical trials.^15^ Patients receiving antivirals and antibiotics were overall more symptomatic, suggesting that florid clinical presentations and not evidence may be guiding decision to treat.

Our cohort included mostly middle-aged adults (44.2 [SD:16.8] years), of which non-survivors were younger and with a higher burden of hypertension, diabetes and obesity compared with other countries, findings consistent with previous studies in Mexican patients.^26^ Rates of patients with cardiovascular diseases, COPD, asthma, and smokers were low, suggesting that underdiagnosis could be occurring. Only 20-30% of patients had been vaccinated against influenza during the prior season.

Of patients treated with antivirals, ∼10% (n=903) started antivirals before seeking medical attention in accredited COVID-19 units. Hospitalization rates were not different when antivirals were used early (≤2 days) vs late (>2 days). In Mexico, antibiotics and most antivirals are sold under prescription. However, private pharmacy-associated clinics are a rapidly growing sector not included in our study where physicians tend to have lower experience, qualifications, compliance with regulations, and higher prescription rates.^27–29^ Self-medication with amantadine could be occurring since it is a widely available over-the-counter medication. Importantly, the high prescription of oseltamivir in Mexico City during the pandemic period cannot be attributed to influenza outbreaks, since patients receiving oseltamivir largely exceed those with a positive test for influenza and follows a similar weekly pattern to that of newly diagnosed COVID-19 patients.

In one single-center study, oseltamivir was associated with decreased mortality risk in COVID-19-hospitalized patients (HR=0.21; 95%CI:0.10-0.43).^24^ Contrary to Liu et al., we found no benefit for oseltamivir in hospitalized patients (HR=1.07; 95%CI:0.99-1.15) which is consistent with studies of oseltamivir for SARS-CoV (HR=0.87; 95%CI:0.55-1.38).^30^ Combination of oseltamivir with antibiotics in hospitalized patients in our study resulted in decreased risk of death (HR=0.92; 95%:0.87-0.98), which could explain findings by Liu et al. since most patients in their cohort (87.7%) received antibiotics. Decreased mortality is likely driven by antibiotics since hospitalized patients in our study receiving only antibiotics had lower risk of dying (HR=0.81, 95%CI:0.77-0.86) than antibiotics plus oseltamivir.

After multivariable adjustment, oseltamivir was associated with increased mortality in the general population (HR=1.72, 95%CI:1.61-1.84), ambulatory (HR=4.79, 95%CI:4.01-5.75), non-critical (HR=2.05, 95%CI:1.88-2.23), and pregnant (HR=8.35, 95%CI:1.77-39.30) patients. Mortality risk was also higher in the cohort of 903 patients with both early (HR=3.00, 95%CI:2.14-4.20) and late (HR=2.99, 95%CI:1.83-4.89) use of oseltamivir. We performed propensity score matching analyses to further account for potential confounders and found that hospitalized (HR=1.13, 95%CI:1.01-1.26) and critical patients (HR:1.22, 95%CI:1.05-1.43) also had an increased risk of death when treated with oseltamivir. Further efforts to limit bias were adjustment for center through GEE and calculation of E-values which support a strong association between oseltamivir and increased mortality risk, since potential confounders not accounted for in our study should have substantially large hazard ratios and lower limit confidence intervals (i.e. 9.05 and 7.48, respectively, for ambulatory patients) to explain our findings.

The potentially inhibitory activity of proteases by oseltamivir^5^ was found to be weak through molecular modeling, while inhibition of SARS-CoV-2 *in vitro* and reduction of symptoms in hospitalized patients failed.^31^ Furthermore, oseltamivir nearly suppresses the production of pro-inflammatory cytokines by dendritic cells, polymorphonuclear leukocytes, and CD8+ T cells through inhibition of endogenous neuraminidase (sialidase),^32^ which could impair the immune response and limit the capacity to eliminate the infection. Use of oseltamivir for infections caused by viruses lacking a neuraminidase gene (i.e. respiratory syncytial virus) decrease viral clearance,^33^ which could explain increased mortality in patients receiving oseltamivir. Additionally, sudden (cardiac arrest, respiratory suppression, hypothermia, and neuropsychiatric events) and delayed-onset (impaired renal function, hyperglycemic events, prolonged QTc interval, immunological impairment, and bleeding) adverse reactions could explain increased risk of death in patients with COVID-19 treated with oseltamivir.^32^ Antiviral drug-related heart damage is a concern since some antivirals may be cardiotoxic, aggravating myocardial damage caused by SARS-CoV-2.^34^ Renal and psychiatric adverse events in patients receiving oseltamivir were higher compared to placebo in a systematic review.^35^

In the RECOVERY study, there were no differences in mortality risk between hospitalized patients receiving lopinavir-ritonavir vs placebo (HR=1.03, 95%CI:0.91-1.17),^25^ which is consistent with our findings in hospitalized patients. Notably, ambulatory and late (>2 days) use of lopinavir-ritonavir were risk factors for death in our study.

Antibiotics were a risk factor for death in the general population after multivariable analysis but a protective factor in both ambulatory and hospitalized patients. Nonetheless, univariate models showed no overall effect of antibiotics in ambulatory patients; when adjusting only for demographic variables no effect persisted but were protective after adjusting only for clinical variables. This is explained by the fact that more symptomatic patients received antibiotics more often. Supporting this conclusion, no benefit was observed for antibiotics in non-critical patients in both matched and unmatched cohorts.

We observed a potential benefit for use of antibiotics in hospitalized, IMV, and critical patients. Increased survival in these groups could be due to prevention or treatment of concomitant bacterial infections, thereby supporting current WHO recommendations.^14^ However, categorization of antibiotics as a single category in this dataset limits our study. Randomized clinical trials testing antibiotics in severe COVID-19 adult patients to prevent secondary infections would be important to corroborate this hypothesis.

For children and adolescents, antibiotics were a risk factor for death (HR=4.22, 95%CI:2.01-8.86). However, we did not differentiate ambulatory from hospitalized pediatric patients and current recommendations include using antibiotics in hospitalized patients with multisystem inflammatory syndrome.^36^ The lack of benefit from antivirals included in our study in pediatric patients supports current guidelines discouraging their use.^37^

The main limitation of our study is that we were not able to assess cointerventions being studied for COVID-19 since only data for antivirals and antibiotics were available. Steroids increase survival in patients requiring oxygen administration and decrease survival in patients without supplementary oxygen.^38^ However, modifications of mortality risks by steroids in our population should be large according to E-values, reducing the likelihood that these associations could be due to steroids.

Another potential limitation is that Mexico has had a low diagnostic testing rate for SARS-CoV-2 (0.08 daily tests per 1,000 people).^39^ However, health authorities require 100% of patients with severe disease to be tested. Since we only studied mortality, an outcome expected to occur in patients who progress to severe disease, our study feasibly included most events. Nonetheless, excess mortality rates suggest there could be an undercounting of deaths in Mexico City.^39^ These patients could have refrained from seeking medical attention or received medical care in non-accredited COVID-19 units where mortality, quality of care, and use of antivirals/antibiotics could be different. Also, the number of ICU beds in Mexico City was relatively low in March 2020 (6.0 per 100,000 population) compared to most European countries (5 to 33.9 per 100,000) in the pre-pandemic period; this capacity was expanded to 29.5 ICU beds per 100,000 by September 2020.^39,40^ Mortality rates, especially in patients younger than 60 years, are lower under high availability of ICU beds.^40^ Thus, mortality rates could have varied throughout our study period.

Although we were not able to determine duration of follow-up in our study, the mechanisms and resources used by epidemiologic authorities in Mexico are robust enough to guarantee adequate matching of patients who had completed follow-up with death certificates. Thus, our finding that 92.7% (95%CI:92.2-93.2) and 99.6% (95%CI:99.5-99.7) of deaths occurred by day 28 and 56, respectively, could be important for the interpretation and design of COVID-19 clinical trials assessing short-term mortality.

## CONCLUSIONS

Repurposed antivirals (oseltamivir, zanamivir, amantadine, rimantadine, acyclovir, and lopinavir/ritonavir) did not provide additional benefit, whereas oseltamivir consistently increased mortality risk. Antibiotics were associated with increased risk of death in the general population but decreased mortality risk only in hospitalized and critical patients, thereby supporting current WHO recommendations.

## Supporting information

More symptomatic ambulatory patients received antivirals and antibiotics more frequently (Supplementary Table 1); hospitalized patients with more sign

Upon evaluating a patient suspected of having COVID-19, healthcare professionals are required to fill out a format (Supplementary Appendix) containing

## Data Availability

Data are available upon reasonable request

## ACKNOWLEDGEMENTS

We thank Dr. César Sebastián Salinas Nájera, Head of Epidemiology in “Hospital Materno Infantil Magdalena Contreras” of Mexico City’s Health Secretariat for his valuable clarifications of epidemiologic surveillance mechanisms in Mexico City. We want to acknowledge the Government of Mexico City for their effort to increase transparency and aid research by providing open datasets in their Open Data Platform; this sort of actions is most needed in low and middle-income countries to understand and solve the problems of our people. JMG would like to thank “Dirección General de Calidad y Educación en Salud” for supporting his participation in “Programa Nacional de Servicio Social en Investigación en Salud”.

## CONFLICTS OF INTEREST STATEMENT

The authors declare no competing interests.

## FUNDING

This research did not receive any specific grant from funding agencies in the public, commercial, or not-for-profit sectors.

## Data availability statement

The data that support the findings of this study are openly available in the Open Data Platform of Mexico City’s Government at https://datos.cdmx.gob.mx/explore/dataset/base-covid-sinave/information/, reference number 19, and the Directorate General of Epidemiology of Mexico City’s Weekly Epidemiological Surveillance Reports of Influenza 2020 at https://www.gob.mx/salud/documentos/informes-semanales-para-la-vigilancia-epidemiologica-de-influenza-2020, reference number 21.

## Author contributions

Conception and design: JMG and AKG; Analysis and interpretation: AKG, JMG, JMS, JOGM, and HIRG; Drafting the manuscript, making tables and figures: All authors; Revising the manuscript for important intellectual content: AKG, JOGM, and HIRG. All authors approved the final draft of the manuscript.

## Notes

### Competing Interest Statement

The authors have declared no competing interest.

### Author Declarations

This is a retrospective study using an open-source dataset of patients receiving medical care for suspected COVID-19 in Mexico City. The Secretariat of Health of Mexico approved the collection and publication of data.

