## Supplementary material for "All-cause mortality among patients treated with repurposed antivirals and antibiotics for COVID-19 in Mexico City: A Real-World Observational Study": More symptomatic ambulatory patients received antivirals and antibiotics more frequently (Supplementary Table 1); hospitalized patients with more sign

**Supplementary Table 1.** Signs and symptoms in ambulatory patients

**Supplementary** **Table 2.** Signs and symptoms in hospitalized patients

**Supplementary** **Table 3.** Baseline and follow-up characteristics of survivors and non-survivors

**Supplementary Table 4.** Survival analysis of patients undergoing IMV

**Supplementary Table 5.** Survival analysis of patients admitted to ICU

**Supplementary Table 6.** Survival analysis of non-pregnant/puerperal adults

**Supplementary Table 7.** Survival analysis of children and adolescents

**Supplementary Table 8.** Survival analysis of pregnant women

**Supplementary Table 9.** Unadjusted Cox regression models for mortality in all patients, ambulatory, hospitalized, non-critical, and critical patients

**Supplementary Table 10.** Mortality risk in patients undergoing IMV

**Supplementary Table 11.** Mortality risk in patients admitted to ICU

**Supplementary Table 12.** Mortality risk in non-pregnant/puerperal adults

**Supplementary Table 13.** Mortality risk in children and adolescents

**Supplementary Table 14.** Mortality risk in pregnant women

**Supplementary Table 15.** E-values for the general population, ambulatory, hospitalized, non-critical, and critical patients

**Supplementary Table 16.** E-values for IMV, ICU, Non-pregnant/puerperal adults, and children and adolescents

**Supplementary Table 17.** Baseline characteristics of propensity score-matched patients treated with Antivirals and controls.

**Supplementary Table 18.** Baseline characteristics of propensity score-matched patients treated with Oseltamivir and controls.

**Supplementary Table 19.** Baseline characteristics of propensity score-matched patients treated with Antibiotics and controls.

**Supplementary Table 20.** Baseline characteristics of patients with complete dates of antiviral initiation

**Supplementary Table 21.** Survival analysis of early and late use of antivirals

**Supplementary Figure 1.** Density functions of propensity score-matched patients treated with Antivirals and controls before and after matching

**Supplementary Figure 2.** Density functions of propensity score-matched patients treated with Oseltamivir and controls before and after matching

**Supplementary Figure 3.** Density functions of propensity score-matched patients treated with Antibiotics and controls before and after matching

**Supplementary Figure 4.** Timing of symptom onset, initiation of antiviral, and hospitalization

**Supplementary Figure 5.** Comparison of hospitalization rates between early and late use of antivirals

**Supplementary Appendix.** Case form: Epidemiologic study of suspected viral respiratory disease

Supplementary Table 1. Signs and symptoms in laboratory-confirmed COVID-19 ambulatory patients treated with antivirals and/or antibiotics in 603 accredited medical units in Mexico City between February 24, 2020 and September 14, 2020.

|  | **No antiviral/ antibiotic**  **n=98060** | **Acyclovir**  **n=30** | **Amantadine**  **n=282** | **Lopinavir-Ritonavir**  **n=8** | **Oseltamivir**  **n=3187** | **Rimantadine**  **n=48** | **Zanamivir**  **n=19** | **Antibiotic only**  **n=8268** | **Total**  **n=109902** | **p value** |
| --- | --- | --- | --- | --- | --- | --- | --- | --- | --- | --- |
| Number of signs and symptoms, mean (SD) | 5.8 (3.8) | 6.8 (3.3)^a^ | 7.2 (3.6) | 7.9 (5.3) | 7.8 (3.3)^a^ | 7.6 (3.2)^a^ | 6.7 (4.6) | 7.0 (3.5)^a^ | 5.9 (3.8) | <0.0001 |
| Fever | 52966 (54) | 21 (70) | 169 (59.9) | 5 (62.5) | 2465 (77.3) | 31 (64.6) | 13 (68.4) | 5471 (66.2) | 61141 (55.6) | <0.0001 |
| Cough | 65216 (66.5) | 19 (63.3) | 216 (76.6) | 6 (75) | 2583 (81) | 34 (70.8) | 15 (78.9) | 6071 (73.4) | 74160 (67.5) | <0.0001 |
| Sore throat | 41927 (42.8) | 12 (40) | 144 (51.1) | 3 (37.5) | 1611 (50.5) | 24 (50) | 5 (26.3) | 4388 (53.1) | 48114 (43.8) | <0.0001 |
| Shortness of breath | 19091 (19.5) | 11 (36.7) | 63 (22.3) | 4 (50) | 1010 (31.7) | 12 (25) | 8 (42.1) | 2237 (27.1) | 22436 (20.4) | <0.0001 |
| Irritability | 16364 (16.7) | 5 (16.7) | 70 (24.8) | 2 (25) | 720 (22.6) | 11 (22.9) | 7 (36.8) | 1593 (19.3) | 18772 (17.1) | <0.0001 |
| Diarrhea | 22096 (22.5) | 10 (33.3) | 86 (30.5) | 3 (37.5) | 981 (30.8) | 12 (25) | 7 (36.8) | 2456 (29.7) | 25651 (23.3) | <0.0001 |
| Chest pain | 23150 (23.6) | 9 (30) | 87 (30.9) | 2 (25) | 1158 (36.3) | 19 (39.6) | 7 (36.8) | 2494 (30.2) | 26926 (24.5) | <0.0001 |
| Chills | 32621 (33.3) | 14 (46.7) | 139 (49.3) | 6 (75) | 1585 (49.7) | 27 (56.3) | 5 (26.3) | 3482 (42.1) | 37879 (34.5) | <0.0001 |
| Headache | 67181 (68.5) | 21 (70) | 202 (71.6) | 6 (75) | 2575 (80.8) | 31 (64.6) | 13 (68.4) | 6022 (72.8) | 76051 (69.2) | <0.0001 |
| Myalgias | 47106 (48) | 23 (76.7) | 175 (62.1) | 5 (62.5) | 2053 (64.4) | 32 (66.7) | 8 (42.1) | 4993 (60.4) | 54395 (49.5) | <0.0001 |
| Arthralgias | 42161 (43) | 18 (60) | 155 (55) | 4 (50) | 1881 (59) | 31 (64.6) | 6 (31.6) | 4530 (54.8) | 48786 (44.4) | <0.0001 |
| Abrupt deterioration | 38549 (39.3) | 15 (50) | 157 (55.7) | 5 (62.5) | 1923 (60.3) | 24 (50) | 6 (31.6) | 4471 (54.1) | 45150 (41.1) | <0.0001 |
| Rhinorrhea | 28354 (28.9) | 6 (20) | 107 (37.9) | 4 (50) | 1079 (33.9) | 23 (47.9) | 6 (31.6) | 2744 (33.2) | 32323 (29.4) | <0.0001 |
| Polypnea | 7412 (7.6) | 3 (10) | 37 (13.1) | 3 (37.5) | 381 (12) | 3 (6.3) | 3 (15.8) | 859 (10.4) | 8701 (7.9) | <0.0001 |
| Vomit | 6406 (6.5) | 2 (6.7) | 18 (6.4) | 0 (0) | 309 (9.7) | 6 (12.5) | 1 (5.3) | 696 (8.4) | 7438 (6.8) | <0.0001 |
| Abdominal pain | 11502 (11.7) | 3 (10) | 49 (17.4) | 0 (0) | 646 (20.3) | 8 (16.7) | 4 (21.1) | 1065 (12.9) | 13277 (12.1) | <0.0001 |
| Conjunctivitis | 12784 (13) | 4 (13.3) | 53 (18.8) | 3 (37.5) | 475 (14.9) | 11 (22.9) | 5 (26.3) | 1315 (15.9) | 14650 (13.3) | <0.0001 |
| Cyanosis | 2843 (2.9) | 1 (3.3) | 9 (3.2) | 1 (12.5) | 152 (4.8) | 6 (12.5) | 2 (10.5) | 245 (3) | 3259 (3) | <0.0001 |
| Sudden onset of symptoms | 32348 (33) | 7 (23.3) | 93 (33) | 1 (12.5) | 1359 (42.6) | 19 (39.6) | 7 (36.8) | 3067 (37.1) | 36901 (33.6) | <0.0001 |
| Data expressed as frequency (%). P values were calculated by *X^2^* or One-way ANOVA.  a: Statistical difference with respect to No antiviral/antibiotic.  SD: Standard deviation | | | | | | | | | | |

Supplementary Table 2. Signs and symptoms in laboratory-confirmed COVID-19 hospitalized patients treated with antivirals and/or antibiotics in 329 accredited medical units in Mexico City between February 24, 2020 and September 14, 2020.

|  | **No antiviral / antibiocitv**  **n=16083** | **Acyclovir**  **n=6** | **Amantadine**  **n=37** | **Lopinavir-Ritonavir**  **n=92** | **Oseltamivir**  **n=5227** | **Rimantadine**  **n=13** | **Zanamivir**  **n=20** | **Antibiotic only**  **n=5475** | **Total**  **n=26953** | **p value** |
| --- | --- | --- | --- | --- | --- | --- | --- | --- | --- | --- |
| Number of signs and symptoms, mean (SD) | 7.9 (3.4) | 6 (2.6) | 7.9 (3.3) | 6.6 (2.9)^a^ | 8.1 (3.4)^a^ | 8.8 (2.4) | 9.4 (2.8) | 6.9 (3.2)^a^ | 7.8 (3.4) | <0.0001 |
| Fever | 13045 (81.1) | 4 (66.7) | 29 (78.4) | 76 (82.6) | 4498 (86.1) | 11 (84.6) | 18 (90) | 4298 (78.5) | 21979 (81.5) | <0.0001 |
| Cough | 13151 (81.8) | 5 (83.3) | 29 (78.4) | 61 (66.3) | 4472 (85.6) | 8 (61.5) | 20 (100) | 4300 (78.5) | 22046 (81.8) | <0.0001 |
| Sore throat | 6918 (43) | 0 (0) | 15 (40.5) | 28 (30.4) | 2090 (40) | 5 (38.5) | 17 (85) | 1853 (33.8) | 10926 (40.5) | <0.0001 |
| Shortness of breath | 11970 (74.4) | 3 (50) | 22 (59.5) | 62 (67.4) | 4160 (79.6) | 10 (76.9) | 15 (75) | 4264 (77.9) | 20506 (76.1) | <0.0001 |
| Irritability | 3096 (19.3) | 0 (0.0) | 11 (29.7) | 5 (5.4) | 1273 (24.4) | 3 (23.1) | 6 (3) | 932 (17) | 5326 (19.8) | <0.0001 |
| Diarrhea | 3725 (23.2) | 1 (16.7) | 10 (27) | 16 (17.4) | 1199 (22.9) | 3 (23.1) | 6 (30) | 1038 (19) | 5998 (22.3) | <0.0001 |
| Chest pain | 6374 (39.6) | 3 (50.0) | 18 (48.6) | 20 (21.7) | 1821 (34.8) | 6 (46.2) | 8 (40) | 1675 (30.6) | 9925 (36.8) | <0.0001 |
| Chills | 6784 (42.2) | 2 (33.3) | 19 (51.4) | 28 (30.4) | 2031 (38.9) | 6 (46.2) | 12 (60) | 1521 (27.8) | 10403 (38.6) | <0.0001 |
| Headache | 11712 (72.8) | 1 (16.7) | 25 (67.6) | 53 (57.6) | 3773 (72.2) | 9 (69.2) | 16 (80) | 3378 (61.7) | 18967 (70.4) | <0.0001 |
| Myalgias | 10086 (62.7) | 4 (66.7) | 26 (70.3) | 55 (59.8) | 3021 (57.8) | 11 (84.6) | 16 (80) | 3052 (55.7) | 16271 (60.4) | <0.0001 |
| Arthralgias | 9631 (59.9) | 4 (66.7) | 24 (64.9) | 44 (47.8) | 2965 (56.7) | 11 (84.6) | 17 (85) | 2899 (52.9) | 15595 (57.9) | <0.0001 |
| Abrupt deterioration | 10442 (64.9) | 5 (83.3) | 23 (62.2) | 61 (66.3) | 3444 (65.9) | 10 (76.9) | 14 (70) | 3311 (60.5) | 17310 (64.2) | <0.0001 |
| Rhinorrhea | 3781 (23.5) | 0 (0) | 11 (29.7) | 16 (17.4) | 1204 (23) | 4 (30.8) | 5 (25) | 944 (17.2) | 5965 (22.1) | <0.0001 |
| Polypnea | 4565 (28.4) | 0 (0) | 9 (24.3) | 10 (10.9) | 1466 (28) | 4 (30.8) | 3 (15) | 1110 (20.3) | 7167 (26.6) | <0.0001 |
| Vomit | 1688 (10.5) | 1 (16.7) | 5 (13.5) | 7 (7.6) | 561 (10.7) | 2 (15.4) | 1 (5) | 420 (7.7) | 2685 (10) | <0.0001 |
| Abdominal pain | 2578 (16) | 0 (0) | 11 (29.7) | 8 (8.7) | 915 (17.5) | 2 (15.4) | 3 (15) | 544 (9.9) | 4061 (15.1) | <0.0001 |
| Conjunctivitis | 1493 (9.3) | 0 (0) | 2 (5.4) | 4 (4.3) | 487 (9.3) | 0 (0) | 1 (5) | 304 (5.6) | 2291 (8.5) | <0.0001 |
| Cyanosis | 1618 (10.1) | 0 (0) | 3 (8.1) | 3 (3.3) | 664 (12.7) | 2 (15.4) | 1 (5) | 367 (6.7) | 2658 (9.9) | <0.0001 |
| Sudden onset of symptoms | 5259 (32.7) | 3 (50) | 3 (8.1) | 47 (51.1) | 2472 (47.3) | 7 (53.8) | 9 (45) | 2022 (36.9) | 9822 (36.4) | <0.0001 |
| Data expressed as frequency (%). P values were calculated by *X^2^* or One-way ANOVA.  a: Statistical difference with respect to No antiviral/antibiotic.  SD: Standard deviation | | | | | | | | | | |

Supplementary Table 3. Baseline and follow-up characteristics of laboratory confirmed COVID-19 survivors and non-survivors who received medical attention in 688 accredited units in Mexico City between February 24, 2020 and September 14, 2020.

|  | **Total population**  **n=136855** | **Survivors**  **n=125176** | **Non-survivors**  **n=11679** | **p Value** |
| --- | --- | --- | --- | --- |
| Sex |  |  |  |  |
| Women | 66683 (48.7) | 62907 (50.3) | 3776 (32.3) | <0.0001 |
| Men | 70172 (51.3) | 62269 (49.7) | 7903 (67.7) |  |
| Age, mean (SD) | 44.2 (16.8) | 42.5 (15.9) | 62 (14.1) | <0.0001 |
| Age categories |  |  |  |  |
| 0-19 years | 7558 (5.5) | 7514 (6) | 44 (0.4) | <0.0001 |
| 20-29 years | 20098 (14.7) | 19974 (16) | 124 (1.1) |  |
| 30-39 years | 29434 (21.5) | 28909 (23.1) | 525 (4.5) |  |
| 40-49 years | 29553 (21.6) | 28084 (22.4) | 1469 (12.6) |  |
| 50-59 years | 24928 (18.2) | 22298 (17.8) | 2630 (22.5) |  |
| 60-69 years | 15070 (11) | 11840 (9.5) | 3230 (27.7) |  |
| 70-79 years | 7183 (5.2) | 4731 (3.8) | 2452 (21) |  |
| 80-89 years | 2594 (1.9) | 1540 (1.2) | 1054 (9) |  |
| 90-99 years | 419 (0.3) | 270 (0.2) | 149 (1.3) |  |
| ≥100 years | 18 (0.01) | 16 (0.01) | 2 (0.001) |  |
| Indigenous self-identification | 713 (0.5) | 639 (0.5) | 74 (0.6) | 0.08 |
| Occupation |  |  |  |  |
| Technical services | 1916 (1.4) | 1702 (1.4) | 214 (1.8) | <0.0001 |
| Education | 10006 (7.3) | 9923 (7.9) | 83 (0.7) |  |
| Healthcare | 17281 (12.6) | 16954 (13.5) | 327 (2.8) |  |
| Agricultural activities | 302 (0.2) | 244 (0.2) | 58 (0.5) |  |
| Commerce | 50450 (36.9) | 47070 (37.6) | 3380 (28.9) |  |
| Other | 24630 (18) | 22383 (17.9) | 2247 (19.2) |  |
| Unemployed | 5685 (4.2) | 4539 (3.6) | 1146 (9.8) |  |
| Stay-at-home | 26585 (19.4) | 22361 (17.9) | 4224 (36.2) |  |
| Last-season flu vaccination | 25691 (19.7) | 24570 (20.7) | 1121 (9.6) | <0.0001 |
| Special populations |  |  |  |  |
| Pregnancy | 583 (0.9) | 573 (0.9) | 10 (0.3) | <0.0001 |
| Age during pregnancy, mean (SD) | 29.8 (7.4) | 29.8 (7.5) | 32.7 (6) | 0.2 |
| Last-season flu vaccination | 161 (27.6) | 158 (27.6) | 3 (30) | 0.9 |
| Pregnancy Age |  |  |  |  |
| Early adolescent (≤14 years) | 2 (0.3) | 2 (0.3) | 0 (0) | 0.2 |
| Late adolescent (15-19 years) | 34 (5.8) | 34 (5.9) | 0 (0) |  |
| Normal age (20-34 years) | 404 (69.3) | 398 (69.5) | 6 (60) |  |
| Advanced maternal age (≥35 years) | 143 (24.5) | 139 (24.3) | 4 (40) |  |
| Trimester of pregnancy |  |  |  |  |
| First trimester | 114 (19.6) | 114 (19.9) | 0 (0.0) | <0.0001 |
| Second trimester | 177 (30.4) | 174 (30.4) | 3 (30) |  |
| Third trimester | 292 (50.1) | 285 (49.7) | 7 (70) |  |
| Puerperium | 103 (0.2) | 102 (0.2) | 1 (0.03) | 0.9 |
| Days of puerperium |  |  |  |  |
| 1 day | 33 (32) | 32 (31.4) | 1 (100) | 0.2 |
| 2-7 days | 33 (32) | 33 (32.4) | 0 (0) |  |
| 8-42 days | 37 (35.9) | 37 (36.3) | 0 (0) |  |
| Age during puerperium, mean (SD) | 31.9 (9.7) | 31.9 (9.7) | 24 | 0.5 |
| Last-season flu vaccination | 22 (21.4) | 22 (21.6) | 0 (0) | 0.6 |
| Children and adolescents (<18 years) | 5791 (4.2) | 5751 (4.6) | 40 (0.3) | <0.0001 |
| Age, mean (SD) | 10.9 (5.2) | 10.9 (5.2) | 7.5 (6.4) | <0.0001 |
| Last-season flu vaccination | 1213 (20.9) | 1211 (21.1) | 2 (5) | 0.01 |
| Non-pregnant/puerperal adults (≥18 years) | 130378 (95.3) | 118750 (94.9) | 11628 (99.6) | <0.0001 |
| Age, mean (SD) | 45.7 (15.5) | 44.1 (14.7) | 62.2 (13.7) | <0.0001 |
| Last-season flu vaccination | 25691 (19.7) | 24570 (20.7) | 1121 (9.6) | <0.0001 |
| Comorbidities |  |  |  |  |
| Diabetes | 18229 (13.3) | 14136 (11.3) | 4093 (35) | <0.0001 |
| COPD | 1741 (1.3) | 1184 (0.9) | 557 (4.8) | <0.0001 |
| Asthma | 3035 (2.2) | 2858 (2.3) | 177 (1.5) | <0.0001 |
| Immunosuppression | 1758 (1.3) | 1380 (1.1) | 378 (3.2) | <0.0001 |
| Hypertension | 22185 (16.2) | 17574 (14) | 4611 (39.5) | <0.0001 |
| HIV/AIDS | 573 (0.4) | 509 (0.4) | 64 (0.5) | 0.02 |
| Cardiovascular disease | 2724 (2) | 2128 (1.7) | 596 (5.1) | <0.0001 |
| Obesity | 23848 (17.4) | 21072 (16.8) | 2776 (23.8) | <0.0001 |
| Chronic kidney disease | 2067 (1.5) | 1298 (1) | 769 (6.6) | <0.0001 |
| Smoker | 14727 (10.8) | 13437 (10.7) | 1290 (11) | 0.3 |
| Time from symptom onset to medical attention | 4.5 (3.9) | 4.5 (3.8) | 4.9 (3.9) | <0.0001 |
| Baseline symptoms |  |  |  |  |
| Fever | 83120 (60.7) | 73561 (58.8) | 9559 (81.8) | <0.0001 |
| Cough | 96206 (70.3) | 86484 (69.1) | 9722 (83.2) | <0.0001 |
| Sore throat | 59040 (43.1) | 54325 (43.4) | 4715 (40.4) | <0.0001 |
| Shortness of breath | 42942 (31.4) | 33762 (27) | 9180 (78.6) | <0.0001 |
| Irritability | 24098 (17.6) | 21767 (17.4) | 2331 (20) | <0.0001 |
| Diarrhea | 31649 (23.1) | 29141 (23.3) | 2508 (21.5) | <0.0001 |
| Chest pain | 36851 (26.9) | 32498 (26) | 4353 (37.3) | <0.0001 |
| Chills | 48282 (35.3) | 43660 (34.9) | 4622 (39.6) | <0.0001 |
| Headache | 95018 (69.4) | 86872 (69.4) | 8146 (69.7) | 0.4 |
| Myalgias | 70666 (51.6) | 63547 (50.8) | 7119 (61.0) | <0.0001 |
| Arthralgias | 64381 (47) | 57539 (46) | 6842 (58.6) | <0.0001 |
| Abrupt deterioration | 62460 (45.6) | 54794 (43.8) | 7666 (65.6) | <0.0001 |
| Rhinorrhea | 38288 (28) | 35677 (28.5) | 2611 (22.4) | <0.0001 |
| Polypnea | 15868 (11.6) | 12283 (9.8) | 3585 (30.7) | <0.0001 |
| Vomit | 10123 (7.4) | 8956 (7.2) | 1167 (10) | <0.0001 |
| Abdominal pain | 17338 (12.7) | 15639 (12.5) | 1699 (14.5) | <0.0001 |
| Conjunctivitis | 16941 (12.4) | 15965 (12.8) | 976 (8.4) | <0.0001 |
| Cyanosis | 5917 (4.3) | 4430 (3.5) | 1487 (12.7) | <0.0001 |
| Sudden onset of symptoms | 46723 (34.1) | 42636 (34.1) | 4087 (35) | 0.04 |
| Contact with confirmed COVID-19 case | 70923 (51.8) | 69506 (55.5) | 1417 (12.1) | <0.0001 |
| Type of treatment received |  |  |  |  |
| No antiviral / antibiotic | 114143 (83.4) | 106792 (85.3) | 7351 (62.9) | <0.0001 |
| Antiviral only | 4044 (3) | 2980 (2.4) | 1064 (9.1) |  |
| Antibiotic only | 13743 (10) | 11788 (9.4) | 1955 (16.7) |  |
| Antiviral + antibiotic | 4925 (3.6) | 3616 (2.9) | 1309 (11.2) |  |
| Type of antiviral used |  |  |  |  |
| Acyclovir | 36 (0.03) | 32 (0.03) | 4 (0.03) | <0.0001 |
| Amantadine | 319 (0.2) | 304 (0.2) | 15 (0.1) |  |
| Lopinavir-Ritonavir | 100 (0.1) | 75 (0.1) | 25 (0.2) |  |
| Oseltamivir | 8414 (6.1) | 6100 (4.9) | 2314 (19.8) |  |
| Rimantadine | 61 (0.04) | 54 (0.04) | 7 (0.1) |  |
| Zanamivir | 39 (0.03) | 31 (0.02) | 8(0.1) |  |
| Data expressed as frequency (%) or median (SD). P values were calculated by *X^2^* or Student’s t-test.  SD: Standard deviation, COPD: Chronic obstructive pulmonary disease, HIV/AIDS: Human immunodeficiency virus/acquired immune deficiency syndrome. | | | | |

Supplementary Table 4. Survival analysis of laboratory-confirmed COVID-19 patients who underwent invasive mechanical ventilation (IMV) and were treated with antivirals and/or antibiotics in one of 197 accredited hospitals in Mexico City, between February 24, 2020 and September 14, 2020.

| **Group** | **Total** | **Deaths** | **Survivors** | **Survival** | **p value** |
| --- | --- | --- | --- | --- | --- |
| Patients receiving IMV | 6752 | 5346 | 1406 | 20.8% | - |
| Type of treatment | | | | | |
| No antiviral/antibiotic | 3931 | 3240 | 691 | 17.6% | Reference |
| Antiviral only | 507 | 431 | 76 | 15.0% | 0.001 |
| Antibiotic only | 1586 | 1087 | 499 | 31.5% | <0.0001 |
| Antiviral + Antibiotic | 768 | 621 | 147 | 19.1% | 0.3 |
| Type of antiviral or antibiotic only | | | | | |
| No antiviral/antibiotic | 3931 | 3240 | 691 | 17.6% | Reference |
| Acyclovir | 2 | 2 | 0 | 0.0% | 0.6 |
| Amantadine | 7 | 7 | 0 | 0.0% | 0.5 |
| Lopinavir-Ritonavir | 23 | 16 | 7 | 30.4% | 0.1 |
| Oseltamivir | 1235 | 1019 | 216 | 17.5% | 0.2 |
| Rimantadine | 3 | 3 | 0 | 0.0% | 0.4 |
| Zanamivir | 5 | 5 | 0 | 0.0% | 0.9 |
| Antibiotic only | 1586 | 1087 | 499 | 31.5% | <0.0001 |
| Treatment with oseltamivir | | | | | |
| No antiviral/antibiotic | 3931 | 3240 | 691 | 17.6% | Reference |
| Oseltamivir | 488 | 413 | 75 | 15.4% | 0.01 |
| Antibiotic only | 1586 | 1087 | 499 | 31.5% | <0.0001 |
| Oseltamivir + Antibiotic | 747 | 606 | 141 | 18.9% | 0.4 |
| Data expressed as number of patients or percentual of survival. P values were calculated by Log-Rank test. | | | | | |

Supplementary Table 5. Survival analysis of laboratory-confirmed COVID-19 patients treated with antivirals and/or antibiotics and admitted to one of 125 accredited intensive care units (ICU) in Mexico City, between February 24, 2020 and September 14, 2020.

| **Group** | **Total** | **Deaths** | **Survivors** | **Survival** | **p value** |
| --- | --- | --- | --- | --- | --- |
| Patients admitted to ICU | 2425 | 1475 | 950 | 39.2% | - |
| Type of treatment | | | | | |
| No antiviral/antibiotic | 908 | 494 | 414 | 45.6% | Reference |
| Antiviral only | 77 | 53 | 24 | 31.2% | 0.001 |
| Antibiotic only | 1011 | 610 | 401 | 39.7% | 0.02 |
| Antiviral + Antibiotic | 461 | 336 | 125 | 27.1% | <0.0001 |
| Antiviral or antibiotic only | | | | | |
| No antiviral/antibiotic | 908 | 494 | 414 | 45.6% | Reference |
| Acyclovir | 1 | 1 | 0 | 0.0% | 0.08 |
| Amantadine | 2 | 0 | 2 | 100.0% | 0.20 |
| Lopinavir-Ritonavir | 29 | 17 | 12 | 41.4% | 0.40 |
| Oseltamivir | 506 | 371 | 135 | 26.7% | <0.0001 |
| Antibiotic only | 1011 | 610 | 401 | 39.7% | 0.02 |
| Oseltamivir | | | | | |
| No antiviral/antibiotic | 908 | 494 | 414 | 45.6% | Reference |
| Oseltamivir | 66 | 48 | 18 | 27.3% | <0.0001 |
| Antibiotic only | 1011 | 610 | 401 | 39.7% | 0.02 |
| Oseltamivir + Antibiotic | 440 | 323 | 117 | 26.6% | <0.0001 |
| Data expressed as number of patients or percentual of survival. P values were calculated by Log-Rank test. | | | | | |

Supplementary Table 6. Survival analysis of laboratory-confirmed COVID-19 non-pregnant/puerperal adults treated with antivirals and/or antibiotics in 379 accredited medical units in Mexico City, between February 24, 2020 and September 14, 2020.

| **Group** | **Total** | **Deaths** | **Survivors** | **Survival** | **p value** |
| --- | --- | --- | --- | --- | --- |
| Non-pregnant/puerperal adults | 129837 | 11569 | 118268 | 91.1% | - |
| Type of treatment | | | | | |
| No antiviral/antibiotic | 108213 | 7314 | 100899 | 93.2% | Reference |
| Antiviral only | 3990 | 1063 | 2927 | 73.4% | <0.0001 |
| Antibiotic only | 13300 | 1945 | 11355 | 85.4% | <0.0001 |
| Antiviral + Antibiotic | 4875 | 1306 | 3569 | 73.2% | <0.0001 |
| Type of antiviral or antibiotic only | | | | | |
| No antiviral/antibiotic | 108213 | 7314 | 100899 | 93.2% | Reference |
| Acyclovir | 36 | 4 | 32 | 88.9% | 0.4 |
| Amantadine | 308 | 15 | 293 | 95.1% | 0.2 |
| Lopinavir-Ritonavir | 99 | 25 | 74 | 74.7% | <0.0001 |
| Oseltamivir | 8324 | 2310 | 6014 | 72.2% | <0.0001 |
| Rimantadine | 59 | 7 | 52 | 88.1% | 0.1 |
| Zanamivir | 39 | 8 | 31 | 79.5% | 0.001 |
| Antibiotic only | 13300 | 1945 | 11355 | 85.4% | <0.0001 |
| Treatment with oseltamivir | | | | | |
| No antiviral / Antibiotic | 108213 | 7314 | 100899 | 93.2% | Reference |
| Oseltamivir | 3738 | 1032 | 2706 | 72.4% | <0.0001 |
| Antibiotic only | 13300 | 1945 | 11355 | 85.4% | <0.0001 |
| Oseltamivir + Antibiotic | 4586 | 1278 | 3308 | 72.1% | <0.0001 |
| Data expressed as number of patients or percentual of survival. P values were calculated by Log-Rank test. | | | | | |

Supplementary Table 7. Survival analysis of laboratory-confirmed COVID-19 children and adolescents treated with antivirals and/or antibiotics in 273 accredited medical units in Mexico City, between February 24, 2020 and September 14, 2020.

| **Group** | **Total** | **Deaths** | **Survivors** | **Survival** | **p value** |
| --- | --- | --- | --- | --- | --- |
| Children and adolescents | 5791 | 40 | 5751 | 99.31% | - |
| Type of treatment | | | | | |
| No antiviral/antibiotic | 5336 | 28 | 5308 | 99.5% | Reference |
| Antiviral only | 40 | 0 | 40 | 100.0% | 0.7 |
| Antibiotic only | 376 | 10 | 366 | 97.3% | <0.0001 |
| Antiviral + Antibiotic | 39 | 2 | 37 | 94.9% | <0.0001 |
| Type of antiviral or antibiotic only | | | | | |
| No antiviral/antibiotic | 5336 | 28 | 5308 | 99.5% | Reference |
| Amantadine | 9 | 0 | 9 | 100.0% | 0.8 |
| Lopinavir-Ritonavir | 1 | 0 | 1 | 100.0% | 0.9 |
| Oseltamivir | 67 | 2 | 65 | 97.0% | 0.008 |
| Rimantadine | 2 | 0 | 2 | 100.0% | 0.9 |
| Antibiotic only | 376 | 10 | 366 | 97.3% | <0.0001 |
| Treatment with oseltamivir | | | | | |
| No antiviral/antibiotic | 5336 | 28 | 5308 | 99.5% | Reference |
| Oseltamivir | 36 | 0 | 36 | 100.0% | 0.7 |
| Antibiotic only | 376 | 10 | 366 | 97.3% | <0.0001 |
| Oseltamivir + Antibiotic | 31 | 2 | 29 | 93.5% | <0.0001 |
| Data expressed as number of patients or percentual of survival. P values were calculated by Log-Rank test. | | | | | |

Supplementary Table 8. Survival analysis of laboratory-confirmed COVID-19 pregnant women treated with antivirals and/or antibiotics in 177 accredited medical units in Mexico City, between February 24, 2020 and September 14, 2020.

| **Group** | **Total** | **Deaths** | **Survivors** | **Survival** | **p value** |
| --- | --- | --- | --- | --- | --- |
| Pregnancy | 581 | 10 | 571 | 98.3% | - |
| Type of treatment | | | | | |
| No antiviral / Antibiotic | 530 | 8 | 522 | 98.5% | Reference |
| Antiviral only | 12 | 1 | 11 | 91.7% | 0.07 |
| Antibiotic only | 35 | 0 | 35 | 100.0% | 0.9 |
| Antiviral + Antibiotic | 6 | 1 | 5 | 83.3% | 0.004 |
| Type of antiviral | | | | | |
| No antiviral / Antibiotic | 530 | 8 | 522 | 98.5% | Reference |
| Amantadine | 2 | 0 | 2 | 100.0% | 0.9 |
| Oseltamivir | 16 | 2 | 14 | 87.5% | 0.001 |
| Antibiotic only | 35 | 0 | 35 | 100.0% | 0.5 |
| Treatment with Oseltamivir | | | | | |
| No antiviral / Antibiotic | 530 | 8 | 522 | 98.5% | Reference |
| Oseltamivir | 11 | 1 | 10 | 90.9% | 0.05 |
| Antibiotic only | 35 | 0 | 35 | 100.0% | 0.5 |
| Oseltamivir + Antibiotic | 5 | 1 | 4 | 80.0% | 0.001 |
| Data expressed as number of patients or percentual of survival. P values were calculated by Log-Rank test. | | | | | |

Supplementary Table 9. Unadjusted mortality risk in laboratory-confirmed COVID-19 patients receiving antivirals, antibiotics, both, or none in accredited COVID-19 medical units in Mexico City.

|  | All patients | | Ambulatory | | Hospitalized | | Non-Critical | | Critical | |
| --- | --- | --- | --- | --- | --- | --- | --- | --- | --- | --- |
|  | HR (95%CI) | p | HR (95%CI) | p | HR (95%CI) | p | HR (95%CI) | p | HR (95%CI) | p |
| **Models for type of treatment** | | | | | | | | | | |
| No antiviral / antibiotic | Reference | | Reference | | Reference | | Reference | | Reference | |
| Antiviral only | 4.43 (4.15-4.72) | <0.0001 | 7.48 (6.29-8.90) | <0.0001 | 1.19 (1.11-1.27) | <0.0001 | 5.07 (4.67-5.52) | <0.0001 | 1.21 (1.09-1.34) | <0.0001 |
| Antibiotic only | 2.15 (2.05-2.27) | <0.0001 | 1.09 (0.88-1.36) | 0.4 | 0.79 (0.75-0.83) | <0.0001 | 1.89 (1.75-2.03) | <0.0001 | 0.63 (0.58-0.67) | <0.0001 |
| Antiviral + antibiotic | 4.31 (4.05-4.58) | <0.0001 | 4.02 (3.09-5.22) | <0.0001 | 0.90 (0.85-0.96) | 0.001 | 4.65 (4.29-5.05) | <0.0001 | 0.96 (0.88-1.05) | 0.4 |
| **Models for type of antiviral** | | | | | | | | | | |
| No antiviral / antibiotic | Reference | | Reference | | Reference | | Reference | | Reference | |
| Acyclovir | 1.71 (0.64-4.56) | 0.3 | Not estimable | | 1.68 (0.63-4.48) | 0.3 | 1.51 (0.38-6.07) | 0.6 | 1.51 (0.38-6.03) | 0.6 |
| Amantadine | 0.71 (0.43-1.17) | 0.2 | 1.06 (0.34-3.29) | 0.9 | 0.77 (0.44-1.36) | 0.4 | 0.68 (0.34-1.36) | 0.3 | 0.85 (0.40-1.78) | 0.7 |
| Lopinavir-Ritonavir | 3.90 (2.61-5.83) | <0.0001 | 12.96 (1.83-92.15) | 0.01 | 0.57 (0.38-0.85) | .006 | 2.69 (1.28-5.65) | 0.009 | 0.57 (0.36-0.90) | 0.02 |
| Oseltamivir | 4.57 (4.36-4.79) | <0.0001 | 6.55 (5.63-7.63) | <0.0001 | 1.01 (0.97-1.07) | 0.6 | 5.12 (4.81-5.46) | <0.0001 | 1.07 (0.99-1.14) | 0.08 |
| Rimantadine | 1.74 (0.84-3.68) | 0.1 | 2.02 (0.28-14.36) | 0.5 | 1.19 (0.54-2.65) | 0.7 | 1.83 (0.69-4.89) | 0.2 | 1.77 (0.57-5.48) | 0.3 |
| Zanamivir | 3.22 (1.61-6.44) | 0.001 | 5.24 (0.73-37.23) | 0.09 | 0.76 (0.36-1.60) | 0.5 | 2.34 (0.76-7.27) | 0.1 | 1.18 (0.49-2.85) | 0.7 |
| Antibiotic only | 2.15 (2.05-2.27) | <0.0001 | 1.09 (0.88-1.36) | 0.4 | 0.79 (0.75-0.83) | <0.0001 | 1.89 (1.75-2.03) | <0.0001 | 0.63 (0.59-0.67) | <0.0001 |
| **Models for Acyclovir** | | | | | | | | | | |
| No antiviral / antibiotic | Reference | | Reference | | Reference | | Reference | | Reference | |
| Acyclovir only | 2.16 (0.30-15.31) | 0.4 | Not estimable | | 4.09 (0.58-29.05) | 0.2 | Not estimable | | 2.21 (0.31-15.68) | 0.4 |
| Antibiotic only | 2.15 (2.05-2.27) | <0.0001 | 1.09 (0.88-1.36) | 0.4 | 0.78 (0.75-0.83) | <0.0001 | 1.89 (1.75-2.03) | <0.0001 | 0.63 (0.58-0.67) | <0.0001 |
| Acyclovir + antibiotic | 1.60 (0.52-4.97) | 0.4 | Not estimable | | 1.41 (0.45-4.37) | 0.6 | 1.85 (0.46-7.38) | 0.4 | 1.16 (0.16-8.24) | 0.9 |
| **Models for Amantadine** | | | | | | | | | | |
| No antiviral / antibiotic | Reference | | Reference | | Reference | | Reference | | Reference | |
| Amantadine only | 1.17 (0.68-2.02) | 0.6 | 1.34 (0.34-5.38) | 0.7 | 1.47 (0.82-2.66) | 0.2 | 1.15 (0.55-2.42) | 0.7 | 1.12 (0.50-2.50) | 0.8 |
| Antibiotic only | 2.15 (2.05-2.27) | <0.0001 | 1.09 (0.88-1.36) | 0.4 | 0.79 (0.75-0.83) | <0.0001 | 1.89 (1.75-2.03) | <0.0001 | 0.63 (0.58-0.67) | <0.0001 |
| Amantadine + antibiotic | 0.19 (0.05-0.08) | 0.02 | 0.74 (0.10-5.27) | 0.8 | 0.13 (0.09-0.88) | 0.04 | 0.18 (0.03-1.25) | 0.08 | 0.34 (0.05-2.44) | 0.3 |
| **Models for Lopinavir-Ritonavir** | | | | | | | | | | |
| No antiviral / antibiotic | Reference | | Reference | | Reference | | Reference | | Reference | |
| Lopinavir-Ritonavir only | 2.79 (1.05-7.44) | 0.04 | 4.24.97 (59.62-3029.30) | <0.0001 | 0.38 (0.14-1.00) | 0.05 | 1.91 (0.27-13.63) | 0.5 | 0.36 (0.14-0.96) | 0.04 |
| Antibiotic only | 2.15 (2.05-2.27) | <0.0001 | 1.09 (0.88-1.36) | 0.4 | 0.79 (0.75-0.83) | <0.0001 | 1.89 (1.75-2.03) | <0.0001 | 0.63 (0.58-0.67) | <0.0001 |
| Lopinavir-Ritonavir + antibiotic | 4.24 (2.73-6.58) | <0.0001 | Not estimable | | 0.64 (0.41-0.99) | 0.04 | 2.88 (1.29-6.42) | 0.01 | 0.67 (0.39-1.14) | 0.1 |
| **Models for Oseltamivir** | | | | | | | | | | |
| No antiviral / antibiotic | Reference | | Reference | | Reference | | Reference | | Reference | |
| Oseltamivir only | 4.62 (4.33-4.94) | <0.0001 | 8.00 (6.71-9.54) | <0.0001 | 1.20 (1.12-1.29) | <0.0001 | 5.36 (4.92-5.83) | <0.0001 | 1.24 (1.12-1.37) | <0.0001 |
| Antibiotic only | 2.15 (2.05-2.27) | <0.0001 | 1.09 (0.88-1.36) | 0.4 | 0.79 (0.75-0.83) | <0.0001 | 1.89 (1.75-2.03) | <0.0001 | 0.63 (0.59-0.67) | <0.0001 |
| Oseltamivir + antibiotic | 4.53 (4.26-4.82) | <0.0001 | 4.56 (3.51-5.94) | <0.0001 | 0.91 (0.86-0.97) | 0.003 | 4.92 (4.54-5.35) | <0.0001 | 0.97 (0.89-1.06) | 0.5 |
| **Models for Rimantadine** | | | | | | | | | | |
| No antiviral / antibiotic | Reference | | Reference | | Reference | | Reference | | Reference | |
| Rimantadine only | 2.75 (1.24-6.12) | 0.01 | 4.09 (0.58-29.09) | 0.2 | 1.21 (0.50-2.90) | 0.7 | 2.54 (0.82-7.88) | 0.1 | 1.79 (0.58-5.55) | 0.3 |
| Antibiotic only | 2.15 (2.05-2.27) | <0.0001 | 1.09 (0.88-1.36) | 0.4 | 0.78 (0.75-0.83) | <0.0001 | 1.88 (1.75-2.03) | <0.0001 | 0.63 (0.58-0.67) | <0.0001 |
| Rimantadine + antibiotic | 0.55 (0.08-3.91) | 0.6 | Not estimable | | 1.14 (0.16-8.06) | 0.9 | 0.99 (0.14-7.08) | 0.9 | - | - |
| **Models for Zanamivir** | | | | | | | | | | |
| No antiviral / antibiotic | Reference | | Reference | | Reference | | Reference | | Reference | |
| Zanamivir only | 3.91 (1.75-8.69) | 0.001 | 10.31 (1.45-73.28) | 0.02 | 0.78 (0.33-1.88) | 0.6 | 1.40 (0.19-9.96) | 0.7 | 1.19 (0.49-2.86) | 0.7 |
| Antibiotic only | 2.15 (2.05-2.28) | <0.0001 | 1.09 (0.88-1.36) | 0.4 | 0.79 (0.75-0.83) | <0.0001 | 1.89 (1.75-2.03) | <0.0001 | 0.63 (0.49-2.86) | <0.0001 |
| Zanamivir + antibiotic | 2.11 (0.53-8.43) | 0.3 | Not estimable | | 0.72 (0.18-2.87) | 0.6 | 3.52 (0.88-14.1) | 0.07 | - | - |
| HR: Hazard ratio, 95%CI: 95% confidence intervals | | | | | | | | | | |

Supplementary Table 10. Mortality risk in laboratory-confirmed COVID-19 patients who underwent invasive mechanical ventilation (IMV) and were treated with antivirals and/or antibiotics in one of 197 accredited hospitals in Mexico City, between February 24, 2020 and September 14, 2020

|  | **Unadjusted model** | | **Adjusted model^a^** | |
| --- | --- | --- | --- | --- |
|  | **HR (95% CI)** | **p value** | **HR (95% CI)** | **p value** |
| **Type of treatment** | | | | |
| No antiviral/antibiotic | Reference | | Reference | |
| Antiviral only | 1.18 (1.07-1.31) | 0.001 | 1.13 (1.02-1.25) | 0.02 |
| Antibiotic only | 0.62 (0.59-0.67) | <0.0001 | 0.64 (0.60-0.69) | <0.0001 |
| Antiviral + antibiotic | 0.95 (0.88-1.04) | 0.3 | 0.98 (0.90-1.07) | 0.7 |
| **Type of antiviral** | | | | |
| No antiviral/antibiotic | Reference | | Reference | |
| Acyclovir | 1.42 (0.36-5.68) | 0.6 | 1.96 (0.49-7.83) | 0.3 |
| Amantadine | 1.29 (0.61-2.70) | 0.5 | 1.29 (0.61-2.72 | 0.5 |
| Lopinavir-Ritonavir | 0.67 (0.41-1.09) | 0.1 | 0.68 (0.42-1.12) | 0.1 |
| Oseltamivir | 1.04 (0.97-1.12) | 0.3 | 1.04 (0.97-1.12) | 0.2 |
| Rimantadine | 1.66 (0.53-5.14) | 0.4 | 1.57 (0.50-4.90) | 0.4 |
| Zanamivir | 1.09 (0.48-2.64) | 0.8 | 0.87 (0.36-2.09) | 0.8 |
| Antibiotic only | 0.63 (0.59-0.67) | <0.0001 | 0.64 (0.60-0.69) | <0.0001 |
| **Acyclovir** | | | | |
| No antiviral/antibiotic | Reference | | Reference | |
| Acyclovir only | 2.09 (0.29-14.9) | 0.5 | 2.85 (0.40-20.35) | 0.3 |
| Antibiotic only | 0.63 (0.58-0.67) | <0.0001 | 0.64 (0.60-0.69) | <0.0001 |
| Acyclovir + antibiotic | 1.09 (0.15-7.74) | 0.9 | 1.52 (0.21-10.78) | 0.7 |
| **Amantadine** | | | | |
| No antiviral/antibiotic | Reference | | Reference | |
| Amantadine only | 1.51 (0.68-3.35) | 0.3 | 1.50 (0.67-3.35) | 0.32 |
| Antibiotic only | 0.63 (0.58-0.67) | <0.0001 | 0.64 (0.6-0.69) | <0.0001 |
| Amantadine + antibiotic | 0.71 (0.09-5.01) | 0.7 | 0.71 (0.10-5.09) | 0.7 |
| **Lopinavir-Ritonavir** | | | | |
| No antiviral/antibiotic | Reference | | Reference | |
| Lopinavir-Ritonavir only | 0.59 (0.19-1.86) | 0.4 | 0.53 (0.17-1.63) | 0.3 |
| Antibiotic only | 0.63 (0.58-0.67) | <0.0001 | 0.64 (0.60-0.69) | <0.0001 |
| Lopinavir-Ritonavir + antibiotic | 0.68 (0.39-1.17) | 0.2 | 0.74 (0.43-1.27) | 0.3 |
| **Oseltamivir** | | | | |
| No antiviral/antibiotic | Reference | | Reference | |
| Oseltamivir only | 1.19 (1.07-1.31) | 0.001 | 1.14 (1.02-1.26) | 0.02 |
| Antibiotic only | 0.63 (0.59-0.67) | <0.0001 | 0.64 (0.60-0.69) | <0.0001 |
| Oseltamivir + antibiotic | 0.96 (0.88-1.05) | 0.4 | 0.99 (0.91-1.08) | 0.8 |
| **Rimantadine** | | | | |
| No antiviral/antibiotic | Reference | | Reference | |
| Rimantadine only | 1.68 (0.54-5.20) | 0.4 | 1.66 (0.53-5.17) | 0.4 |
| Antibiotic only | 0.63 (0.58-0.67) | <0.0001 | 0.64 (0.60-0.69) | <0.0001 |
| **Zanamivir** | | | | |
| No antiviral/antibiotic | Reference | | Reference | |
| Zanamivir only | 1.10 (0.46-2.66) | 0.8 | 0.87 (0.36-2.10) | .80 |
| Antibiotic only | 0.63 (0.58-0.67) | <0.0001 | 0.64 (0.60-0.69) | <0.0001 |
| HR: Hazard ratio, 95%CI: 95% confident interval.  a: Models adjusted by: Sex (men), Age, Diabetes, Chronic obstructive pulmonary disease, Immunosuppression, Hypertension, Cardiovascular disease, Chronic kidney disease, Smoker, Unemployed, Irritability, Headache, Myalgias, Arthralgias, Rhinorrhea, Abdominal pain, Cyanosis. | | | | |

Supplementary Table 11. Mortality risk in laboratory-confirmed COVID-19 patients who were treated with antivirals and/or antibiotics and admitted to one of 125 accredited intensive care units (ICU) in Mexico City, between February 24, 2020 and September 14, 2020

|  | **Unadjusted model** | | **Adjusted model^a^** | |
| --- | --- | --- | --- | --- |
|  | **HR (95% CI)** | **p value** | **HR (95% CI)** | **p value** |
| **Type of treatment** | | | | |
| No antiviral/antibiotic | Reference | | Reference | |
| Antiviral only | 1.60 (1.21-2.13) | 0.001 | 1.42 (1.07-1.89) | 0.02 |
| Antibiotic only | 1.15 (1.02-1.29) | 0.02 | 1.05 (0.93-1.19) | 0.4 |
| Antiviral + antibiotic | 1.71 (1.49-1.97) | <0.0001 | 1.49 (1.30-1.72) | <0.0001 |
| **Type of antiviral** | | | | |
| No antiviral/antibiotic | Reference | | Reference | |
| Acyclovir | 5.33 (0.75-37.9) | 0.09 | 8.38 (1.16-60.62) | 0.04 |
| Amantadine | Not estimable | | Not estimable | |
| Lopinavir-Ritonavir | 1.21 (0.75-1.97) | 0.44 | 0.99 (0.61-1.61) | 0.9 |
| Oseltamivir | 1.74 (1.52-1.96) | <0.0001 | 1.52 (1.33-1.74) | <0.0001 |
| Antibiotic only | 1.15 (1.02-1.29) | 0.02 | 1.05 (0.93-1.18) | 0.4 |
| **Acyclovir** | | | | |
| No antiviral/antibiotic | Reference | | Reference | |
| Acyclovir only | 5.46 (0.77-38.9) | 0.09 | 10.21 (1.40-74.38) | 0.02 |
| Antibiotic only | 1.15 (1.02-1.29) | 0.02 | 1.05 (0.93-1.18) | 0.4 |
| **Amantadine** | | | | |
| No antiviral/antibiotic | Reference | | Reference | |
| Amantadine only | Not estimable | | Not estimable | |
| Antibiotic only | 1.15 (1.02-1.29) | 0.02 | 1.05 (0.93-1.18) | 0.4 |
| Amantadine + antibiotic | Not estimable | | Not estimable | |
| **Lopinavir-Ritonavir** | | | | |
| No antiviral/antibiotic | Reference | | Reference | |
| Lopinavir-Ritonavir only | 0.74 0.28-1.98) | 0.6 | 0.50 (0.19-1.35) | 0.2 |
| Antibiotic only | 1.15 (1.02-1.29) | 0.02 | 1.05 (0.93-1.18) | 0.5 |
| Lopinavir-Ritonavir + antibiotic | 1.50 (0.89-2.61) | 0.2 | 1.32 (0.76-2.29) | 0.3 |
| **Oseltamivir** | | | | |
| No antiviral/antibiotic | Reference | | Reference | |
| Oseltamivir only | 1.79 (1.34-2.42) | <0.0001 | 1.64 (1.22-2.21) | <0.0001 |
| Antibiotic only | 1.15 (1.02-1.29) | 0.02 | 1.05 (0.93-1.19) | 0.4 |
| Oseltamivir + antibiotic | 1.73 (1.51-1.99) | <0.0001 | 1.50 (1.30-1.73) | <0.0001 |
| HR: Hazard ratio, 95%CI: 95% confident interval.  a: Models adjusted by: Sex (men), Age, Diabetes, Chronic obstructive pulmonary disease, Hypertension, Obesity, Chronic kidney disease, Unemployed, Fever, Cough, Shortness of breath, Myalgias, Vomit, Cyanosis | | | | |

Supplementary Table 12. Mortality risk in laboratory-confirmed COVID-19 non-pregnant/puerperal adults treated with antivirals and/or antibiotics in 379 accredited medical units in Mexico City, between February 24, 2020 and September 14, 2020.

|  | **Unadjusted model** | | **Adjusted model^a^** | |
| --- | --- | --- | --- | --- |
|  | **HR (95% CI)** | **p value** | **HR (95% CI)** | **p value** |
| **Type of treatment** | | | | |
| No antiviral/antibiotic | Reference | | Reference | |
| Antiviral only | 4.34 4.07-4.62) | <0.0001 | 1.80 (1.68-1.92) | <0.0001 |
| Antibiotic only | 2.18 (2.08-2.29) | <0.0001 | 1.19 (1.13-1.25) | <0.0001 |
| Antiviral + antibiotic | 4.31 (4.07-4.57) | <0.0001 | 1.64 (1.54-1.74) | <0.0001 |
| **Type of antiviral** | | | | |
| No antiviral/antibiotic | Reference | | Reference | |
| Acyclovir | 1.57 0.59-4.18) | 0.4 | 1.40 (0.52-3.72) | 0.5 |
| Amantadine | 0.69 (0.42-1.16) | 0.2 | 0.72 (0.43-1.20) | 0.2 |
| Lopinavir-Ritonavir | 3.88 (2.62-5.74) | <0.0001 | 1.14 (0.77-1.69) | 0.5 |
| Oseltamivir | 4.52 (4.32-4.74) | <0.0001 | 1.73 (1.65-1.82) | <0.0001 |
| Rimantadine | 1.75 (0.83-3.67) | 0.1 | 1.47 (0.70-3.08) | 0.3 |
| Zanamivir | 3.04 (1.52-6.08) | 0.002 | 1.61 (0.81-3.22) | 0.2 |
| Antibiotic only | 2.18 (2.08-2.29) | <0.0001 | 1.19 (1.13-1.25) | <0.0001 |
| **Acyclovir** | | | | |
| No antiviral/antibiotic | Reference | | Reference | |
| Acyclovir only | 2.09 (0.29-14.8) | 0.5 | 7.50 (1.06-53.27) | 0.04 |
| Antibiotic only | 2.18 (2.08-2.29) | <0.0001 | 1.16 (1.10-1.22) | <0.0001 |
| Acyclovir + antibiotic | 1.45 (0.46-4.48) | 0.5 | 1.11 (0.36-3.45) | 0.9 |
| **Amantadine** | | | | |
| No antiviral/antibiotic | Reference | | Reference | |
| Amantadine only | 1.15 (0.67-1.98) | 0.6 | 1.75 (1.02-3.02) | 0.04 |
| Antibiotic only | 2.18 (2.08-2.29) | <0.0001 | 1.16 (1.10-1.22) | <0.0001 |
| Amantadine + antibiotic | 0.19 (0.05-0.78) | 0.02 | 0.15 (0.04-0.59) | 0.007 |
| **Lopinavir-Ritonavir** | | | | |
| No antiviral/antibiotic | Reference | | Reference | |
| Lopinavir-Ritonavir only | 3.27 (1.36-7.88) | 0.008 | 0.83 (0.34-1.99) | 0.7 |
| Antibiotic only | 2.18 (2.08-2.29) | <0.0001 | 1.16 (1.11-1.22) | <0.0001 |
| Lopinavir-Ritonavir + antibiotic | 4.06 (2.62-6.29) | <0.0001 | 1.18 (0.76-1.83) | 0.5 |
| **Oseltamivir** | | | | |
| No antiviral/antibiotic | Reference | | Reference | |
| Oseltamivir only | 4.52 (4.24-4.83) | <0.0001 | 1.81 (1.69-1.93) | <0.0001 |
| Antibiotic only | 2.18 (2.08-2.29) | <0.0001 | 1.19 (1.13-1.25) | <0.0001 |
| Oseltamivir + antibiotic | 4.52 (4.26-4.79) | <0.0001 | 1.68 (1.58-1.78) | <0.0001 |
| **Rimantadine** | | | | |
| No antiviral/antibiotic | Reference | | Reference | |
| Rimantadine only | 2.71 (1.22-6.03) | 0.02 | 2.02 (0.91-4.50) | 0.1 |
| Antibiotic only | 2.18 (2.08-2.29) | <0.0001 | 1.16 (1.11-1.22) | <0.0001 |
| Rimantadine + antibiotic | 0.56 (0.08-3.97) | 0.6 | 0.53 (0.07-3.77) | 0.5 |
| **Zanamivir** | | | | |
| No antiviral/antibiotic | Reference | | Reference | |
| Zanamivir only | 3.79 (1.71-8.45) | 0.001 | 2.02 (0.91-4.50) | 0.1 |
| Antibiotic only | 2.18 (2.08-2.29) | <0.0001 | 1.16 (1.11-1.22) | <0.0001 |
| Zanamivir + antibiotic | 1.90 (0.48-7.62) | 0.36 | 0.97 (0.24-3.86) | 0.9 |
| HR: Hazard ratio, 95%CI: 95% confident interval.  a: Models adjusted by: Sex (men), Age, Diabetes, Chronic obstructive pulmonary disease, Immunosuppression, Hypertension, Human immunodeficiency virus/acquired immune deficiency syndrome, Obesity, Cardiovascular disease, Chronic kidney disease, Smoker, Unemployed, Fever, Cough, Shortness of breath, Irritability, Chest pain, Myalgias, Arthralgias, Polypnea, Vomit, Abdominal pain. | | | | |

Supplementary Table 13. Mortality risk in laboratory-confirmed COVID-19 children and adolescents treated with antivirals and/or antibiotics in 273 accredited medical units in Mexico City, between February 24, 2020 and September 14, 2020

|  | **Unadjusted model** | | **Adjusted model^a^** | |
| --- | --- | --- | --- | --- |
|  | **HR (95% CI)** | **p value** | **HR (95% CI)** | **p value** |
| **Type of treatment** | | | | |
| No antiviral/antibiotic | Reference | | Reference | |
| Antiviral only | Not estimable | | Not estimable | |
| Antibiotic only | 5.01 (2.43-10.32) | <0.0001 | 4.22 (2.01-8.86) | <0.0001 |
| Antiviral + antibiotic | 9.68 (2.31-40.62) | 0.002 | 6.54 (1.44-29.63) | 0.01 |
| **Type of antiviral** | | | | |
| No antiviral/antibiotic | Reference | | Reference | |
| Amantadine | Not estimable | | Not estimable | |
| Lopinavir-Ritonavir | Not estimable | | Not estimable | |
| Oseltamivir | 5.66 (1.35-23.73) | 0.02 | 4.22 (0.94-18.99) | 0.06 |
| Rimantadine | Not estimable | | Not estimable | |
| Antibiotic only | 5.01 (2.43-10.31) | <0.0001 | 4.20 (2.00-8.83) | <0.0001 |
| **Oseltamivir** | | | | |
| No antiviral/antibiotic | Reference | | Reference | |
| Oseltamivir only | Not estimable | | Not estimable | |
| Antibiotic only | 5.01 (2.43-10.3) | <0.0001 | 4.20 (2.00-8.82) | <0.0001 |
| Oseltamivir + antibiotic | 12.10 (2.89-50.84) | 0.001 | 7.95 (1.70-37.21) | 0.01 |
| HR: Hazard ratio, 95%CI: 95% confident interval.  a: Models adjusted by: Diabetes, Immunosuppression, Chronic kidney disease. | | | | |

Supplementary Table 14. Mortality risk in laboratory-confirmed COVID-19 pregnant women treated with antivirals and/or antibiotics in 117 accredited medical units in Mexico City, between February 24, 2020 and September 14, 2020.

|  | **Unadjusted model** | |
| --- | --- | --- |
|  | **HR (95% CI)** | **P value** |
| **Type of treatment** | | |
| No antiviral/antibiotic | Reference | |
| Antiviral only | 5.51 (0.69-44.06) | 0.1 |
| Antibiotic only | Not estimable | |
| Antiviral + antibiotic | 11.14 (1.39-89.07) | 0.02 |
| **Type of antiviral** | | |
| No antiviral/antibiotic | Reference | |
| Amantadine | Not estimable | |
| Oseltamivir | 8.35 (1.77-39.30) | 0.007 |
| Antibiotic only | Not estimable | |
| **Oseltamivir** | | |
| No antiviral/antibiotic | Reference | |
| Oseltamivir only | 6.04 (0.76-48.26) | 0.09 |
| Antibiotic only | Not estimable | |
| Oseltamivir + antibiotic | 13.52 (1.69-108.10) | 0.01 |
| HR: Hazard ratio, 95%CI: 95% confident interval. | | |

Supplementary Table 15. E-Values for Cox regression models in the general population, ambulatory, hospitalized, non-critical, and critical COVID-19 patients.

|  | All patients | Ambulatory | Hospitalized | Non-Critical | Critical |
| --- | --- | --- | --- | --- | --- |
| **Models for type of treatment** | | | | | |
| Antiviral only | 2.83 (2.60) | 8.87 (7.34) | - | 3.48 (3.12) | - |
| antibiotic only | 1.51 (1.37) | 2.12 (1.5) | 1.77 (1.60) | - | 2.35 (2.12) |
| Antiviral + antibiotic | 2.52 (2.30) | 3.23 (2.3) | 1.43 (1.21) | 2.64 (2.34) | - |
| **Models for type of antiviral** | | | | | |
| Acyclovir | - | - | 4.94 (1.21) | - | - |
| Amantadine | - | - | - | - | - |
| Lopinavir-Ritonavir | - | - | 2.78 (1.5) | - | - |
| Oseltamivir | 2.71 (2.54) | 6.50 (5.47) | - | 3.08 (2.83) | - |
| Rimantadine | - | - | - | - | - |
| Zanamivir | - | - | - | - | - |
| Antibiotic only | 1.54 (1.37) | 2.12 (1.46) | 1.77 (1.60) | - | 2.30 (2.12) |
| **Models for Acyclovir** | | | | | |
| Acyclovir only | 15.68 (1.54) | - | 17.45 (1.83) | - | - |
| Antibiotic only | 1.46 (1.28) | 2.17 (1.5) | 1.74 (1.60) | - | 2.35 (2.12) |
| Acyclovir + antibiotic | - | - | - | - | - |
| **Models for Amantadine** | | | | | |
| Amantadine only | 2.96 (1.21) | - | - | - | - |
| Antibiotic only | 1.46 (1.28) | 2.17 (1.50) | 1.74 (1.60) | - | 2.35 (2.12) |
| Amantadine + antibiotic | 12.81 (2.78) | - | - | 14.87 (1.46) | - |
| **Models for Lopinavir-Ritonavir** | | | | | |
| Lopinavir-Ritonavir only | - | 113 (15.22) | - | - | - |
| Antibiotic only | 1.46 (1.28) | 2.17 (1.50) | 1.74 (1.60) | - | 2.35 (2.12) |
| Lopinavir-Ritonavir + antibiotic | - | - | - | - | - |
| **Models for Oseltamivir** | | | | | |
| Oseltamivir only | 2.83 (2.60) | 9.05 (7.48) | - | 3.52 (3.17) | - |
| Antibiotic only | 1.51 (1.37) | 2.12 (1.50) | 1.74 (1.60) | - | 2.35 (2.12) |
| Oseltamivir + antibiotic | 2.60 (2.39) | 3.62 (2.69) | 1.39 (1.16) | 2.75 (2.47) | - |
| **Models for Rimantadine** | | | | | |
| Rimantadine only | - | - | - | - | - |
| Antibiotic only | 1.46 (1.28) | 2.17 (1.50) | 1.74 (1.60) | - | 2.35 (2.12) |
| Rimantadine + antibiotic | - | - | - | - | - |
| **Models for Zanamivir** | | | | | |
| Zanamivir only | - | - | - | - | - |
| Antibiotic only | 1.46 (1.28) | 2.17 (1.50) | 1.74 (1.60) | - | 2.35 (2.12) |
| Zanamivir + antibiotic | - | - | - | - | - |
| Data presented as E-value for point estimate (E-value for lower-limit confidence interval) | | | | | |

Supplementary Table 16. E-Values for Cox regression models in non-pregnant/puerperal adults, children and adolescents, VMI, and ICU patients

|  | IMV | ICU | Non-pregnant/puerperal adults | Children and adolescents |
| --- | --- | --- | --- | --- |
| **Models for type of treatment** | | | | |
| Antiviral only | 1.51 (1.16) | 2.19 (1.34) | 3.00 (2.75) | - |
| Antibiotic only | 2.50 (2.26) | - | 1.67 (1.51) | 7.91 (3.43) |
| Antiviral + antibiotic | - | 2.34 (1.92) | 2.66 (2.45) | 12.56 (2.24) |
| **Models for type of antiviral** | | | | |
| Acyclovir | - | 16.24 (1.59) | - | - |
| Amantadine | - | - | - | - |
| Lopinavir-Ritonavir | - | - | - | - |
| Oseltamivir | - | 2.41 (1.99) | 2.85 (2.69) | - |
| Rimantadine | - | - | - | - |
| Zanamivir | - | - | - | - |
| Antibiotic only | 2.50 (2.26) | - | 1.67 (1.51) | 7.87 (3.41) |
| **Models for Acyclovir** | | | | |
| Acyclovir only | - | 19.91 (2.15) | 14.48 (1.31) | - |
| Antibiotic only | 2.50 (2.26) | - | 1.59 (1.43) | - |
| Acyclovir + antibiotic | - | - | - | - |
| **Models for Amantadine** | | | | |
| Amantadine only | - | - | 2.9 (1.16) | - |
| Antibiotic only | 2.50 (2.26) | - | 1.59 (1.43) | - |
| Amantadine + antibiotic | - | - | - | - |
| **Models for Lopinavir-Ritonavir** | | | | |
| Lopinavir-Ritonavir only | - | - | - | - |
| Antibiotic only | 2.50 (2.26) | - | 1.59 (1.43) | - |
| Lopinavir-Ritonavir + antibiotic | - | - | - | - |
| **Models for Oseltamivir** | | | | |
| Oseltamivir only | 1.54 (1.16) | 2.66 (1.74) | 3.02 (2.77) | - |
| Antibiotic only | 2.50 (2.26) | - | 1.67 (1.51) | 7.87 (3.41) |
| Oseltamivir + antibiotic | - | 2.37 (1.92) | 2.75 (2.54) | 15.38 (2.79) |
| Data presented as E-value for point estimate (E-value for lower-limit confidence interval) | | | | |

Supplementary Table 17. Baseline characteristics of patients with COVID-19 treated with antivirals and propensity score-matched controls not treated with antiviral sor antibiotics.

|  | Total  n=8088 | Antiviral  n=4044 | Control  n=4044 |
| --- | --- | --- | --- |
| Sex |  | | |
| Women | 3634 (44.9) | 1821 45.0 | 1813 (44.8) |
| Men | 4454 (55.1) | 2223 (55.0) | 2231 (55.2) |
| Age, mean (SD) | 50.5 (16.5) | 50.5 (16.5) | 50.5 (16.5) |
| Age categories |  |  |  |
| 0-19 years | 114 (1.4) | 57 (1.4) | 57 (1.4) |
| 20-29 years | 747 (9.2) | 372 (9.2) | 375 (9.3) |
| 30-39 years | 1411 (17.4) | 704 (17.4) | 707 (17.5) |
| 40-49 years | 1667 (20.6) | 830 (20.5) | 837 (20.7) |
| 50-59 years | 1724 (21.3) | 872 (21.6) | 852 (21.1) |
| 60-69 years | 1264 (15.6) | 632 (15.6) | 632 (15.6) |
| 70-79 years | 821 (10.2) | 403 (10.0) | 418 (10.3) |
| 80-89 years | 302 (3.7) | 156 (3.9) | 146 (3.6) |
| 90-99 years | 38 (0.5) | 18 (0.4) | 20 (0.5) |
| ≥100 years | 0 (0.0) | 0 (0.0) | 0 (0.0) |
| Indigenous self-identification | 58 (0.7) | 21 (0.5) | 37 (0.9) |
| Occupation |  | | |
| Technical services | 133 (1.6) | 63 (1.6) | 70 (1.7) |
| Education | 236 (2.9) | 131 (3.2) | 105 (2.6) |
| Healthcare | 1114 (13.8) | 459 (11.4) | 655 (16.2) |
| Agricultural activities | 15 (0.2) | 10 (0.2) | 5 (0.1) |
| Commerce | 2532 (31.3) | 1454 (36.0) | 1078 (26.7) |
| Other | 1618 (20.0) | 707 (17.5) | 911 (22.5) |
| Unemployed | 483 (6.0) | 206 (5.1) | 277 (6.8) |
| Stay-at-home | 1957 (24.2) | 1014 (25.1) | 943 (23.3) |
| Last-season flu vaccination | 1396 (17.3) | 701 (17.3) | 695 (17.2) |
| Comorbidities |  | | |
| Diabetes | 1783 (22.0) | 873 (21.6) | 910 (22.5) |
| COPD | 239 (3.0) | 120 (3.0) | 119 (2.9) |
| Asthma | 181 (2.2) | 85 (2.1) | 96 (2.4) |
| Immunosuppression | 145 (1.8) | 61 (1.5) | 84 (2.1) |
| Hypertension | 2144 (26.5) | 1070 (26.5) | 1074 (26.6) |
| HIV/AIDS | 56 (0.7) | 26 (0.6) | 30 (0.7) |
| Cardiovascular disease | 276 (3.4) | 123 (3.0) | 153 (3.8) |
| Obesity | 1716 (21.2) | 879 (21.7) | 837 (20.7) |
| Chronic kidney disease | 266 (3.3) | 116 (2.9) | 150 (3.7) |
| Smoker | 924 (11.4) | 463 (11.4) | 461 (11.4) |
| Type of medical attention |  | | |
| Ambulatory | 4794 (59.3) | 2782 (68.8) | 2012 (49.8) |
| Hospitalization | 3294 (40.7) | 1262 (31.2) | 2032 (50.2) |
| Severity of the disease |  | | |
| Non-critical | 7232 (89.4) | 3714 (91.8) | 3518 (87.0) |
| Critical | 856 (10.6) | 330 (8.2) | 526 (13.0) |
| Time from symptom onset to medical attention | 4.7 (3.9) | 4.5 (3.9) | 4.9 (3.9) |
| Baseline symptoms |  | | |
| Fever | 6706 (82.9) | 3374 (83.4) | 3332 (82.4) |
| Cough | 6646 (82.2) | 3240 (80.1) | 3406 (84.2) |
| Sore throat | 3985 (49.3) | 1971 (48.7) | 2014 (49.8) |
| Shortness of breath | 4448 (55.0) | 2214 (54.7) | 2234 (55.2) |
| Irritability | 1947 (24.1) | 868 (21.5) | 1079 (26.7) |
| Diarrhea | 2292 (28.3) | 1140 (28.2) | 1152 (28.5) |
| Chest pain | 3122 (38.6) | 1460 (36.1) | 1662 (41.1) |
| Chills | 4006 (49.5) | 1868 (46.2) | 2138 (52.9) |
| Headache | 6422 (79.4) | 3138 (77.6) | 3284 (81.2) |
| Myalgias | 5306 (65.6) | 2673 (66.1) | 2633 (65.1) |
| Arthralgias | 4810 (59.5) | 2433 (60.2) | 2377 (58.8) |
| Abrupt deterioration | 4995 (61.8) | 2291 (56.7) | 2704 (66.9) |
| Rhinorrhea | 2599 (32.1) | 1249 (30.9) | 1350 (33.4) |
| Polypnea | 1830 (22.6) | 749 (18.5) | 1081 (26.7) |
| Vomit | 888 (11.0) | 409 (10.1) | 479 (11.8) |
| Abdominal pain | 1693 (20.9) | 668 (16.5) | 1025 (25.3) |
| Conjunctivitis | 1116 (13.8) | 603 (14.9) | 513 (12.7) |
| Cyanosis | 719 (8.9) | 256 (6.3) | 463 (11.4) |
| Sudden onset of symptoms | 3191 (39.5) | 1574 (38.9) | 1617 (40.0) |
| Data expressed as Frequency (%) or mean (SD)  SD: Standard deviation, COPD: Chronic obstructive pulmonary disease, HIV/AIDS: Human immunodeficiency virus/acquired immune deficiency syndrome. | | | |

Supplementary Table 18. Baseline characteristics of patients with COVID-19 treated with Oseltamivir and propensity score-matched controls not treated with antiviral sor antibiotics.

|  | Total  n=7574 | Oseltamivir  n=3787 | Control  n=3787 |
| --- | --- | --- | --- |
| Sex |  | | |
| Women | 3351 (44.2) | 1676 (44.3) | 1675 (44.2) |
| Men | 4223 (55.8) | 2111 (55.7) | 2112 (55.8) |
| Age, mean (SD) | 50.8 (16.5) | 50.8 (16.6) | 50.7 (16.4) |
| Age categories |  |  |  |
| 0-19 years | 98 (1.3) | 47 (1.2) | 51 (1.3) |
| 20-29 years | 698 (9.2) | 349 (9.2) | 349 (9.2) |
| 30-39 years | 1267 (16.7) | 632 (16.7) | 635 (16.8) |
| 40-49 years | 1569 (20.7) | 788 (20.8) | 781 (20.6) |
| 50-59 years | 1624 (21.4) | 822 (21.7) | 802 (21.2) |
| 60-69 years | 1213 (16.0) | 607 (16.0) | 606 (16.0) |
| 70-79 years | 785 (10.4) | 386 (10.2) | 399 (10.5) |
| 80-89 years | 282 (3.7) | 138 (3.6) | 144 (3.8) |
| 90-99 years | 38 (0.5) | 18 (0.5) | 20 (0.5) |
| ≥100 years | 0 (0.0) | 0 (0.0) | 0 (0.0) |
| Indigenous self-identification | 51 (0.7) | 17 (0.4) | 34 (0.9) |
| Occupation |  | | |
| Technical services | 127 (1.7) | 58 (1.5) | 69 (1.8) |
| Education | 202 (2.7) | 112 (3.0) | 90 (2.4) |
| Healthcare | 1027 (13.6) | 408 (10.8) | 619 (16.3) |
| Agricultural activities | 17 (0.2) | 12 (0.3) | 5 (0.1) |
| Commerce | 2388 (31.5) | 1394 (36.8) | 994 (26.2) |
| Other | 1484 (19.6) | 626 (16.5) | 858 (22.7) |
| Unemployed | 464 (6.1) | 204 (5.4) | 260 (6.9) |
| Stay-at-home | 1865 (24.6) | 973 (25.7) | 892 (23.6) |
| Last-season flu vaccination | 1297 (17.1) | 660 (17.4) | 637 (16.8) |
| Comorbidities |  | | |
| Diabetes | 1730 (22.8) | 853 (22.5) | 877 (23.2) |
| COPD | 219 (2.9) | 102 (2.7) | 117 (3.1) |
| Asthma | 177 (2.3) | 88 (2.3) | 89 (2.4) |
| Immunosuppression | 142 (1.9) | 61 (1.6) | 81 (2.1) |
| Hypertension | 2070 (27.3) | 1031 (27.2) | 1039 (27.4) |
| HIV/AIDS | 47 (0.6) | 17 (0.4) | 30 (0.8) |
| Cardiovascular disease | 263 (3.5) | 114 (3.0) | 149 (3.9) |
| Obesity | 1583 (20.9) | 796 (21.0) | 787 (20.8) |
| Chronic kidney disease | 259 (3.4) | 113 (3.0) | 146 (3.9) |
| Smoker | 864 (11.4) | 436 (11.5) | 428 (11.3) |
| Type of medical attention |  | | |
| Ambulatory | 4412 (58.3) | 2588 (68.3) | 1824 (48.2) |
| Hospitalization | 3162 (41.7) | 1199 (31.7) | 1963 (51.8) |
| Severity of the disease |  | | |
| Non-critical | 6760 (89.3) | 3474 (91.7) | 3286 (86.8) |
| Critical | 814 (10.7) | 313 (8.3) | 501 (13.2) |
| Time from symptom onset to medical attention | 4.7 (3.9) | 4.4 (3.9) | 4.9 (4) |
| Baseline symptoms |  | | |
| Fever | 6370 (84.1) | 3204 (84.6) | 3166 (83.6) |
| Cough | 6220 (82.1) | 3013 (79.6) | 3207 (84.7) |
| Sore throat | 3719 (49.1) | 1831 (48.3) | 1888 (49.9) |
| Shortness of breath | 4286 (56.6) | 2137 (56.4) | 2149 (56.7) |
| Irritability | 1895 (25.0) | 882 (23.3) | 1013 (26.7) |
| Diarrhea | 2127 (28.1) | 1043 (27.5) | 1084 (28.6) |
| Chest pain | 2975 (39.3) | 1388 (36.7) | 1587 (41.9) |
| Chills | 3768 (49.7) | 1758 (46.4) | 2010 (53.1) |
| Headache | 6046 (79.8) | 2947 (77.8) | 3099 (81.8) |
| Myalgias | 4970 (65.6) | 2508 (66.2) | 2462 (65.0) |
| Arthralgias | 4506 (59.5) | 2273 (60.0) | 2233 (59.0) |
| Abrupt deterioration | 4746 (62.7) | 2187 (57.8) | 2559 (67.6) |
| Rhinorrhea | 2450 (32.3) | 1198 (31.6) | 1252 (33.1) |
| Polypnea | 1834 (24.2) | 790 (20.9) | 1044 (27.6) |
| Vomit | 853 (11.3) | 395 (10.4) | 458 (12.1) |
| Abdominal pain | 1585 (20.9) | 608 (16.1) | 977 (25.8) |
| Conjunctivitis | 1014 (13.4) | 540 (14.3) | 474 (12.5) |
| Cyanosis | 745 (9.8) | 291 (7.7) | 454 (12.0) |
| Sudden onset of symptoms | 3024 (39.9) | 1494 (39.5) | 1530 (40.4) |
| Data expressed as Frequency (%) or mean (SD)  SD: Standard deviation, COPD: Chronic obstructive pulmonary disease, HIV/AIDS: Human immunodeficiency virus/acquired immune deficiency syndrome. | | | |

Supplementary Table 19. Baseline characteristics of patients with COVID-19 treated with antibiotics and propensity score-matched controls not treated with antiviral sor antibiotics..

|  | Total  n=27486 | Antibiotic  n=13743 | Control  n=13743 |
| --- | --- | --- | --- |
| Sex |  | | |
| Women | 11940 (43.4) | 5940 (43.2) | 6000 (43.7) |
| Men | 15546 (56.6) | 7803 (56.8) | 7743 (56.3) |
| Age, mean (SD) | 48.2 (16.7) | 48.3 (16.8) | 48.1 (16.6) |
| Age categories |  |  |  |
| 0-19 years | 928 (3.4) | 445 (3.2) | 483 (3.5) |
| 20-29 years | 2778 (10.1) | 1399 (10.2) | 1379 (10.0) |
| 30-39 years | 4912 (17.9) | 2475 (18.0) | 2437 (17.7) |
| 40-49 years | 5976 (21.7) | 3010 (21.9) | 2966 (21.6) |
| 50-59 years | 5830 (21.2) | 2924 (21.3) | 2906 (21.1) |
| 60-69 years | 4104 (14.9) | 2056 (15.0) | 2048 (14.9) |
| 70-79 years | 2097 (7.6) | 1025 (7.5) | 1072 (7.8) |
| 80-89 years | 744 (2.7) | 355 (2.6) | 389 (2.8) |
| 90-99 years | 111 (0.4) | 52 (0.4) | 59 (0.4) |
| ≥100 years | 6 (0.0) | 2 (0.0) | 4 (0.0) |
| Indigenous self-identification | 152 (0.6) | 60 (0.4) | 92 (0.7) |
| Occupation |  | | |
| Technical services | 466 (1.7) | 229 (1.7) | 237 (1.7) |
| Education | 1284 (4.7) | 622 (4.5) | 662 (4.8) |
| Healthcare | 2829 (10.3) | 1555 (11.3) | 1274 (9.3) |
| Agricultural activities | 90 (0.3) | 39 (0.3) | 51 (0.4) |
| Commerce | 10282 (37.4) | 5227 (38.0) | 5055 (36.8) |
| Other | 4993 (18.2) | 2394 (17.4) | 2599 (18.9) |
| Unemployed | 1131 (4.1) | 674 (4.9) | 457 (3.3) |
| Stay-at-home | 6411 (23.3) | 3003 (21.9) | 3408 (24.8) |
| Last-season flu vaccination | 5289 (19.2) | 2538 (18.5) | 2751 (20.0) |
| Comorbidities |  | | |
| Diabetes | 5268 (19.2) | 2591 (18.9) | 2677 (19.5) |
| COPD | 480 (1.7) | 232 (1.7) | 248 (1.8) |
| Asthma | 622 (2.3) | 334 (2.4) | 288 (2.1) |
| Immunosuppression | 406 (1.5) | 171 (1.2) | 235 (1.7) |
| Hypertension | 6049 (22.0) | 2976 (21.7) | 3073 (22.4) |
| HIV/AIDS | 120 (0.4) | 58 (0.4) | 62 (0.5) |
| Cardiovascular disease | 659 (2.4) | 289 (2.1) | 370 (2.7) |
| Obesity | 6009 (21.9) | 3026 (22.0) | 2983 (21.7) |
| Chronic kidney disease | 531 (1.9) | 231 (1.7) | 300 (2.2) |
| Smoker | 3115 (11.3) | 1554 (11.3) | 1561 (11.4) |
| Type of medical attention |  |  |  |
| Ambulatory | 18548 (67.5) | 10280 (74.8) | 8268 (60.2) |
| Hospitalization | 8938 (32.5) | 3463 (25.2) | 5475 (39.8) |
| Severity of the disease |  |  |  |
| Non-critical | 24844 (90.4) | 12826 (93.3) | 12018 (87.4) |
| Critical | 2642 (9.6) | 917 (6.7) | 1725 (12.6) |
| Time from symptom onset to medical attention | 5.1 (3.9) | 5.5 (3.8) | 4.8 (3.9) |
| Baseline symptoms |  | | |
| Fever | 19681 (71.6) | 9912 (72.1) | 9769 (71.1) |
| Cough | 20783 (75.6) | 10412 (75.8) | 10371 (75.5) |
| Sore throat | 12673 (46.1) | 6432 (46.8) | 6241 (45.4) |
| Shortness of breath | 12972 (47.2) | 6471 (47.1) | 6501 (47.3) |
| Irritability | 5405 (19.7) | 2880 (21.0) | 2525 (18.4) |
| Diarrhea | 7083 (25.8) | 3589 (26.1) | 3494 (25.4) |
| Chest pain | 8782 (32.0) | 4613 (33.6) | 4169 (30.3) |
| Chills | 10668 (38.8) | 5665 (41.2) | 5003 (36.4) |
| Headache | 19521 (71.0) | 10121 (73.6) | 9400 (68.4) |
| Myalgias | 16206 (59.0) | 8161 (59.4) | 8045 (58.5) |
| Arthralgias | 14959 (54.4) | 7530 (54.8) | 7429 (54.1) |
| Abrupt deterioration | 14853 (54.0) | 7071 (51.5) | 7782 (56.6) |
| Rhinorrhea | 7814 (28.4) | 4126 (30.0) | 3688 (26.8) |
| Polypnea | 4169 (15.2) | 2200 (16.0) | 1969 (14.3) |
| Vomit | 2353 (8.6) | 1237 (9.0) | 1116 (8.1) |
| Abdominal pain | 3643 (13.3) | 2034 (14.8) | 1609 (11.7) |
| Conjunctivitis | 3532 (12.9) | 1913 (13.9) | 1619 (11.8) |
| Cyanosis | 1420 (5.2) | 808 (5.9) | 612 (4.5) |
| Sudden onset of symptoms | 10253 (37.3) | 5089 (37.0) | 5164 (37.6) |
| Data expressed as Frequency (%) or mean (SD)  SD: Standard deviation, COPD: Chronic obstructive pulmonary disease, HIV/AIDS: Human immunodeficiency virus/acquired immune deficiency syndrome. | | | |

Supplementary Table 20. Baseline characteristics of 903 patients with data of timing of initiation of antiviral treatment with laboratory-confirmed COVID-19, who received medical attention in one of 154 accredited medical units in Mexico City.

|  | **Total**  **n=903** | **Acyclovir**  **n=26** | **Amantadine**  **n=131** | **Lopinavir-Ritonavir**  **n=9** | **Oseltamivir**  **n=702** | **Rimantadine**  **n=24** | **Zanamivir**  **n=11** |
| --- | --- | --- | --- | --- | --- | --- | --- |
| Sex |  |  |  |  |  |  |  |
| Women | 403 (44.6) | 14 (53.8) | 72 (55) | 2 (22.2) | 294 (41.9) | 13 (54.2) | 8 (72.7) |
| Men | 500 (55.4) | 12 (46.2) | 59 (45) | 7 (77.8) | 408 (58.1) | 11 (45.8) | 3 (27.3) |
| Age, mean (SD) | 46.7 (14.8) | 44.5 (13.9) | 42.3 (14.3) | 51 (11.1) | 47.8 (14.8) | 42.6 (13.9) | 40 (13.9) |
| Age categories |  |  |  |  |  |  |  |
| 0-19 years | 17 (1.9) | 0 (0) | 5 (3.8) | 0 (0) | 11 (1.6) | 1 (4.2) | 0 (0) |
| 20-29 years | 88 (9.7) | 5 (19.2) | 18 (13.7) | 0 (0) | 62 (8.8) | 1 (4.2) | 2 (18.2) |
| 30-39 years | 204 (22.6) | 5 (19.2) | 40 (30.5) | 0 (0) | 147 (20.9) | 8 (33.3) | 4 (36.4) |
| 40-49 years | 220 (24.4) | 7 (26.9) | 26 (19.8) | 5 (55.6) | 170 (24.2) | 10 (41.7) | 2 (18.2) |
| 50-59 years | 192(21.3) | 4 (15.4) | 25 (19.1) | 2 (22.2) | 157 (22.4) | 2 (8.3) | 2 (18.2) |
| 60-69 years | 118 (13.1) | 5 (19.2) | 11 (8.4) | 1 (11.1) | 100 (14.2) | 0 (0) | 1 (9.1) |
| 70-79 years | 51 (5.6) | 0 (0) | 5 (3.8) | 1 (11.1) | 43 (6.1) | 2 (8.3) | 0 (0) |
| 80-89 years | 10 (1.1) | 0 (0) | 1 (0.8) | 0 (0) | 9 (1.3) | 0 (0) | 0 (0) |
| 90-99 years | 3 (0.3) | 0 (0) | 0 (0) | 0 (0) | 3 (0.4) | 0 (0) | 0 (0) |
| Indigenous self-identification | 10 (1.1) | 0 (0) | 1 (0.8) | 0 (0) | 7 (1) | 0 (0) | 2 (18.2) |
| Occupation |  |  |  |  |  |  |  |
| Technical services | 29 (3.2) | 0 (0) | 3 (2.3) | 0 (0) | 25 (3.6) | 0 (0) | 1 (9.1) |
| Education | 34 (3.8) | 0 (0) | 8 (6.1) | 0 (0) | 25 (3.6) | 1 (4.2) | 0 (0) |
| Healthcare | 117 (13) | 2 (7.7) | 12 (9.2) | 2 (22.2) | 97 (13.8) | 4 (16.7) | 0 (0) |
| Agricultural activities | 2 (0.2) | 0 (0) | 0 (0) | 0 (0.0) | 2 (0.3) | 0 (0) | 0 (0) |
| Commerce | 327 (36.2) | 12 (46.2) | 53 (40.5) | 0 (0.0) | 251 (35.8) | 8 (33.3) | 3 (27.3) |
| Other | 197 (21.8) | 5 (19.2) | 31 (23.7) | 6 (66.7) | 149 (21.2) | 5 (20.8) | 1 (9.1) |
| Unemployed | 17 (1.9) | 0 (0) | 0 (0) | 1 (11.1) | 15 (2.1) | 1 (4.2) | 0 (0) |
| Stay-at-home | 180 19.9) | 7 (26.9) | 24 (18.3) | 0 (0) | 138 (19.7) | 5 (20.8) | 6 (54.5) |
| Last-season flu vaccination | 222 (24.6) | 5 (19.2) | 38 (29) | 1 (11.1) | 173 (24.6) | 4 (16.7) | 1 (9.1) |
| Special populations |  |  |  |  |  |  |  |
| Children and adolescents (<18 years) | 11 (1.2) | 0 (0.0) | 3 (2.3) | 0 (0) | 7 (1) | 1 (4.2) | 0 (0) |
| Age, mean (SD) | 11.6 (5.5) | - | 11 (7.8) | - | 11.9 (5.5) | 12 | - |
| Last-season flu vaccination | 2 (18.2) | 0 (0) | 0 (0) | 0 (0) | 2 (28.6) | 0 (0) | 0 (0) |
| Non-pregnant/puerperal adults (≥18 years) | 890 (98.6) | 26 (100) | 128 (97.7) | 9 (100) | 693 (98.7) | 23 (95.8) | 11 (100) |
| Age, mean (SD) | 47.2 (14.3) | 44.5 (13.9) | 42.9 (13.6) | 51 (11.1) | 48.2 (14.2) | 43.9 (12.6) | 40 (12.9) |
| Last-season flu vaccination | 220 (24.7) | 5 (19.2) | 38 (29.7) | 1 (11.1) | 171 (24.7) | 4 (17.4) | 1 (9.1) |
| Comorbidities |  |  |  |  |  |  |  |
| Diabetes | 138 (15.3) | 0 (0) | 10 (7.6) | 2 (22.2) | 123 (17.5) | 3 (12.5) | 0 (0) |
| COPD | 12 (1.3) | 0 (0) | 2 (1.5) | 0 (0) | 10 (1.4) | 0 (0) | 0 (0) |
| Asthma | 23 (2.5) | 1 (3.8) | 3 (2.3) | 1 (11.1) | 18 (2.6) | 0 (0) | 0 (0) |
| Immunosuppression | 14 (1.6) | 0 (0) | 5 (3.8) | 0 (0) | 9 (1.3) | 0 (0) | 0 (0) |
| Hypertension | 166 (18.4) | 4 (15.4) | 11 (8.4) | 0 (0) | 147 (20.9) | 3 (12.5) | 1 (9.1) |
| HIV/AIDS | 2 (0.2) | 0 (0) | 0 (0) | 0 (0) | 2 (0.3) | 0 (0) | 0 (0) |
| Cardiovascular disease | 27 (1.6) | 2 (7.7) | 1 (0.8) | 1 (11.1) | 23 (3.3) | 0 (0) | 0 (0) |
| Obesity | 241 (3) | 9 (34.6) | 41 (31.3) | 2 (22.2) | 182 (25.9) | 3 (12.5) | 4 (36.4) |
| Chronic kidney disease | 17 (26.7) | 0 (0) | 0 (0) | 0 (0) | 17 (2.4) | 0 (0) | 0 (0) |
| Smoker | 107 (1.9) | 0 (0) | 20 (15.3) | 1 (11.1) | 82 (11.7) | 2 (8.3) | 2 (18.2) |
| Type of medical attention |  |  |  |  |  |  |  |
| Ambulatory | 675 (74.8) | 26 (100) | 127 (96.9) | 0 (0) | 491 (69.9) | 22 (91.7) | 9 (81.8) |
| Hospitalization | 228 (25.2) | 0 (0) | 4 (3.1) | 9 (10) | 211 (30.1) | 2 (8.3) | 2 (18.2) |
| Severity of the disease |  |  |  |  |  |  |  |
| Non-critical | 833 (92.2) | 26 (100) | 130 (99.2) | 4 (44.4) | 638 (90.9) | 24 (100) | 11 (100) |
| Critical | 70 (7.8) | 0 (0) | 1 (0.8) | 5 (55.6) | 64 (9.1) | 0 (0) | 0 (0) |
| Baseline symptoms |  |  |  |  |  |  |  |
| Fever | 643 (71.2) | 20 (76.9) | 82 (62.6) | 9 (100) | 511 (72.8) | 12 (50) | 9 (81.8) |
| Cough | 675 (74.8) | 17 (65.4) | 98 (74.8) | 7 (77.8) | 527 (75.1) | 17 (70.8) | 9 (81.8) |
| Sore throat | 434 (48.1) | 10 (38.5) | 63 (48.1) | 3 (33.3) | 338 (48.1) | 14 (58.3) | 6 (54.5) |
| Shortness of breath | 402 (44.5) | 11 (42.3) | 36 (27.5) | 6 (66.7) | 335 (47.7) | 9 (37.5) | 5 (45.5) |
| Irritability | 199 (22) | 5 (19.2) | 23 (17.6) | 1 (11.1) | 158 (22.5) | 6 (25) | 6 (54.5) |
| Diarrhea | 285 (31.6) | 9 (34.6) | 39 (29.8) | 1 (11.1) | 226 (32.2) | 4 (16.7) | 6 (54.5) |
| Chest pain | 330 (36.5) | 8 (30.8) | 37 (28.2) | 3 (33.3) | 266 (37.9) | 9 (37.5) | 7 (63.6) |
| Chills | 434 (48.1) | 12 (46.2) | 60 (45.8) | 3 (33.3) | 342 (48.7) | 11 (45.8) | 6 (54.5) |
| Headache | 645 (71.4) | 19 (73.1) | 93 (71) | 7 (77.8) | 502 (71.5) | 15 (62.5) | 9 (81.8) |
| Myalgias | 590 (65.3) | 20 (76.9) | 78 (59.5) | 5 (55.6) | 464 (66.1) | 15 (62.5) | 8 (72.7) |
| Arthralgias | 552 (61.1) | 16 (61.5) | 73 (55.7) | 5 (55.6) | 436 (62.1) | 16 (66.7) | 6 (54.5) |
| Abrupt deterioration | 556 (61.6) | 15 (57.7) | 77 (58.8) | 6 (66.7) | 440 (62.7) | 12 (50) | 6 (54.5) |
| Rhinorrhea | 294 (32.6) | 5 (19.2) | 45 (34.4) | 1 (11.1) | 229 (32.6) | 10 (41.7) | 4 (36.4) |
| Polypnea | 171 (18.9) | 3 (11.5) | 22 (16.8) | 1 (11.1) | 141 (20.1) | 2 (8.3) | 2 (18.2) |
| Vomit | 106 (11.7) | 2 (7.7) | 9 (6.9) | 1 (11.1) | 90 (12.8) | 3 (12.5) | 1 (9.1) |
| Abdominal pain | 153 (16.9) | 1 (3.8) | 17 (13) | 2 (22.2) | 127 (18.1) | 3 (12.5) | 3 (27.3) |
| Conjunctivitis | 157 (17.4) | 4 (15.4) | 20 (15.3) | 1 (11.1) | 123 (17.5) | 5 (20.8) | 4 (36.4) |
| Cyanosis | 67 (7.4) | 1 (3.8) | 7 (5.3) | 1 (11.1) | 56 (8) | 1 (4.2) | 1 (9.1) |
| Sudden onset of symptoms | 349 (38.6) | 7 (26.9) | 47 (35.9) | 5 (55.6) | 274 (3) | 12 (50) | 4 (36.4) |
| Concomitant use of antibiotics | 566 (62.7) | 22 (84.6) | 84 (64.1) | 5 (55.6) | 434 (61.8) | 15 (62.5) | 6 (54.5) |
| Timing of antiviral treatment |  |  |  |  |  |  |  |
| Before medical attention | 783 (86.7) | 19 (73.1) | 121 (92.4) | 7 (77.8) | 603 (85.9) | 23 (95.8) | 10 (90.9) |
| After medical attention | 120 (13.3) | 7 (26.9) | 10 (7.6) | 2 (22.2) | 99 (14.1) | 1 (4.2) | 1 (9.1) |
| ≤2 days after symptom onset | 579 (64.2) | 8 (30.8) | 99 (75.6) | 1 (11.1) | 448 (63.9) | 16 (66.7) | 7 (63.6) |
| >2 days after symptom onset | 323 (35.8) | 18 (69.2) | 32 (24.4) | 8 (88.9) | 253 (36.1) | 8 (33.3) | 4 (36.4) |
| Data expressed as frequency (%) or mean (SD)  SD: Standard deviation, COPD: Chronic obstructive pulmonary disease, HIV/AIDS: Human immunodeficiency virus/acquired immune deficiency syndrome. | | | | | | | |

Supplementary Table 21. Survival analysis of laboratory-confirmed COVID-19 patients according to early or late use of antivirals.

| **Group** | **Total** | **Deaths** | **Survivors** | **Survival** | **p value** |
| --- | --- | --- | --- | --- | --- |
| Early use of antivirals (≤2 days) | 579 | 64 | 515 | 88.9% | - |
| Type of antiviral | | | | | |
| No antiviral/antibiotic | 114143 | 7351 | 106792 | 93.6% | Reference |
| Acyclovir | 8 | 0 | 8 | 100% | 0.5 |
| Amantadine | 99 | 2 | 97 | 98% | 0.08 |
| Lopinavir-Ritonavir | 1 | 0 | 1 | 100% | 0.8 |
| Oseltamivir | 448 | 62 | 386 | 86.2% | <0.0001 |
| Rimantadine | 16 | 0 | 16 | 100% | 0.3 |
| Zanamivir | 7 | 0 | 7 | 100% | 0.5 |
| Late use of antivirals (>2 days) | 323 | 28 | 295 | 91.3% | - |
| Type of antiviral | | | | | |
| No antiviral/antibiotic | 114143 | 7351 | 106792 | 93.6% | Reference |
| Acyclovir | 18 | 0 | 18 | 100% | 0.3 |
| Amantadine | 32 | 1 | 31 | 96.9% | 0.4 |
| Lopinavir-Ritonavir | 8 | 2 | 6 | 75% | 0.03 |
| Oseltamivir | 253 | 15 | 228 | 90.1% | 0.04 |
| Rimantadine | 8 | 0 | 8 | 100% | 0.5 |
| Zanamivir | 4 | 0 | 4 | 100% | 0.6 |
| Data expressed as number of patients or percentual of survival. P values were calculated by Log-Rank test. | | | | | |

Supplementary Figure 1. Density functions of propensity score-matched patients treated with antivirals and controls before and after matching.

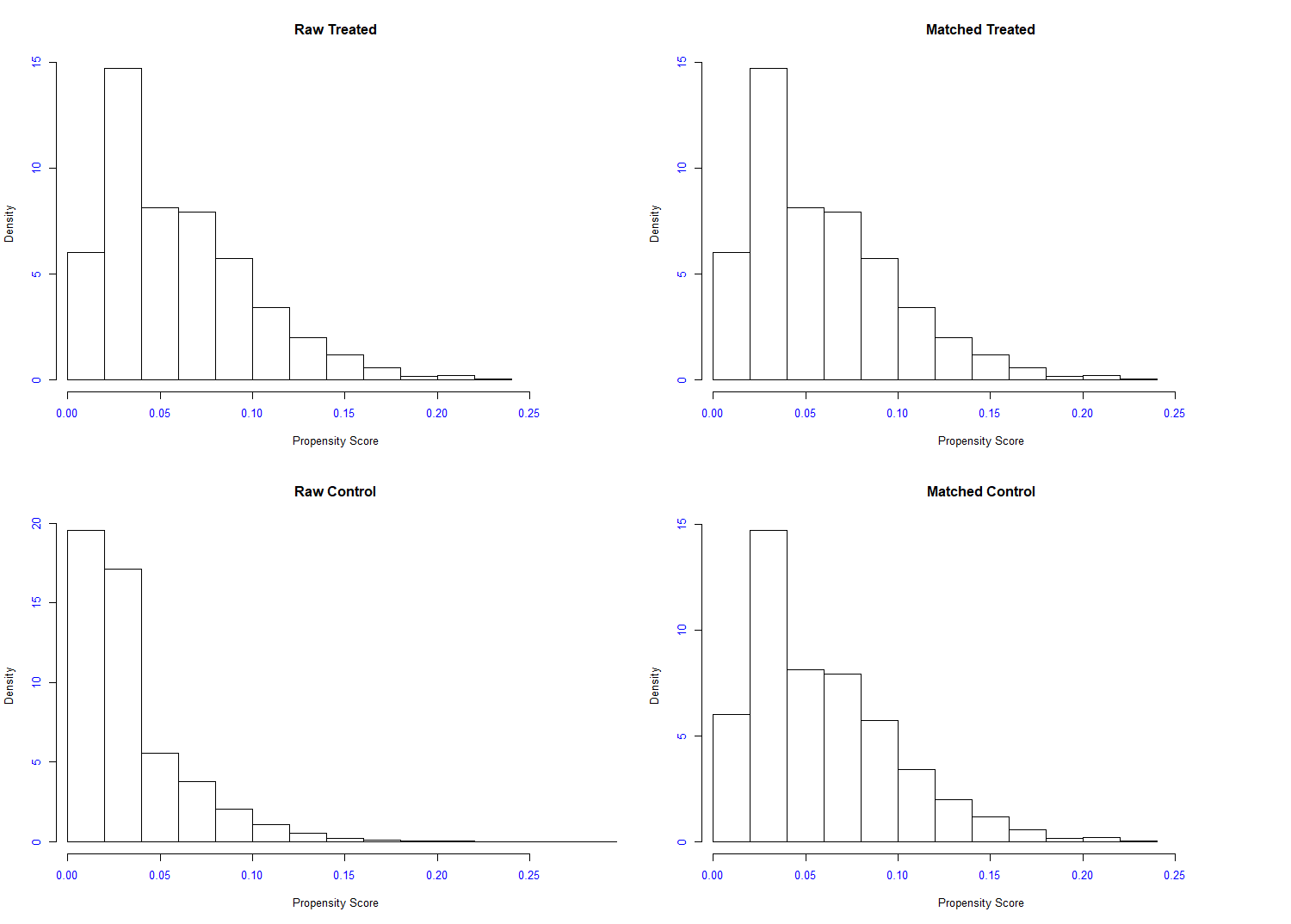

Supplementary Figure 2. Density functions of propensity score-matched patients treated with Oseltamivir and controls before and after matching.

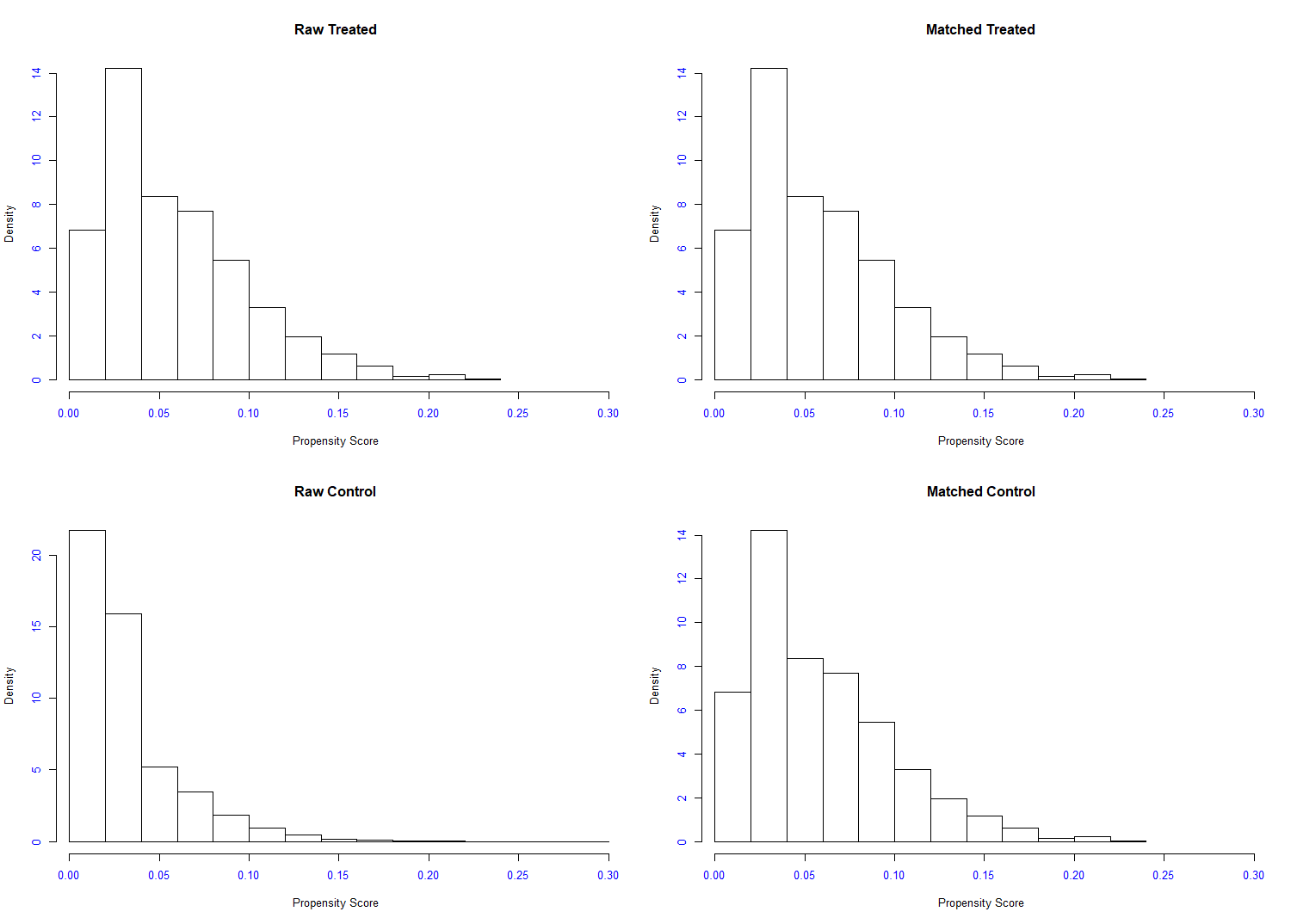

Supplementary Figure 3. Density functions of propensity score-matched patients treated with antibiotics and controls before and after matching.

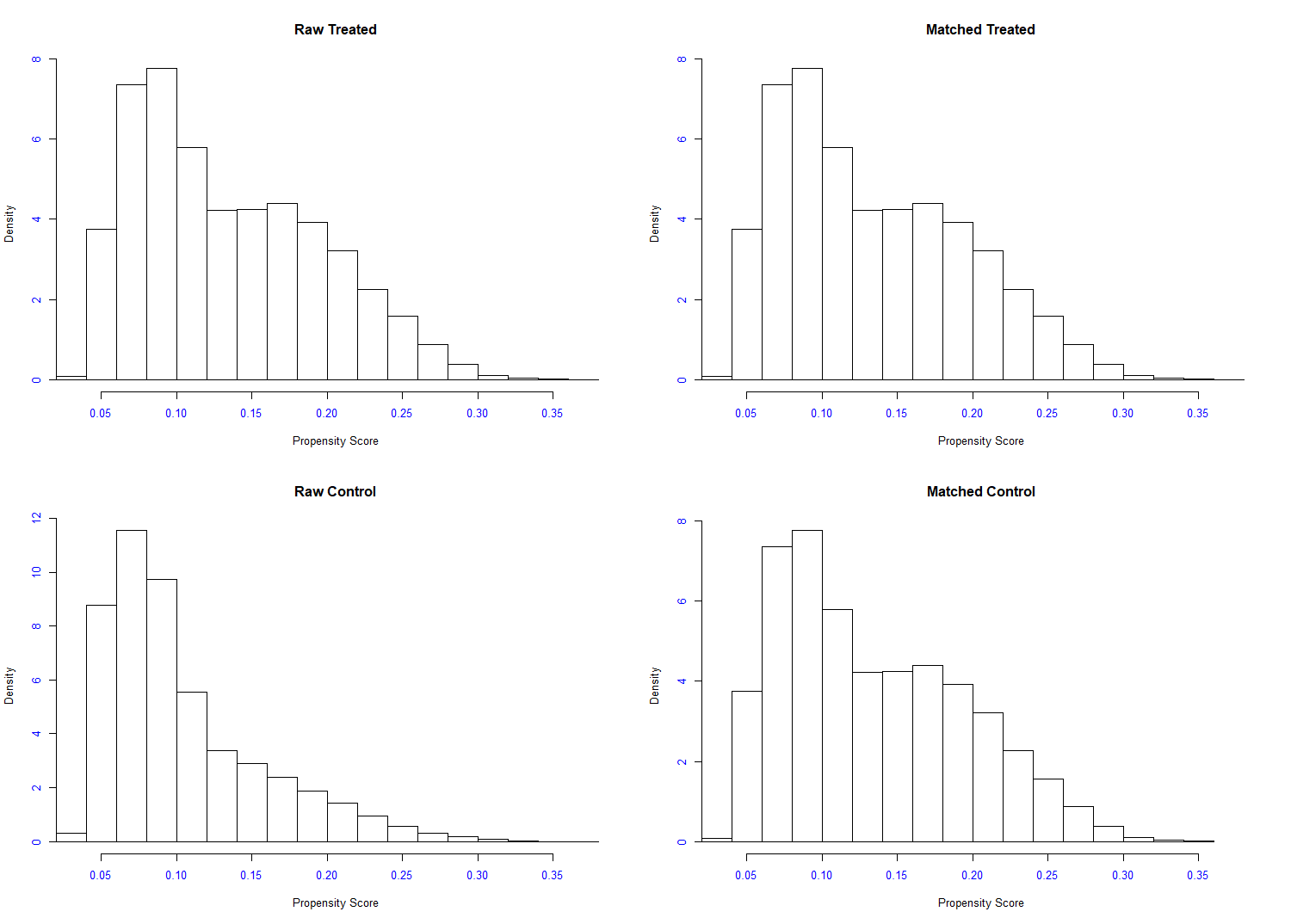

Supplementary Figure 4. Time from symptom onset to initiation of antivirals (A) and time from initiation of antivirals to hospitalization (B) in 903 patients with laboratory-confirmed COVID-19 receiving medical attention in one of 154 accredited medical units in Mexico City.

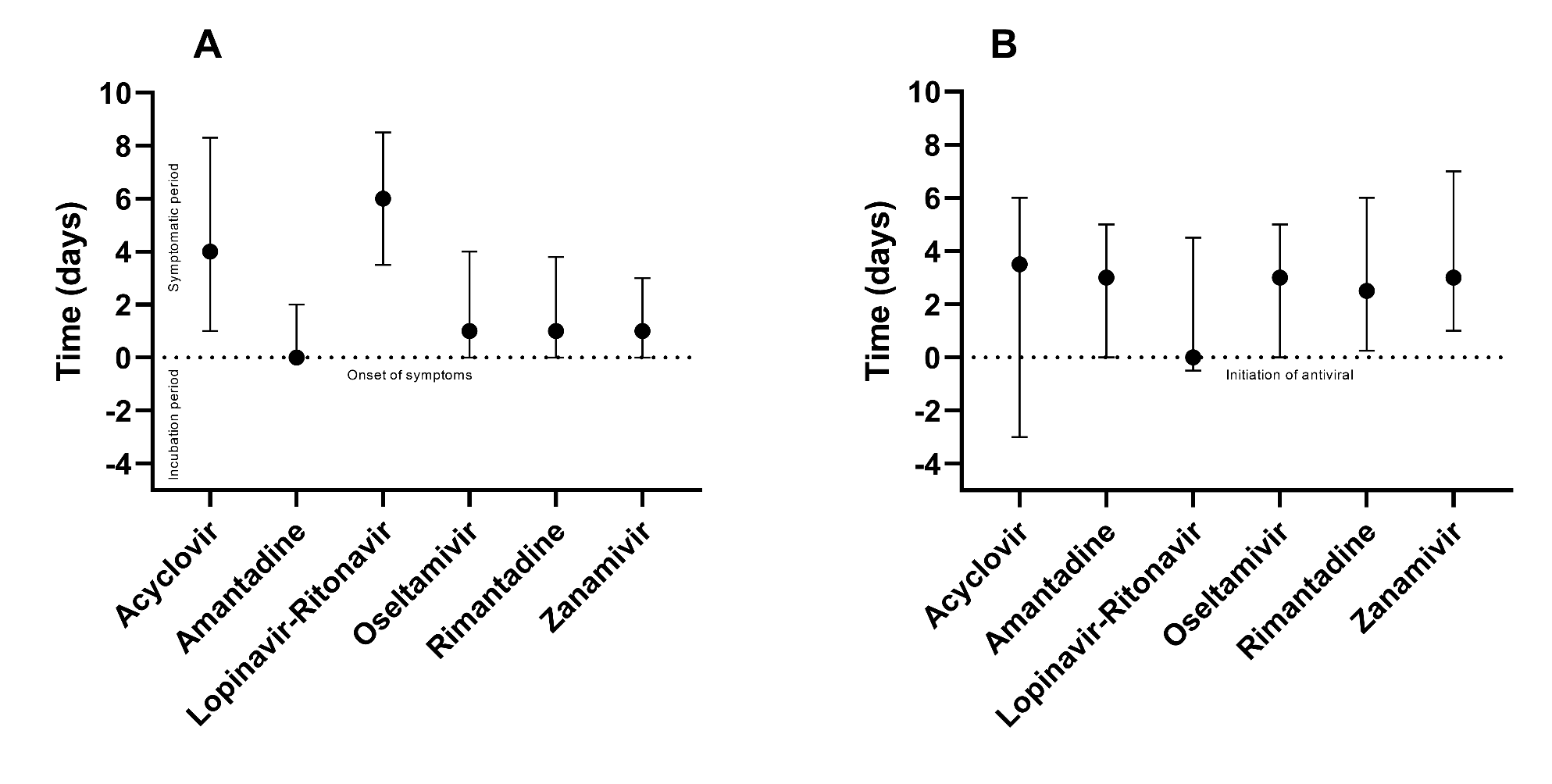

Supplementary Figure 5. Comparisons of hospitalization rates in 903 patients with laboratory-confirmed COVID-19 receiving medical attention in one of 154 accredited medical units in Mexico City who received early (≤2 days) or late (>2 days) antivirals.

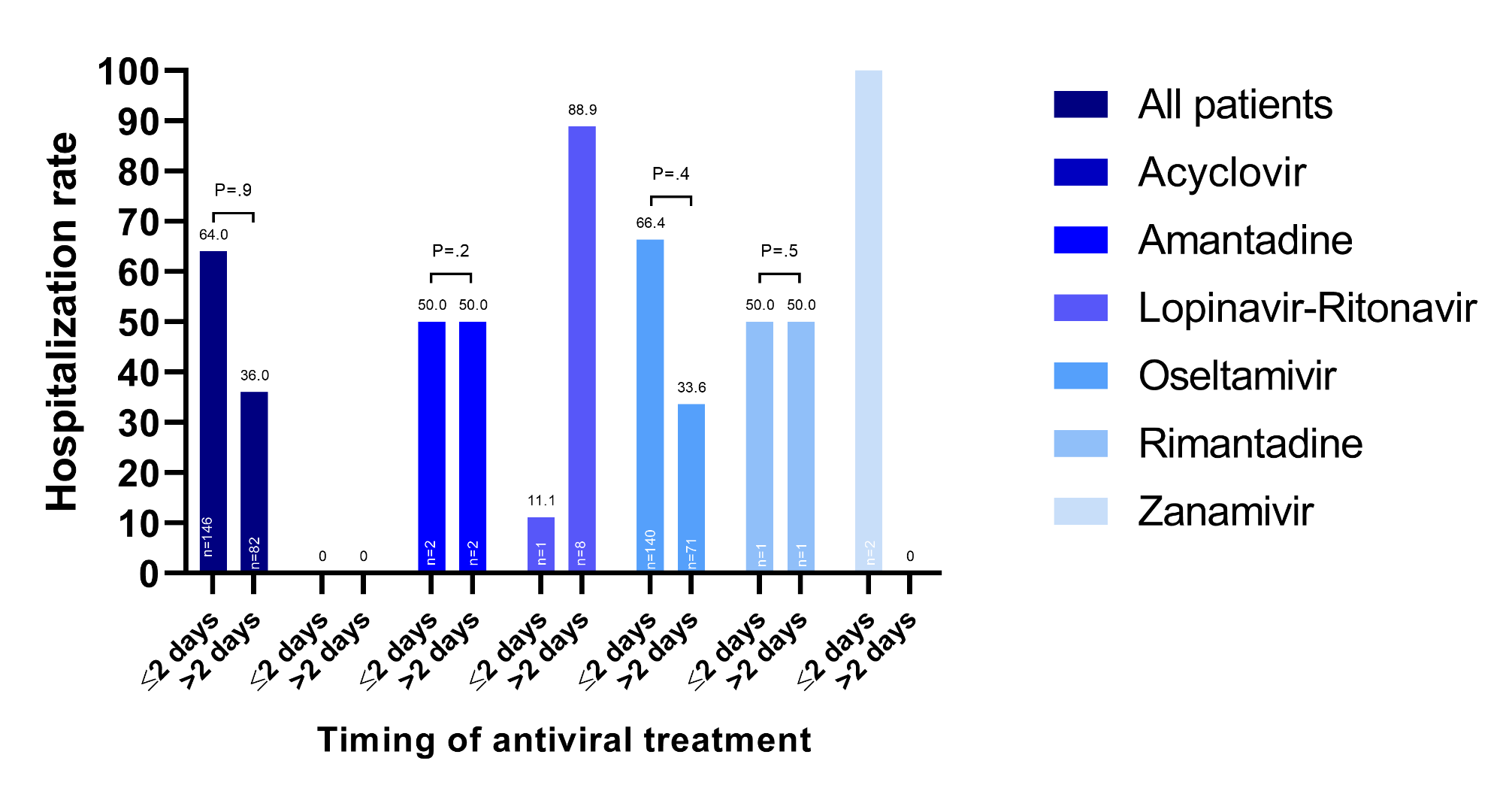
