## Supplementary material for "All-cause mortality among patients treated with repurposed antivirals and antibiotics for COVID-19 in Mexico City: A Real-World Observational Study": Upon evaluating a patient suspected of having COVID-19, healthcare professionals are required to fill out a format (Supplementary Appendix) containing

SALUD Secretaría de Salud | IMSS | ISSSTE | PEMEX | SEDENA Secretaría de la Defensa Nacional | SEMAR Secretaría de Marina | SNDIF Sistema Nacional DIF | INPI Instituto Nacional de los Pueblos Indígenas

### Epidemiologic study of suspected viral respiratory disease

#### GENERAL INFORMATION

Name of medical unit: ______

Date of registration in the platform: ______ dd/mm/yyyy Platform number: ______

Last Name: ______ Second Last Name: ______ Name(s): ______

Date of birth: Day: ____ Month: ____ Year: ____ CURP: ______

Sex: Man: ____ Woman: ____ Pregnant patient: Yes ____ No ____ Months of pregnancy: ____ Patient in puerperium Yes ____ No ____ Days of puerperium ____

Nationality: Mexican: ____ Foreigner: ____ Migrant: Yes ____ No ____ Country of nationality: ______ Country of origin: ______

Visited countries in the last three months: 1 ____ 2 ____ 3 ____ Other: ____ Date of entry into Mexico: ______

Country of birth: ______ Place of birth (state): ______

Place of residency (state): ______ Municipality of residence: ______

Locality: ______

Street: ______ Number: ____

Between streets: ______ & ______

Suburb: ______ PC: ____ Telephone: ______

Indigenous self-identification: Yes ____ No ____ Speaker of indigenous language: Yes ____ No ____

Occupation: ______

Is the patient affiliated to an educational institution? ______

#### CLINICAL DATA

Admitted to (service): ______ Type of attention: 1=Ambulatory ____ 2=Hospitalization ____

Date of admission: ______ dd/mm/yyyy Date of symptom onset: ______ dd/mm/yyyy

Since the onset of symptoms, has the patient…

…had any of the following signs or symptoms?

| | Yes | No |
|---|---|---|
| Sudden onset of symptoms | | |
| Fever | | |
| Cough | | |
| Headache | | |
| Shortness of breath | | |
| Irritability | | |
| Chest pain | | |
| Chills | | |
| Sore throat | | |
| Myalgias | | |
| Arthralgias | | |
| Anosmia | | |
| Dysgeusia | | |
| Rhinorrhea | | |
| Conjunctivitis | | |

Comorbidities

| | Yes | No |
|---|---|---|
| Diabetes | | |
| COPD | | |
| Asthma | | |
| Immunosuppression | | |
| Hypertension | | |
| HIV/AIDS | | |
| Cardiovascular disease | | |
| Obesity | | |
| Chronic kidney disease | | |
| Smoker | | |
| Other | | |

Specify any others: ______

| Other symptoms | Yes | No |
|---|---|---|
| Abrupt deterioration | | |
| Diarrhea | | |
| Polypnea | | |
| Abdominal pain | | |
| Vomit | | |
| Cyanosis | | |

Probable diagnosis: 1=Influenza-like illness (ILI)* ____ 2=Severe acute respiratory infection (SARI) ____

*ILI is a mild respiratory disease

#### TREATMENT

Since the onset of symptoms, has the patient…

…been treated with antipyretics? Yes ☐ No ☐

…been treated with antivirals? Yes ☐ No ☐

If the answer was yes:

Specify the antiviral: ☐ 1=Amantadine 2=Rimantadine 3=Oseltamivir 4=Zanamivir 5=Other, specify: ☐

When was the antiviral medication started? ☐ dd/mm/yyyy

In the medical unit…

…were antibiotics started? Yes ☐ No ☐

…were antivirals started? Yes ☐ No ☐

Specify the antiviral: ☐ 1=Amantadine 2=Rimantadine 3=Oseltamivir 4=Zanamivir 5=Other, specify: ☐

#### EPIDEMIOLOGICAL HISTORY

Has the patient had any contact with persons with respiratory disease in the last two weeks? Yes ☐ No ☐

During the weeks prior to symptom onset did the patient have contact with:

| | Yes | No |
|---|---|---|
| Birds: | | |
| Swine: | | |

Other animals: ☐

Recent travels in the 7-day period before the onset of signs/symptoms: Yes ☐ No ☐

Country: ☐ City: ☐

Did the patient receive last-year influenza vaccination? Yes ☐ No ☐

Date of vaccination: ☐ dd/mm/yyyy

#### LABORATORY

Was a sample for testing obtained? Yes ☐ No ☐

Laboratory that will be processing the sample: ☐

Site of sample: ☐ 1=Pharyngeal swab 2=Nasopharyngeal swab 3=Bronchoalveolar lavage 4=Pulmonary biopsy

Date sample was obtained: ☐ dd/mm/yyyy

Result: ☐

#### FOLLOW-UP

Follow-up status: ☐ 1=Discharged 2=Undergoing treatment/Referred/Domiciliary follow-up/End of follow-up 3=Severe case 4=Mild case 5=Death*

In case of discharge: Indicate cause of discharge: ☐ 1=Improvement 2=Curation 3=Voluntary 4=Transferred

Has the patient been admitted to an ICU at any moment of the disease? Yes ☐ No ☐

Has the patient been put on invasive mechanical ventilation at any moment of the disease? Yes ☐ No ☐

Has the patient been diagnosed with pneumonia at any moment of the disease? Yes ☐ No ☐

Date of discharge: ☐ dd/mm/ yyyy

Specify if this is a positive COVID-19 case by association or clinical-epidemiological ruling: *

** Mark with an X only one of the following options*

**a.** Confirmed COVID-19 by clinical-epidemiological association ☐

**b.** Confirmed COVID-19 by clinical-epidemiological ruling (applies only for deaths) ☐

**c**. No (none of the previous options) ☐

Death: Date of death: ☐ dd/mm/yyyy

Death Certificate (number): ☐ ***Death due to influenza or COVID-19** Yes ☐ No ☐

*Attach a copy of the death certificate if the patient met criteria for suspected viral respiratory disease case

Name and position of person registering data

______________________________

______________________________

Name and position of authorizing person

______________________________

______________________________

Date of elaboration: ☐ dd/mm/yyyy
